# Differential Impacts of COVID-19 Lockdowns on PM_2.5_ across the United States

**DOI:** 10.1101/2021.03.10.21253284

**Authors:** Kevin L. Chen, Lucas R.F. Henneman, Rachel C. Nethery

## Abstract

The COVID-19 pandemic has induced large-scale social, economic, and behavioral changes, presenting a unique opportunity to study how air pollution is affected by unprecedented societal shifts. At each of 455 PM_2.5_ monitoring sites across the United States, we conduct a causal inference analysis to determine the impacts of COVID-19 interventions and behavioral changes (“lockdowns”) on PM_2.5_ concentrations. Our approach allows for rigorous confounding adjustment and provides highly spatio-temporally resolved effect estimates. We find that, with the exception of the Southwest, most of the US experienced increases in PM_2.5_ during lockdown, compared to the concentrations expected under business-as-usual. To investigate possible drivers of this phenomenon, we use regression to characterize the relationship of many environmental, geographical, meteorological, mobility, and socioeconomic factors with the lockdown-attributable changes in PM_2.5_. Our findings have immense environmental policy relevance, suggesting that large-scale mobility and economic activity reductions may be insufficient to substantially and uniformly reduce PM_2.5_.

## Introduction

Acute widespread social, behavioral, and economic changes have occurred across the United States in the wake of the COVID-19 pandemic. Most notably, mobility levels decreased substantially^1–3^ nationwide during the initial “lockdown period” in March and April 2020 as a result of changes in human behavior and other nonpharmaceutical interventions, such as government-imposed stay-at-home orders, in response to the rapid spread of the virus. The unprecedented actions taken to curb the spread of COVID-19 have created a unique “quasi-experiment” that can be leveraged to study the effect of large-scale behavioral change on air quality.

Exposure to fine particle matter, PM_2.5_, has been shown to have significant adverse health effects, including respiratory and cardiovascular morbidity and mortality^4–6^. To create effective policy to limit PM_2.5_ exposure, it is crucial to understand the impact of reductions in certain emission-generating human behaviors on ambient concentrations. Studies of other, smaller scale quasi-experiments have provided some of the strongest evidence for the health impacts of air pollution and effective reduction strategies, e.g., the ban of bituminous coal in Dublin^7^, restrictions on transportation and industrial activities during the 1996 Atlanta^8^ and 2008 Beijing Olympic Games^9,10^.

Several recent studies, some of which are in pre-print at the time of writing, conduct analyses of the lockdown effects on air quality using data from a single or a few monitors^11^ or using nationwide and/or statewide averages of monitored values^12^. However, due to the vast differences across space in source-specific contributions to PM_2.5_, the crudeness of these aggregated analyses limits their policy relevance. Additionally, a study of 122 US counties by Berman and Ebisu^13^ reports a decline in PM_2.5_, on average, during the lockdown period. However, this study did not report or analyze trends over space, nor did they consider the environmental policy implications of their results. In our study, we estimate the effects of the lockdown on PM_2.5_ concentrations at each of 455 individual monitors in the Environmental Protection Agency (EPA) monitoring network located throughout the contiguous US. The high spatial resolution of our effect estimates provides more specific policy insights and enables deeper investigation into the factors influencing lockdown-related changes in PM_2.5_.

Even in the context of quasi-experimental conditions, isolating the effects of COVID-19 interventions on air pollution in general cannot be achieved through simple before-and-after comparisons nor comparison to a concentrations in the previous year, strategies that are used in much of the existing literature and oft-cited in news media. Numerous time-varying factors that influence air pollution levels, both observable and unobservable, are unaccounted for with such approaches, e.g., meteorology, year-to-year trends, and seasonality. For example, PM_2.5_ levels have decreased 44% in the US since 2000^14^. More rigorous approaches are needed to account for pollution trends and time-varying confounders as a means of truly characterizing the causal effects of pandemic-related behavioral changes.

The aims of our study are best represented as a two-stage approach: first, we estimate the lockdown-attributable daily changes in PM_2.5_ concentrations at 455 monitoring sites using a causal inference approach that can adjust for both observed and unobserved time-varying contributors to PM_2.5_. Formally, at each monitor, we estimate the PM_2.5_ concentrations that would have been expected each day in the absence of the COVID-19 pandemic and subsequent interventions (the “counterfactual”) and compare these to the corresponding observed levels at the monitor during the lockdown. Second, we use these effect estimates to identify environmental, geographical, meteorological, mobility, and socioeconomic factors that are associated with changes in PM_2.5_ during the lockdown. We also quantify the impact of these short-term PM_2.5_ changes on respiratory and cardiovascular disease hospitalizations using the EPA’s Core Particulate Matter Health Impact Functions.

## Results

We define the “lockdown period” for each ground monitor site as beginning on the day of the corresponding state’s state-of-emergency declaration and ending on the earlier of April 30th, 2020 or the day businesses began to reopen in that state (see Extended Data Table E1 for the state-specific dates). We estimate the lockdown-attributable changes in PM_2.5_ at each of 455 EPA ground monitor sites using a causal inference approach called the Synthetic Control Method (SCM)^15–17^, which leverages a pre/post-intervention study design to estimate intervention effects adjusted for both time-varying and time-invariant unmeasured confounders under mild assumptions.

We use SCM to estimate the “counterfactual PM_2.5_ concentrations” during the lockdown period, i.e., the daily concentrations that would have been expected during the 2020 lockdown period in the absence of the lockdown or any non-mandated personal behavioral changes that took place due to COVID-19 during same period. Briefly, for a given monitor site, SCM creates a time series of synthetic 2020 daily PM_2.5_ concentrations by forming a weighted average of the year-specific time series of daily PM_2.5_ concentrations for 2010-2019. The weights are selected to result in a synthetic 2020 time series that provides the closest approximation of the 2020 pre-lockdown observed time series of PM_2.5_ concentrations. The values of the synthetic times series during the lockdown represent our best guess at what PM_2.5_ concentrations would have been under a “business-as-usual” scenario, simultaneously accounting for daily, seasonal, and long-term PM_2.5_ trends. The values in this synthetic 2020 time series during the lockdown period are thus taken as estimates of the daily counterfactual PM_2.5_ concentrations. We then estimate daily lockdown-attributable changes in PM_2.5_ concentrations by taking the difference between the observed and estimated counterfactual PM_2.5_ levels for each day during the lockdown period. Cumulative estimates of lockdown-attributable changes in PM_2.5_ reported hereafter are averages of these daily effect estimates across all days in the lockdown period for the specified monitor. For consistency with the causal inference literature, we refer to these estimates as “average treatment effects on the treated” (ATTs).

Figure 1 maps the estimated lockdown-attributable changes in PM_2.5_ concentrations (ATTs) at each monitoring site as well as showing a smoothed map of these estimates, interpolated using inverse distance weighting. Negative ATT values, depicted in green, indicate that the location experienced a lockdown-attributable decrease in PM_2.5_ concentrations, while positive (orange/red) values represent lockdown-attributable increases.

**Figure 1.**
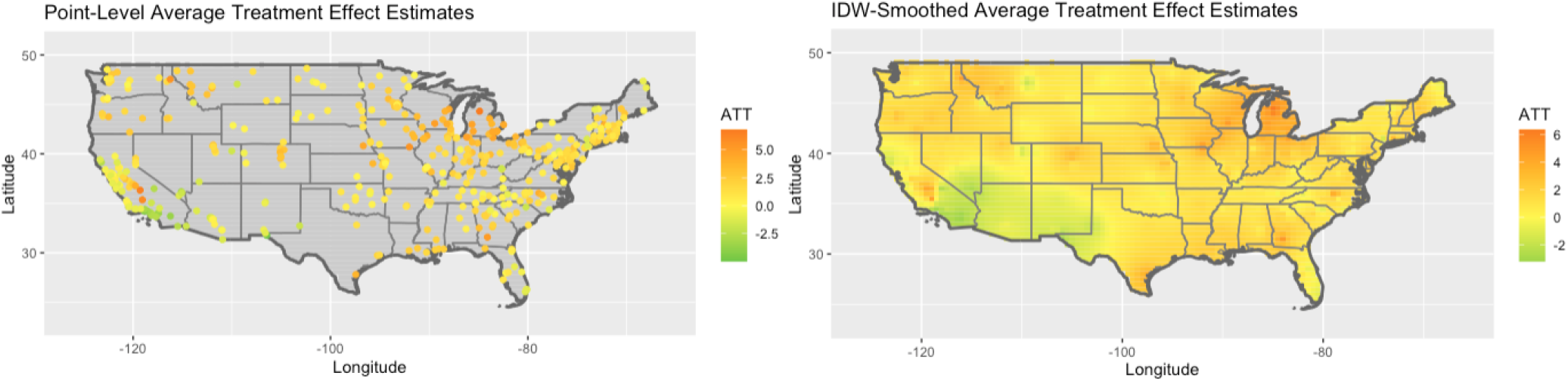
(Left) Monitor-level estimated average treatment effect on the treated (“ATT”), in *µ*g/m^3^; i.e., the average PM_2.5_ change attributable to COVID-19 interventions over the lockdown period. (Right) ATTs (in *µ*g/m^3^) smoothed across the US using inverse distance weighting.

We find that, during the COVID-19 lockdowns, PM_2.5_ increased across most of the US compared to what would have been expected under a business-as-usual scenario. We henceforth refer to smaller lockdown-attributable increases or larger lockdown-attributable decreases as “smaller increases”, and larger lockdown-attributable increases or smaller lockdown-attributable decreases as “larger increases”. The maps suggest that any lockdown-attributable decreases in PM_2.5_ are generally limited to areas of the Western and Southwestern United States. However, substantial lockdown-attributable increases were observed in much of the South, Midwest, and Pacific Northwest. Stratifying by US Census-defined regions, we find average region-wide increases of 1.19 *µ*g/m^3^, 1.11 *µ*g/m^3^, 1.94 *µ*g/m^3^, and 0.80 *µ*g/m^3^ for the Northeast, South, Midwest, and West, respectively. Over the entire country, we observe an average increase of 1.36 *µ*g/m^3^ attributable to pandemic interventions.

The formation of PM_2.5_ is known to be a very complex process– along with being directly emitted, much of it is formed secondarily in the atmosphere from other pollutants. To better understand factors that may have contributed to the heterogeneity in lockdown-attributable PM_2.5_ changes detected by our study (Figure 1), we investigate associations between the monitor-level effect estimates and the environmental, geographical, regional, meteorological, mobility, and socioeconomic conditions in the area surrounding the monitor. County-level measures of each feature are obtained and each monitor is assigned the features of the county in which it lies. Using the estimated lockdown-attributable PM_2.5_ changes as the outcome, we fit a linear regression model including all of the following features as predictors: 2017 primary PM_2.5_ emissions from various sources (residential, industrial processes, industrial boilers, dust, agriculture, and mobile)^18^; socioeconomic and demographic variables (e.g. poverty rate, proportion of population age 65+, population density)^19^; county’s average relative mobility change during the lockdown period^20^; level of urbanization^21^; and indicators for region.

Using this model, we find that industrial boiler emissions and population density have significant positive associations with lockdown-attributable increases in PM_2.5_, while a significant negative association was observed for relative mobility decrease, proportion of population age 65+, and the West region. In addition, areas not classified as “large central metro”, the most urban characterization, have significantly positive associations with lockdown-attributable increases in PM_2.5_. A summary of model estimates is provided in Figure 2.

**Figure 2.**
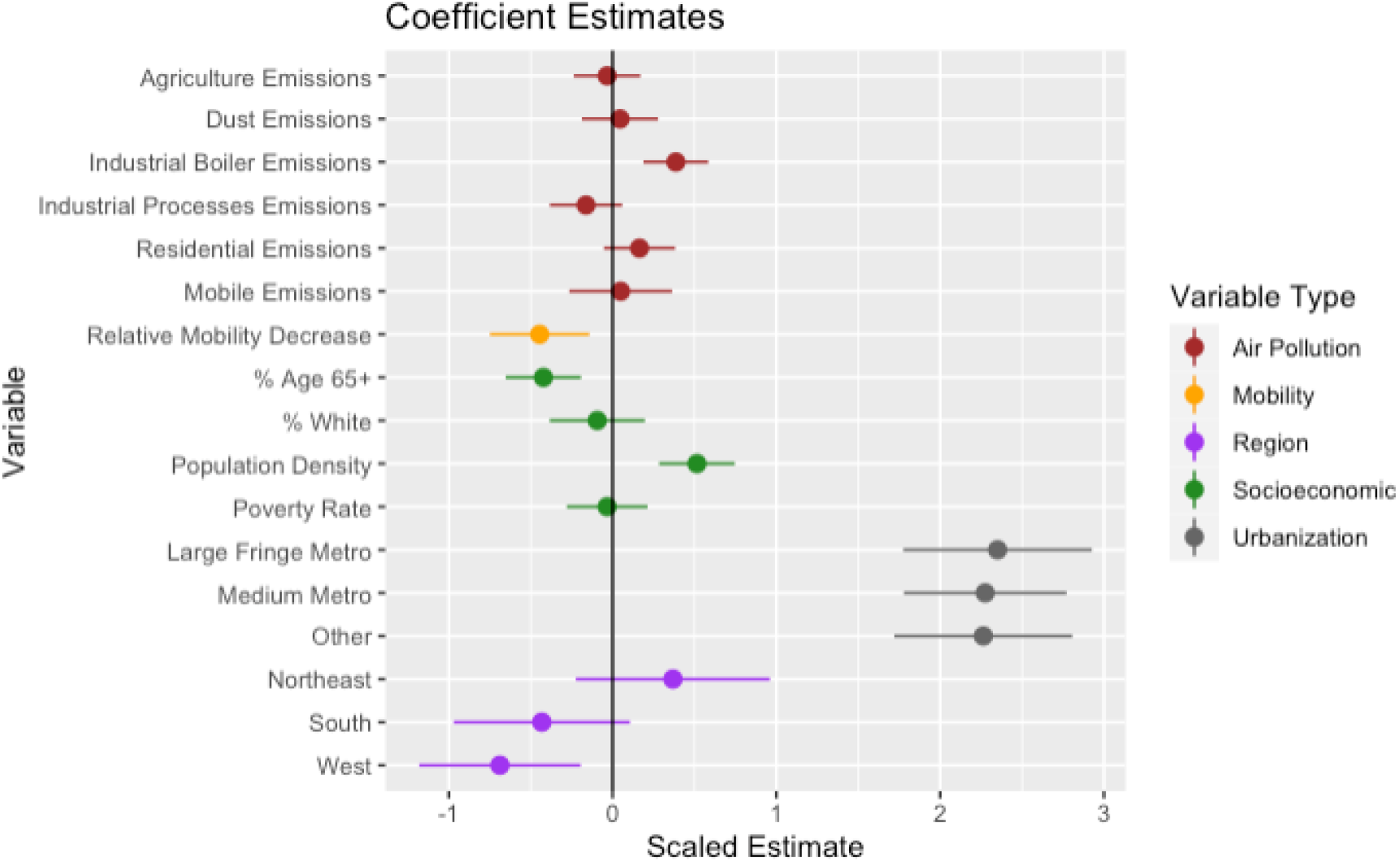
Coefficient estimates and 95% confidence intervals from linear regression model.

Although areas with larger decreases in mobility tended to experience smaller increases in PM_2.5_, our results suggest that any positive air quality impacts of these large-scale mobility decreases were insufficient to offset other non-meteorological factors promoting PM_2.5_ formation. Supporting this finding is Table 1, which shows the amount of PM_2.5_ emissions for each US Census region stratified by source^22^. Regions with higher emissions from stationary sources (i.e. fuel combustion sources such as power plants, solvents, agriculture, and other industrial processes) correspond to areas in which we found higher propensity for lockdown-attributable increases in PM_2.5_ during the lockdown. Additionally, mobile sources have a comparatively much smaller impact on PM_2.5_ emissions (Table 1).

**Table 1.** Sources of PM_2.5_ emissions by US Census region, in tons, with corresponding proportion of total regional PM_2.5_ emissions in parentheses from the EPA 2014 National Emissions Inventory^22^.

| Type | Northeast | South | Midwest | West | US Total |
| --- | --- | --- | --- | --- | --- |
| Stationary | 260,853 (82.7%) | 1,181,440 (60.9%) | 1,262,981 (81.1%) | 568,122 (42.4%) | 3,273,396 (63.5%) |
| Mobile | 41,432 (13.1%) | 129,699 (6.9%) | 97,710 (6.3%) | 64,945 (4.9%) | 333,786 (6.5%) |
| Other | 13,091 (4.2%) | 629734 (32.4%) | 196,898 (12.6%) | 705,595 (52.7%) | 1,545,318 (30.0%) |

We also investigate these lockdown-attributable PM_2.5_ changes relative to a “baseline” PM_2.5_ level prior to the pandemic. For each monitor site, we calculate the baseline as the average PM_2.5_ concentration observed in the month of April for 2017-2019. In a linear model, we find a nonsignificant, negative association (−0.076, 95% CI [-0.165, 0.014]) between baseline PM_2.5_ and the estimated lockdown-attributable changes in PM_2.5_.

We also assess the sensitivity of our findings to a 1-2 day spike in PM_2.5_ of unknown origin that we observed during the lockdown period for many monitors in the Midwest. To do so, we identify and remove these outlier days, defined as days on which PM_2.5_ levels spiked to 35 *µg/m*^3^ or higher, from our ATT estimates for all monitors. However, the removal of these outliers did not significantly affect the results (see Extended Data Figure E2).

We calculate the expected impacts of the estimated lockdown-attributable changes in PM_2.5_ on respiratory and cardiovascular disease hospitalizations in the age 65+ population for each county in the US, using the EPA’s Core Health Impact Functions for Particulate Matter^23^. We sum these changes across counties within each US region to obtain the results displayed in Table 2. While these figures may not be truly representative of health impacts during the lockdown due to the numerous changes in the public health sphere catalyzed by the pandemic, they are included in order to quantify the scope of the potential impacts under such changes in air pollution. These estimates are small relative to the overall PM_2.5_ burden in the US^24^, as expected given the short time-scale and the varying directions of the PM_2.5_ changes.

**Table 2.** Estimates of changes in respiratory and cardiovascular disease hospitalizations for populations age 65+ stratified by US census region, both over the lockdown period (“LD”) and in the hypothetical scenario that the PM_2.5_ changes persisted for one year.

| Region | Respiratory (LD) | CVD (LD) | Respiratory (Yearly) | CVD (Yearly) |
| --- | --- | --- | --- | --- |
| Northeast | 35 | 64 | 256 | 469 |
| South | 57 | 105 | 416 | 764 |
| Midwest | 83 | 153 | 609 | 1117 |
| West | -14 | -26 | -104 | -191 |
| US Total | 161 | 296 | 1177 | 2160 |

There are a number of limitations of our study. First, due to sporadic data collection at certain monitor sites, there is a considerable amount of missing PM_2.5_ data, particularly in prior years that are used to establish expected PM_2.5_ in the absence of the lockdown. However, much of the missingness is by-design due to EPA monitoring procedures, and we have set fairly stringent monitor inclusion criteria for our analyses to ensure that observed data are sufficient to capture important trends. In addition, our models are flexible enough to account for some missing data and we have visually inspected our results to ensure suitable model fit (Supplementary Figure 1). Furthermore, the heterogeneity of lockdown procedures, stringency, and adherence to advisories creates difficulty in a clear definition of the lockdown period. However, the time period we have defined was the period of the most acute lockdowns and we have further mitigated this issue by using consistent criteria for each state.

Our study has found that the impacts of the 2020 COVID-19 lockdowns on PM_2.5_ levels vary dramatically across the US, with strong regional trends. In the Midwestern and Southern regions, we unexpectedly observe consistent increases in PM_2.5_ compared to expected levels absent the lockdowns, while some areas in the Southwest experienced substantial decreases. The results of this important quasi-experiment provide evidence that policies to limit individual-level emissions-generating behaviors like mobility may not, alone, reduce PM_2.5_ levels, particularly in certain regions and when accompanied by other social and economic changes. Regulation of stationary emissions sources may be necessary to meaningfully and uniformly reduce PM_2.5_ and improve human health.

## Methods

All data preparation and analyses are conducted in R statistical software version 4.0.2.

The state-level lockdown period dates used throughout this study are given in Extended Data Table E1. Daily average PM_2.5_ concentrations between 2010 and 2019 from 1,580 monitor sites in the continental US are obtained from the EPA Air Quality System (AQS)^25^. For 2020, daily average PM_2.5_ concentrations are obtained from the EPA AirNow system^25^, where monitor data are made available prior to their integration into AQS, which occurs approximately twice per year. Daily meteorological factors (total precipitation, maximum temperature, maximum relative humidity, wind speed, and wind direction), day of the week, and season are obtained from the Google Earth Engine^26^ and merged with the PM_2.5_ data. Because some monitors have prohibitive amounts of missing measurements, we set inclusion criteria to select the monitors with sufficient data to establish the time trends needed for our analyses. Starting with 1,580 monitor sites with PM_2.5_ measurements between 2010-2020, we remove monitors (1) with no PM_2.5_ measurements during the defined lockdown period for their respective state, roughly mid-March to late-April of 2020 (918 monitors); (2) with less than 30 days of data during the lockdown period (23 monitors); (3) with no data prior to 2016 (117 monitors); or (4) with data entirely missing for five or more total years 2010-2019 (67 monitors). After applying these exclusion criteria, 455 monitors remain to be used in our analyses.

We rely solely on observed data for our analysis. Our goal is to estimate the daily PM_2.5_ concentrations that would have been expected at each monitoring site during the lockdown period under a business-as-usual scenario (i.e., without the lockdowns or the non-mandated personal behavioral changes that took place due to COVID-19 during the same period). Hereafter we refer to these as the “counterfactual PM_2.5_ concentrations”. As a result of long-term, seasonal and daily trends in PM_2.5_, complex meteorological variability, and other potential unobserved confounding factors, PM_2.5_ concentrations from a single previous year cannot be directly utilized to infer the counterfactual PM_2.5_ concentrations, nor can a simple average of PM_2.5_ concentrations over multiple years of historical data. We therefore use a causal inference approach often referred to as the “synthetic control method” (SCM) to estimate counterfactual PM_2.5_ concentrations^15–17^.

SCM was created to analyze the effects of a policy or intervention on an outcome using a case study. It leverages time series containing pre- and post-intervention outcome data from (1) a single unit that received an intervention (the “treated” unit) and (2) a set of control units that did not receive the intervention. Conceptually, using the pre-intervention data from both treated and control units, it creates a weighted average of the time series from the control units that best captures the pre-intervention trends in the time series for the treated unit. Then that same weighted average of the control units’ outcomes is used to estimate the outcome that would have been expected in the treated unit during the post-treatment period, in the absence of the intervention (the counterfactual outcome). This weighted average created by SCM is called a “synthetic control”. Formally, the optimal weights are identified by obtaining a latent factor representation of the multivariate time series data. SCM is flexible enough to account for both time-varying and time-invariant confounders of the intervention effect under mild assumptions. In addition, it accounts for any remaining pre-lockdown missing data by imputation.

In the classic SCM framework, for a given monitor, we consider each year 2010-2020 to be a “unit”, and the time series of PM_2.5_ concentrations for each year are the outcomes. 2020 is considered the treated unit and all other years are controls. Thus, SCM will create a weighted average of daily 2010-2019 PM_2.5_ concentrations at that monitor, and use that weighted average to estimate the counterfactual daily PM_2.5_ concentrations during the lockdown period.

To create a proper synthetic control, we must ensure that the time series from each year are aligned so that the day represented at a given position in the time series is comparable across years. Because PM_2.5_ exhibits weekday and weekend trends that must be accounted for when creating the synthetic control, we align the time series based on day-of-week rather than day-of-year. In particular, we let the time series for each year start on the first Monday of the year, so that aligning entries in the time series represent the same day-of-week and only a few days difference in day-of-year.

We implement SCM separately on the data from each of the 455 monitors using the *gsynth* package in R^16^ with a matrix completion estimator^17^. In addition to the default latent factor representation used by SCM, we include in the model fixed effects for year and time series position and adjust for both weather (maximum temperature, maximum relative humidity, precipitation, wind speed, and wind direction) and seasonality (month of year) as time-varying covariates. See Supplementary Figure 1 for monitor-level model fit diagnostic plots.

At each monitor, we take the simple difference of the observed PM_2.5_ concentrations during the lockdown and the SCM-estimated counterfactual PM_2.5_ concentrations to obtain the lockdown-attributable changes in PM_2.5_ for each day. We then average these daily effect estimates over the entire lockdown period to obtain the ATT estimate at each monitor, which is shown in Figure 1.

In the second stage of modeling, we use a linear regression model to identify features associated with the estimated lockdown-attributable changes in PM_2.5_. The effect of meteorological variables adjusted for in the first stage of modeling is first subtracted from the treatment effect estimates. Each monitor is linked to a large set of features of the county it resides in. The units of analysis in this model are monitors, and the outcome in the regression model is the monitor-level estimated lockdown-attributable change in PM_2.5_. Features included as predictors in the model are: residential emissions, industrial processes emissions, industrial boiler emissions, dust emissions, agriculture emissions, mobile emissions, mobility change relative to baseline, socioeconomic and demographic variables (proportion of population age 65+, racial composition, poverty rate, population density), urbanization level (classified into large central metro, large fringe metro, medium metro, and other), and US census-defined regions (Midwest, Northwest, South, and West). All features are obtained at the county-level. Descriptions of each variable and data sources are provided in Extended Data Table E2.

Mobility change was measured relative to a February 2020 baseline and defined by quantifying the number of Bing tiles Facebook users are seen in during a day^27^. Socioeconomic and demographic variables were taken from the 5-year 2018 American Community Survey^19^ and meteorological variables were obtained from the Google Earth Engine^26^. Data on sources of emissions were obtained from the EPA 2017 National Emissions Inventory reports^18,22^. Urban-rural classifications are obtained from 2013 National Center for Health Statistics Urban-Rural Classification Scheme for Counties^21^ and large central metro is used as the reference variable in the model. Areas classified as smaller than “medium metro” are grouped into a variable called “other”. The Midwest Region is the reference variable in the model for region indicators. Coefficient estimates and respective confidence intervals for the model including interaction effects between relative mobility change and population age 65+, are provided in Extended Data Table E1. However, the addition of these interaction effects did not significantly affect the model coefficients.

We use exposure-response functions from the EPA’s Core Health Impact Functions for Particulate Matter and Hospital Admissions^23^ to characterize associations between short-term PM_2.5_ exposure and (1) respiratory disease hospitalization risk and (2) cardiovascular disease hospitalization risk^28,29^,. To estimate the health impacts across the entire continental US, we first interpolate the estimated lockdown-attributable PM_2.5_ changes to obtain an estimate for each county (including those without an included monitor). Inverse distance weighting is used to interpolate the estimated effects at the monitor sites to each county’s centroid, and the resulting interpolated values are treated as each county’s lockdown-attributable change in PM_2.5_. We insert each county’s PM_2.5_ changes into the pre-existing short-term exposure-response functions for each hospitalization type for individuals 65+ (a log-linear model with parameter estimates taken from Kloog et al.^28^ and Bell et al.^29^) to obtain an estimate of its change in hospitalization incidence rate. Baseline incidence rates are calculated using a weighted average of hospitalization incidence rates for people age 65+ (3.352 respiratory hospitalizations/100 people per year; 5.385 cardiovascular hospitalizations/100 people per year). The change in incidence rate is then used to obtain the county’s absolute change in hospitalizations for each of the two health outcomes in people age 65+. These values are summed across each US census region and scaled to account for the number of days in the state’s lockdown period, i.e., the number of days of the specified change in PM_2.5_ exposure. We also estimate the changes in hospitalization incidence that would have occurred if these PM_2.5_ changes had been sustained for an entire year.

## Data Availability

Requests for materials should be addressed to Rachel Nethery at.

## Acknowledgements

The authors gratefully acknowledge funding from NIH grants 5T32ES007142, 1K01ES032458-01, and R01AG060232.

## Author contributions

KLC led all data preparation, conducted all statistical analyses, and prepared a first draft of the manuscript. LRFH provided expertise in air pollution and atmospheric chemistry and assisted with manuscript preparation. RCN assisted with idea generation and study design, aided in data acquisition and preparation, provided statistical input, and assisted with manuscript preparation.

## Competing interests

The authors declare no competing interests.

## Additional information

Supplementary Information is available for this paper. Correspondence and requests for materials should be addressed to Rachel Nethery at.

## Extended Data

**Table E1.** Defined lockdown period for each contiguous US state (and the District of Columbia), beginning on the day of the declared state of emergency and ending on April 30th, 2020 or the day of business reopenings, whichever came first.

| State | State of Emergency | End Date | State | State of Emergency | End Date |
| --- | --- | --- | --- | --- | --- |
| Alabama | 2020-03-13 | 2020-04-30 | Nebraska | 2020-03-13 | 2020-04-30 |
| Arizona | 2020-03-11 | 2020-04-30 | Nevada | 2020-03-12 | 2020-04-30 |
| Arkansas | 2020-03-11 | 2020-04-30 | New Hampshire | 2020-03-13 | 2020-04-30 |
| California | 2020-03-04 | 2020-04-30 | New Jersey | 2020-03-09 | 2020-04-30 |
| Colorado | 2020-03-11 | 2020-04-30 | New Mexico | 2020-03-11 | 2020-04-30 |
| Connecticut | 2020-03-10 | 2020-04-30 | New York | 2020-03-07 | 2020-04-30 |
| Delaware | 2020-03-13 | 2020-04-30 | North Carolina | 2020-03-10 | 2020-04-30 |
| District of Columbia | 2020-03-11 | 2020-04-30 | North Dakota | 2020-03-13 | 2020-04-30 |
| Florida | 2020-03-09 | 2020-04-30 | Ohio | 2020-03-09 | 2020-04-30 |
| Georgia | 2020-03-14 | 2020-04-30 | Oklahoma | 2020-03-15 | 2020-04-24 |
| Idaho | 2020-03-13 | 2020-04-30 | Oregon | 2020-03-08 | 2020-04-30 |
| Illinois | 2020-03-09 | 2020-04-30 | Pennsylvania | 2020-03-06 | 2020-04-30 |
| Indiana | 2020-03-06 | 2020-04-30 | Rhode Island | 2020-03-09 | 2020-04-30 |
| Iowa | 2020-03-09 | 2020-04-30 | South Carolina | 2020-03-13 | 2020-04-20 |
| Kansas | 2020-03-12 | 2020-04-30 | South Dakota | 2020-03-13 | 2020-04-28 |
| Kentucky | 2020-03-06 | 2020-04-30 | Tennessee | 2020-03-12 | 2020-04-30 |
| Louisiana | 2020-03-11 | 2020-04-30 | Texas | 2020-03-13 | 2020-04-30 |
| Maine | 2020-03-15 | 2020-04-30 | Utah | 2020-03-06 | 2020-04-30 |
| Maryland | 2020-03-05 | 2020-04-30 | Vermont | 2020-03-13 | 2020-04-27 |
| Massachusetts | 2020-03-10 | 2020-04-30 | Virginia | 2020-03-12 | 2020-04-30 |
| Michigan | 2020-03-10 | 2020-04-30 | Washington | 2020-02-29 | 2020-04-30 |
| Minnesota | 2020-03-13 | 2020-04-27 | West Virginia | 2020-03-16 | 2020-04-30 |
| Mississippi | 2020-03-14 | 2020-04-27 | Wisconsin | 2020-03-12 | 2020-04-30 |
| Missouri | 2020-03-13 | 2020-04-30 | Wyoming | 2020-03-13 | 2020-04-30 |
| Montana | 2020-03-12 | 2020-04-27 |  |  |  |

**Table E2.** List of variables included in regression model, with units of measure and abbreviations used throughout.

| Variable | Unit of Measure | Abbreviations Used | Reference |
| --- | --- | --- | --- |
| Primary PM<sub>2.5</sub> emissions from agricultural sources | $\mu g/m^3$ | "Agriculture Emissions" | US EPA<sup>18</sup> |
| Primary PM<sub>2.5</sub> emissions from dust sources | $\mu g/m^3$ | "Dust Emissions" | US EPA<sup>18</sup> |
| Primary PM<sub>2.5</sub> emissions from industrial boilers | $\mu g/m^3$ | "Industrial Boiler Emissions" | US EPA<sup>18</sup> |
| Primary PM<sub>2.5</sub> emissions from industrial processes | $\mu g/m^3$ | "Industrial Processes Emissions" | US EPA<sup>18</sup> |
| Primary PM<sub>2.5</sub> emissions from residential sources | $\mu g/m^3$ | "Residential Emissions" | US EPA<sup>18</sup> |
| Primary PM<sub>2.5</sub> emissions from mobile sources | $\mu g/m^3$ | "Mobile Emissions" | US EPA<sup>18</sup> |
| Mobility change rel. to baseline | % | "Relative Mobility Change" | Facebook<sup>20</sup> |
| Proportion of population age 65+ | % | "% Age 65+" | US Census Bureau<sup>19</sup> |
| Proportion of whites in population | % | "% White" | US Census Bureau<sup>19</sup> |
| Population density | people/mi.<sup>2</sup> | N/A | US Census Bureau<sup>19</sup> |
| Poverty rate | % | N/A | US Census Bureau<sup>19</sup> |
| Large fringe metro | 0/1 | N/A | Ingram et al.<sup>21</sup> |
| Medium metro | 0/1 | N/A | Ingram et al.<sup>21</sup> |
| All other areas | 0/1 | "Other" | Ingram et al.<sup>21</sup> |
| Northeast region | 0/1 | "Northeast" |  |
| South region | 0/1 | "South" |  |
| West region | 0/1 | "West" |  |
| Maximum temperature | *K* | "Max Temp" | Gorelick et al.<sup>26</sup> |
| Maximum relative humidity | % | "Relative Humidity" | Gorelick et al.<sup>26</sup> |
| Cumulative precipitation | *mm* | "Precipitation" | Gorelick et al.<sup>26</sup> |
| Wind speed | *m/s* | N/A | Gorelick et al.<sup>26</sup> |
| Wind direction | Deg. from north | N/A | Gorelick et al.<sup>26</sup> |

**Figure E1.**
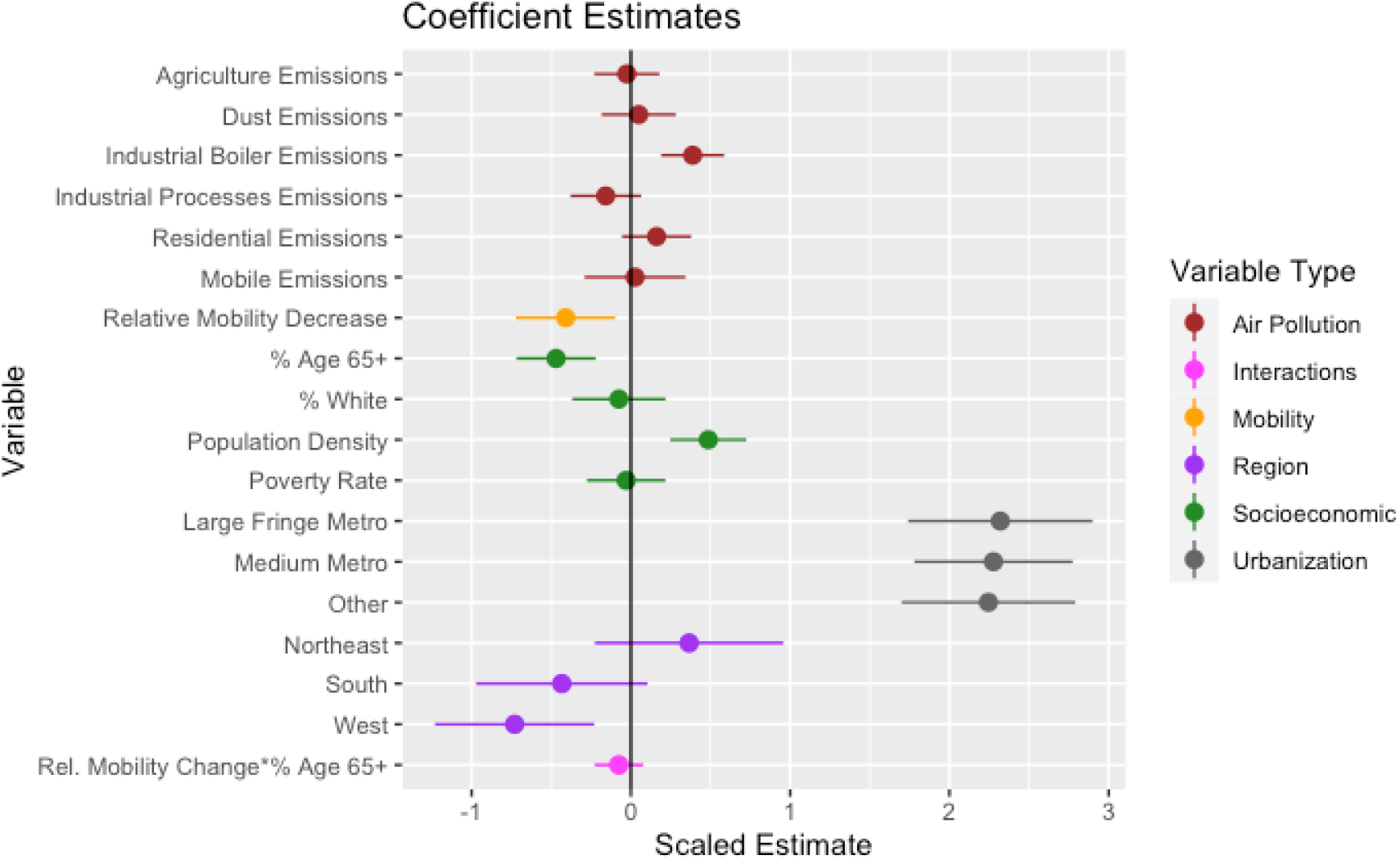
Coefficient estimates and 95% confidence intervals including interaction effects.

**Figure E2.**
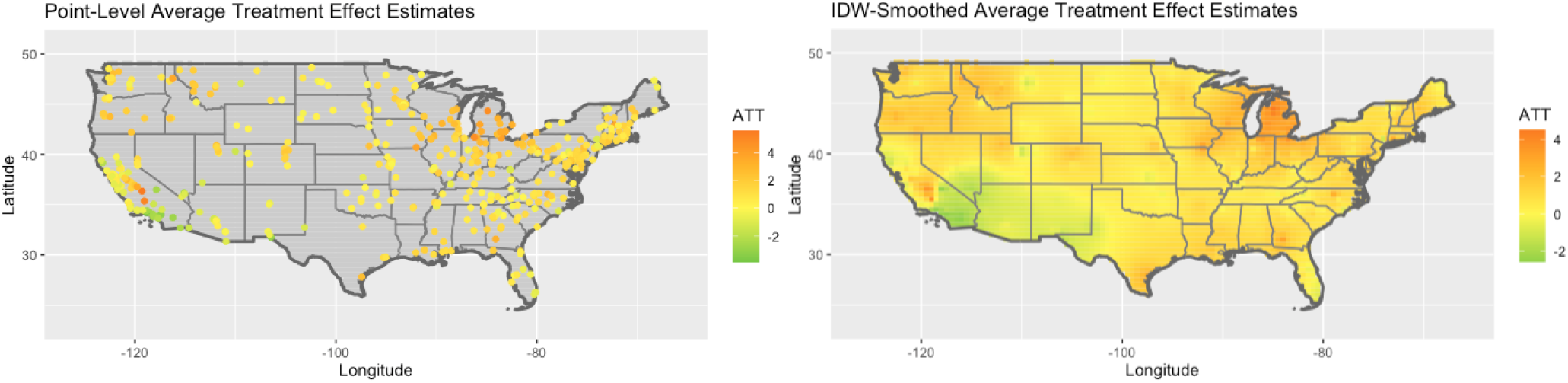
(Left) Monitor-level estimated ATTs, in *µ*g/m^3^, after removal of 1-2 day spikes of unknown origin. (Right) ATTs (in *µ*g/m^3^) smoothed across the US using inverse distance weighting.

## Notes

### Competing Interest Statement

The authors have declared no competing interest.

