## Supplementary Figure 1 for "Differential Impacts of COVID-19 Lockdowns on PM_2.5_ across the United States"

### Supplemental Information

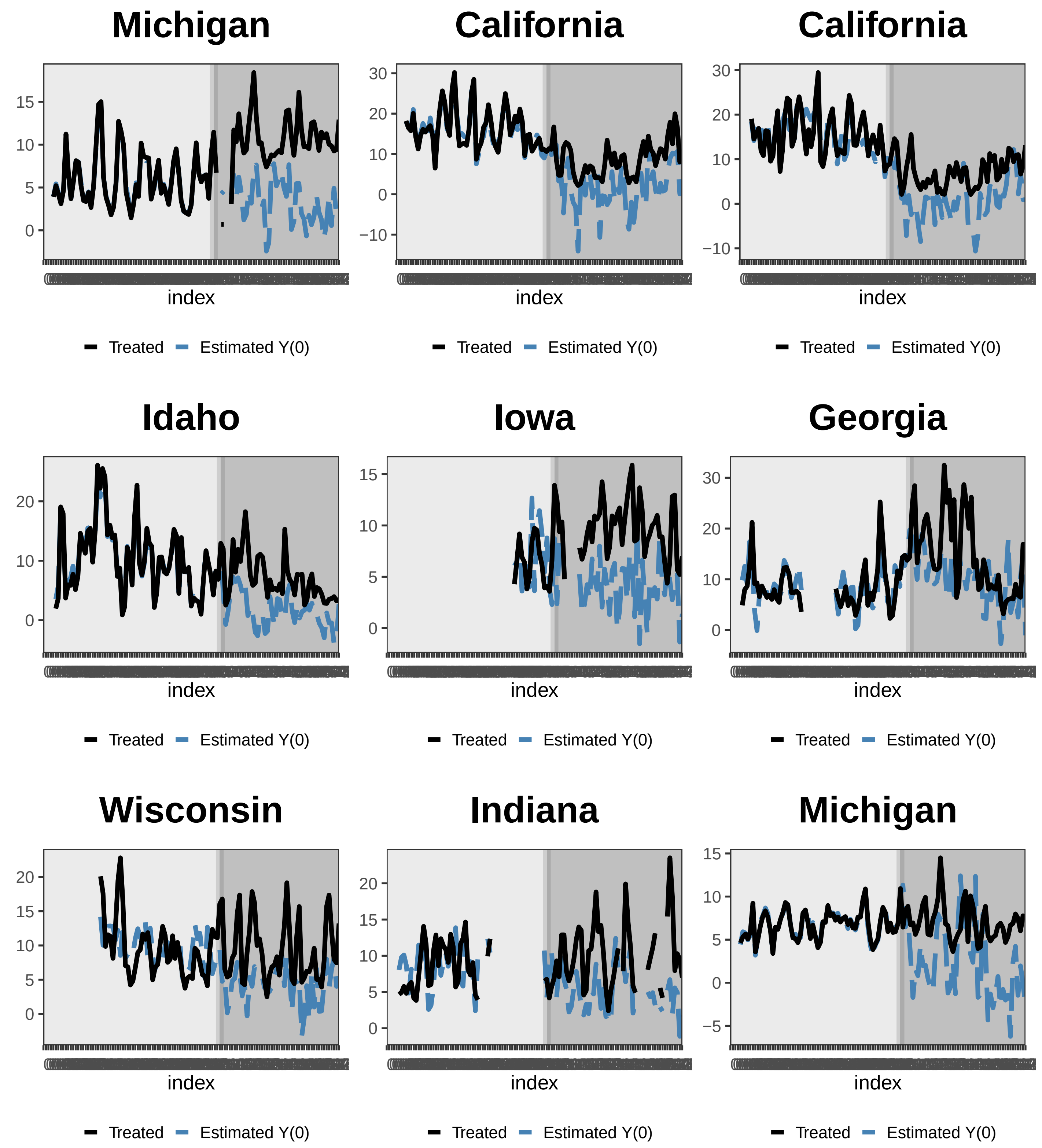
Michigan
15
10
5
0
index
Treated
Estimated Y(0)
California
30
20
10
0
−10
index
Treated
Estimated Y(0)
California
30
20
10
0
−10
index
Treated
Estimated Y(0)
Idaho
20
10
0
index
Treated
Estimated Y(0)
Iowa
15
10
5
0
index
Treated
Estimated Y(0)
Georgia
30
20
10
0
index
Treated
Estimated Y(0)
Wisconsin
20
15
10
5
0
index
Treated
Estimated Y(0)
Indiana
20
15
10
5
0
index
Treated
Estimated Y(0)
Michigan
15
10
5
0
−5
index
Treated
Estimated Y(0)

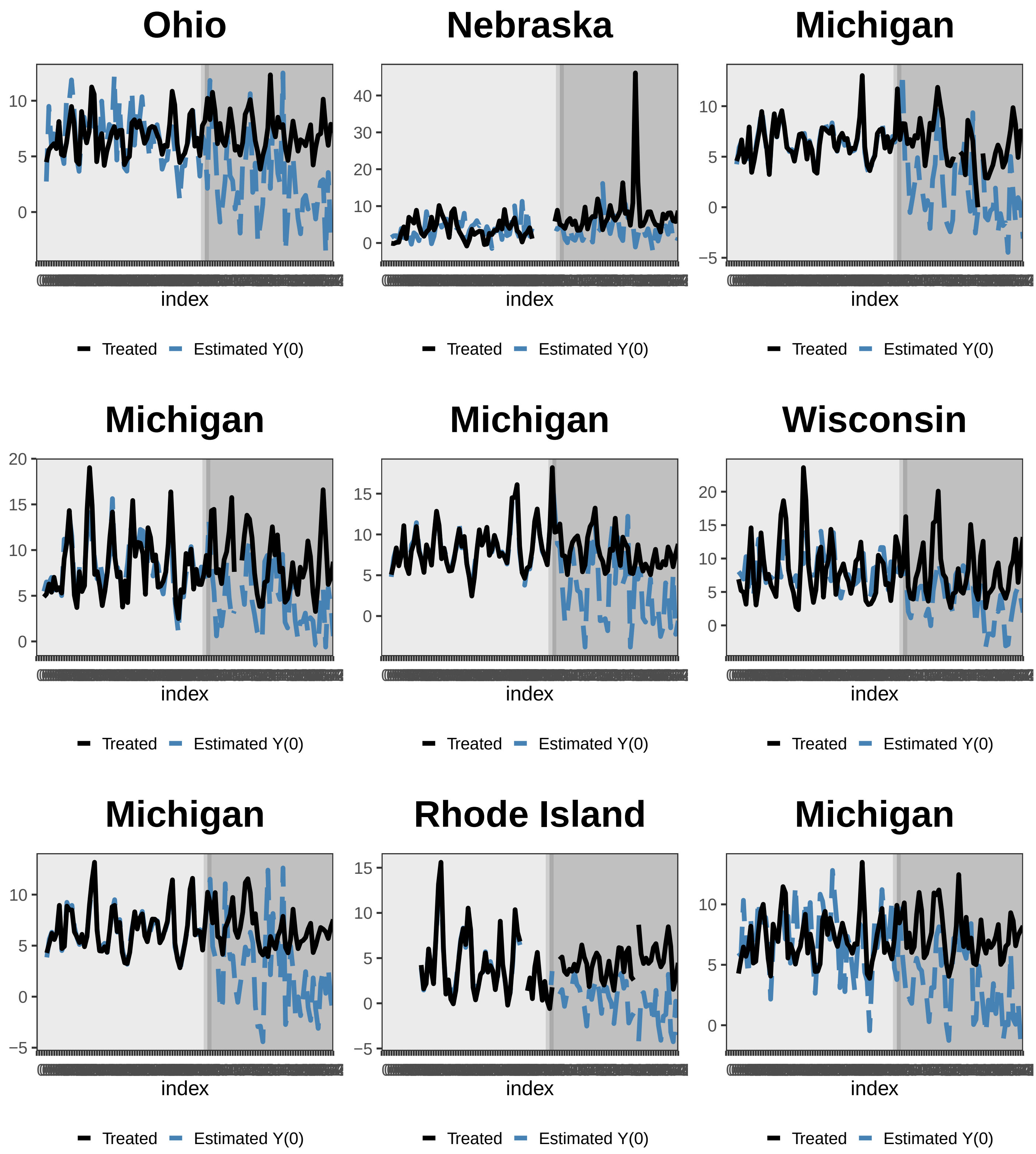
Ohio
index
Treated
Estimated Y(0)
Nebraska
index
Treated
Estimated Y(0)
Michigan
index
Treated
Estimated Y(0)
Michigan
index
Treated
Estimated Y(0)
Michigan
index
Treated
Estimated Y(0)
Wisconsin
index
Treated
Estimated Y(0)
Michigan
index
Treated
Estimated Y(0)
Rhode Island
index
Treated
Estimated Y(0)
Michigan
index
Treated
Estimated Y(0)

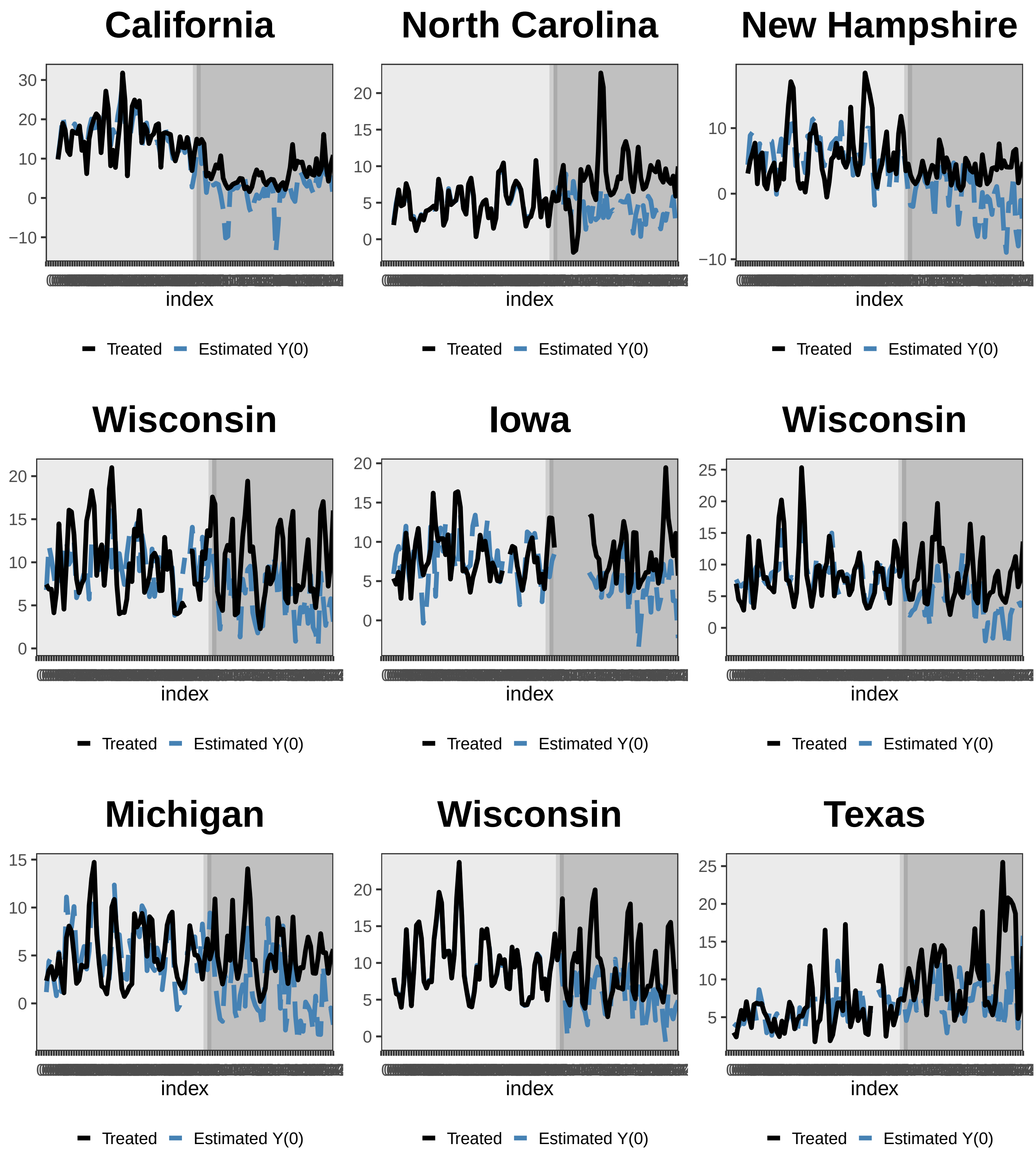
California
30
20
10
0
−10
index
Treated
Estimated Y(0)
North Carolina
20
15
10
5
0
index
Treated
Estimated Y(0)
New Hampshire
10
0
−10
index
Treated
Estimated Y(0)
Wisconsin
20
15
10
5
0
index
Treated
Estimated Y(0)
Iowa
20
15
10
5
0
index
Treated
Estimated Y(0)
Wisconsin
25
20
15
10
5
0
index
Treated
Estimated Y(0)
Michigan
15
10
5
0
index
Treated
Estimated Y(0)
Wisconsin
20
15
10
5
0
index
Treated
Estimated Y(0)
Texas
25
20
15
10
5
index
Treated
Estimated Y(0)

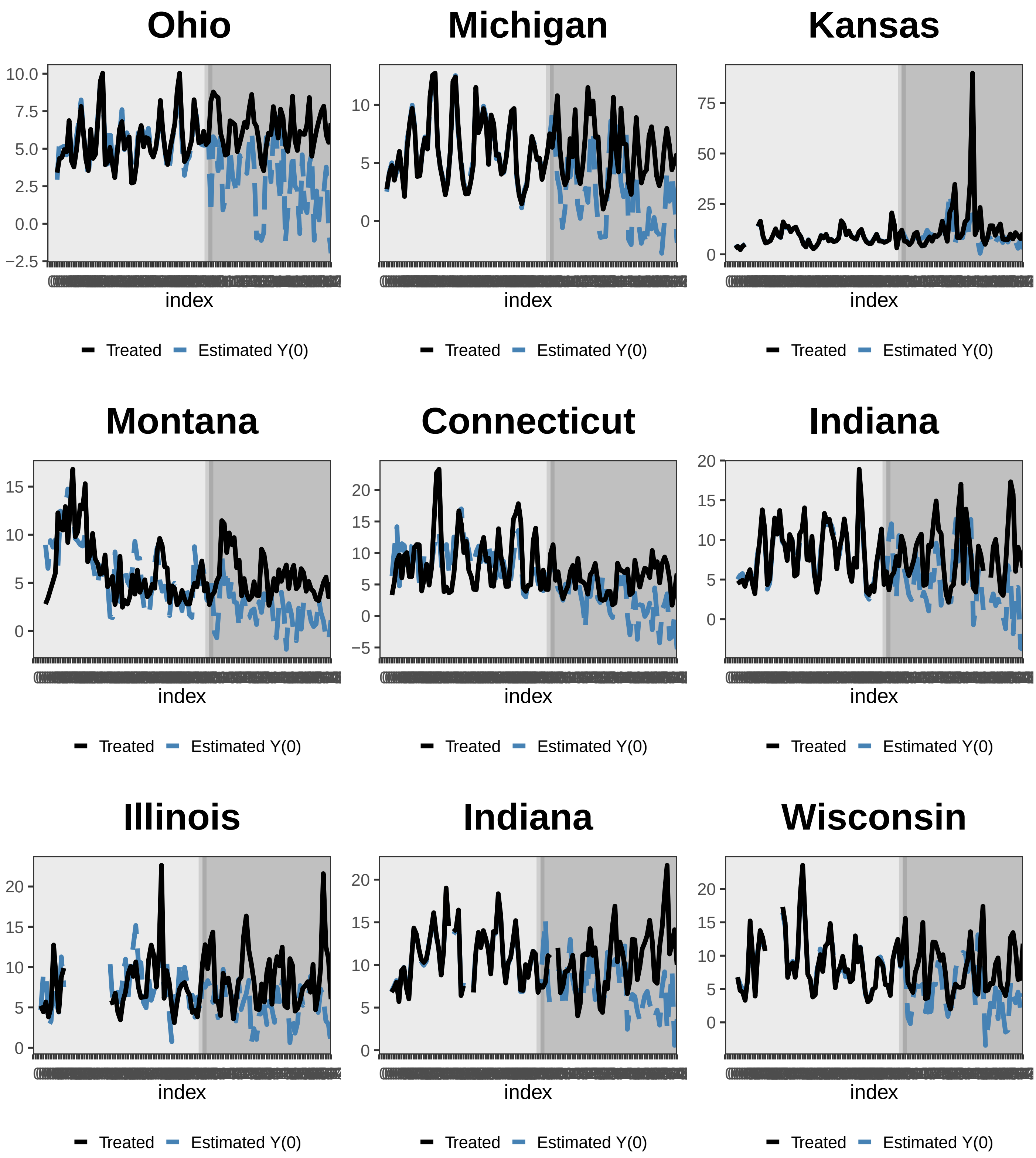
Ohio
10.0
7.5
5.0
2.5
0.0
−2.5
index
Treated
Estimated Y(0)
Michigan
10
5
0
index
Treated
Estimated Y(0)
Kansas
75
50
25
0
index
Treated
Estimated Y(0)
Montana
15
10
5
0
index
Treated
Estimated Y(0)
Connecticut
20
15
10
5
0
−5
index
Treated
Estimated Y(0)
Indiana
20
15
10
5
0
index
Treated
Estimated Y(0)
Illinois
20
15
10
5
0
index
Treated
Estimated Y(0)
Indiana
20
15
10
5
0
index
Treated
Estimated Y(0)
Wisconsin
20
15
10
5
0
index
Treated
Estimated Y(0)

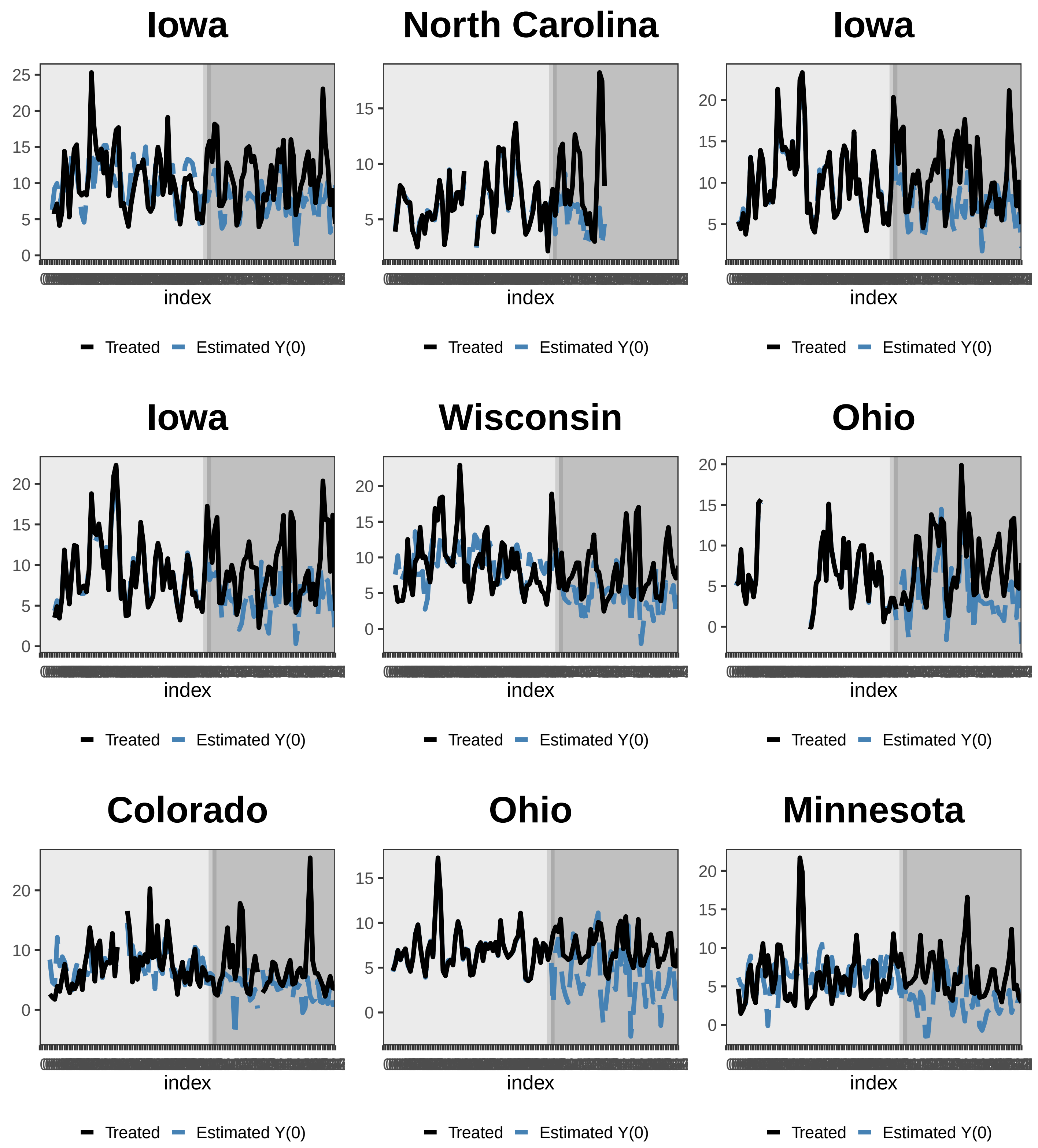
Iowa
North Carolina
Iowa
Iowa
Wisconsin
Ohio
Colorado
Ohio
Minnesota
index
Treated
Estimated Y(0)

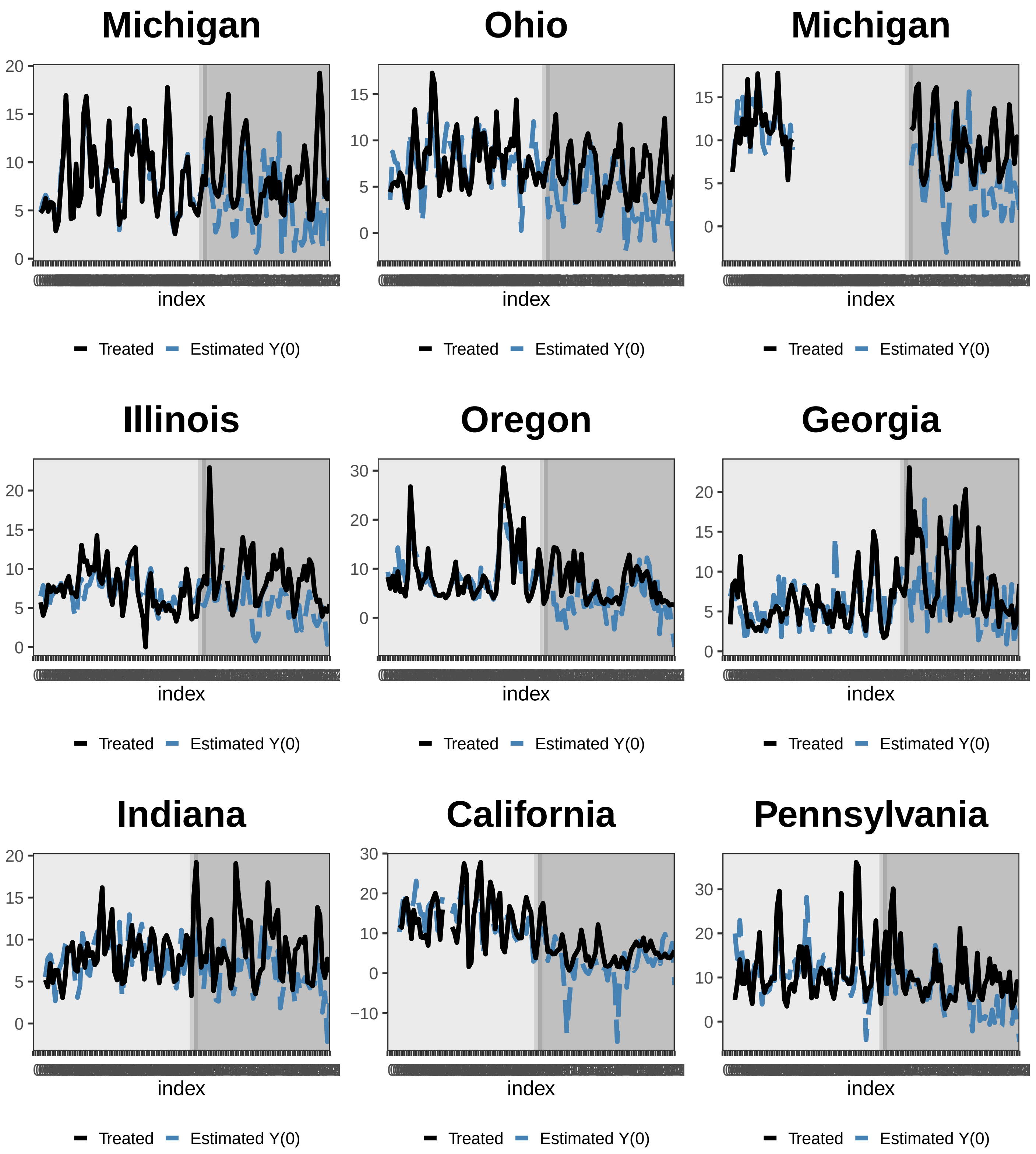
Michigan
20
15
10
5
0
index
Treated
Estimated Y(0)
Ohio
15
10
5
0
index
Treated
Estimated Y(0)
Michigan
15
10
5
0
index
Treated
Estimated Y(0)
Illinois
20
15
10
5
0
index
Treated
Estimated Y(0)
Oregon
30
20
10
0
index
Treated
Estimated Y(0)
Georgia
20
15
10
5
0
index
Treated
Estimated Y(0)
Indiana
20
15
10
5
0
index
Treated
Estimated Y(0)
California
30
20
10
0
−10
index
Treated
Estimated Y(0)
Pennsylvania
30
20
10
0
index
Treated
Estimated Y(0)

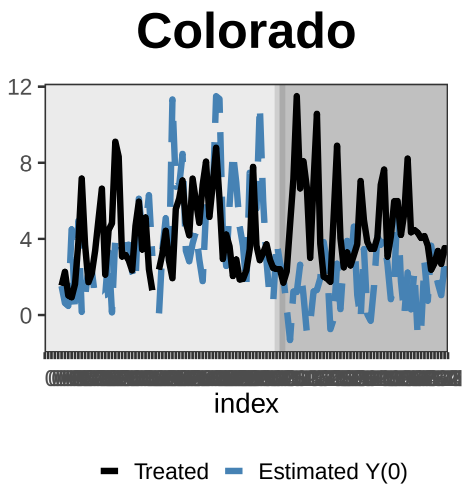
Colorado
12
8
4
0
index
Treated
Estimated Y(0)

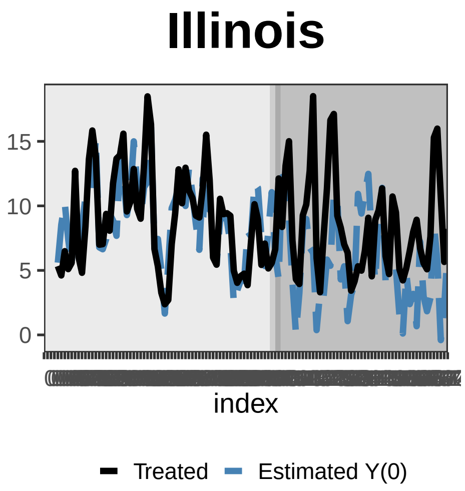
Illinois
15
10
5
0
index
Treated
Estimated Y(0)

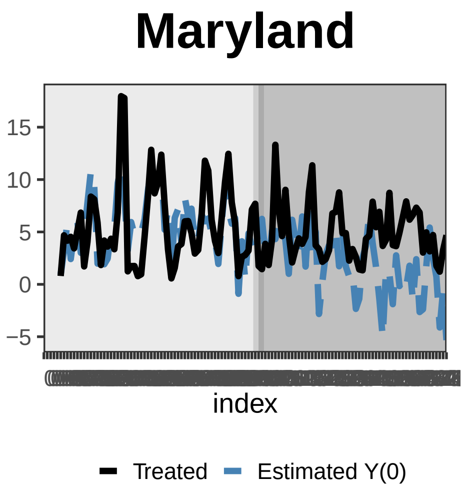
Maryland
15
10
5
0
−5
index
Treated
Estimated Y(0)

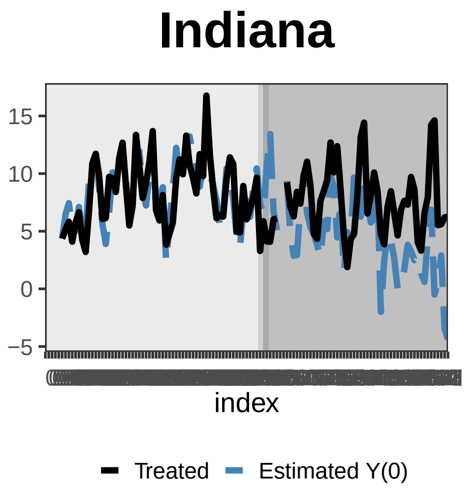
Indiana
15
10
5
0
−5
index
Treated
Estimated Y(0)

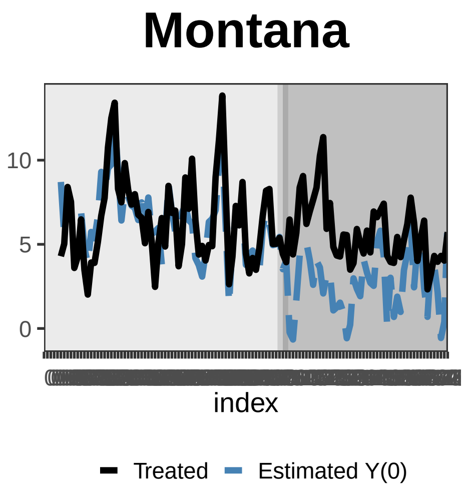
Montana
10
5
0
index
Treated
Estimated Y(0)

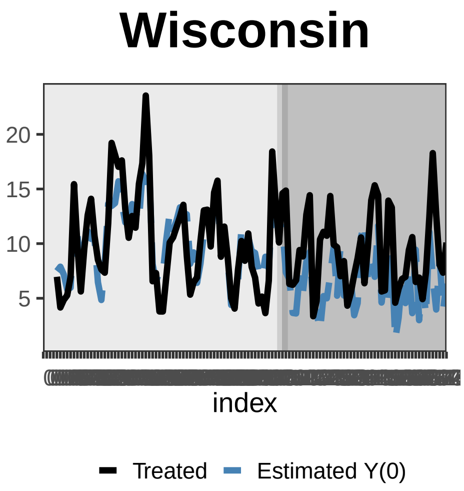
Wisconsin
20
15
10
5
index
Treated
Estimated Y(0)

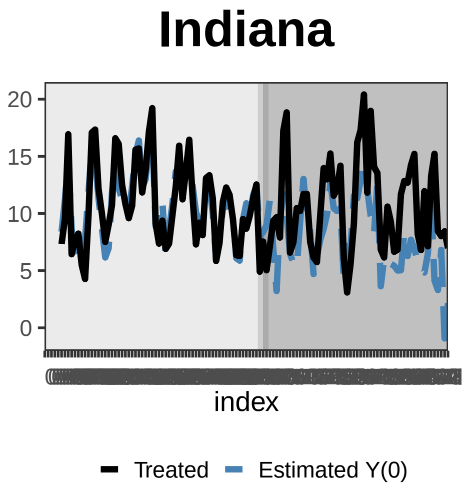
Indiana
20
15
10
5
0
index
Treated
Estimated Y(0)

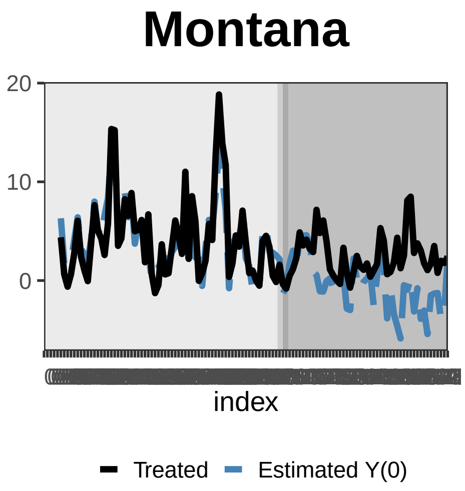
Montana
20
10
0
index
Treated
Estimated Y(0)

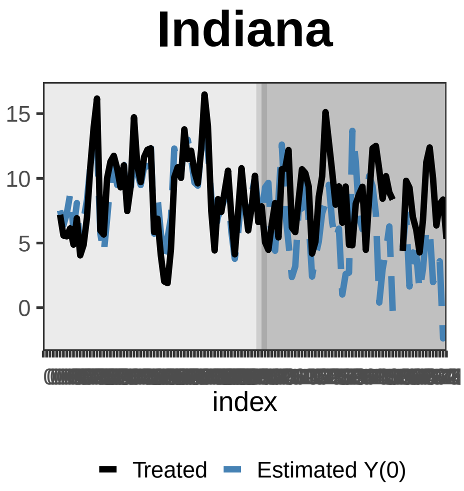
Indiana
15
10
5
0
index
Treated
Estimated Y(0)

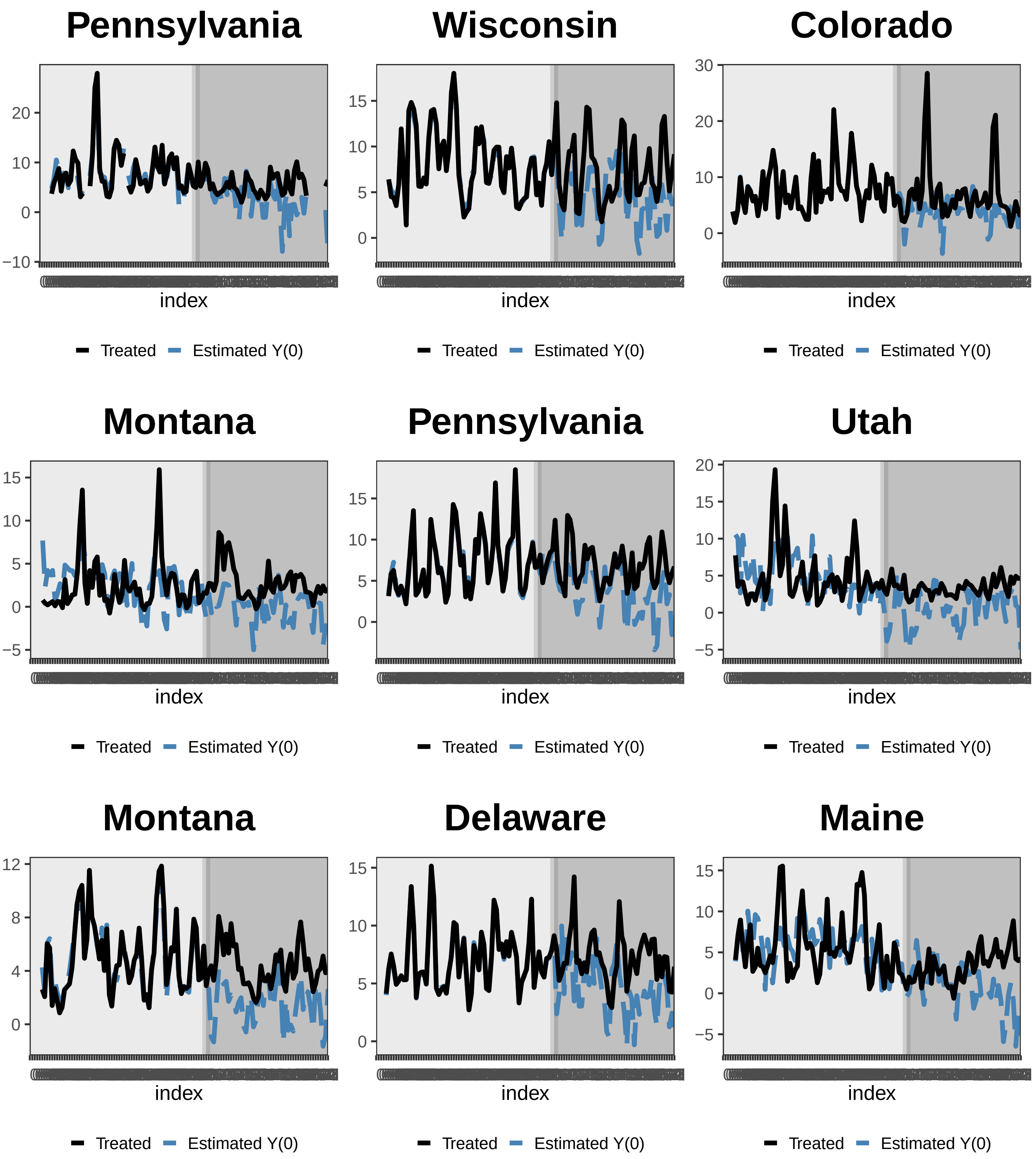
Pennsylvania
index
Treated
Estimated Y(0)
Wisconsin
index
Treated
Estimated Y(0)
Colorado
index
Treated
Estimated Y(0)
Montana
index
Treated
Estimated Y(0)
Pennsylvania
index
Treated
Estimated Y(0)
Utah
index
Treated
Estimated Y(0)
Montana
index
Treated
Estimated Y(0)
Delaware
index
Treated
Estimated Y(0)
Maine
index
Treated
Estimated Y(0)

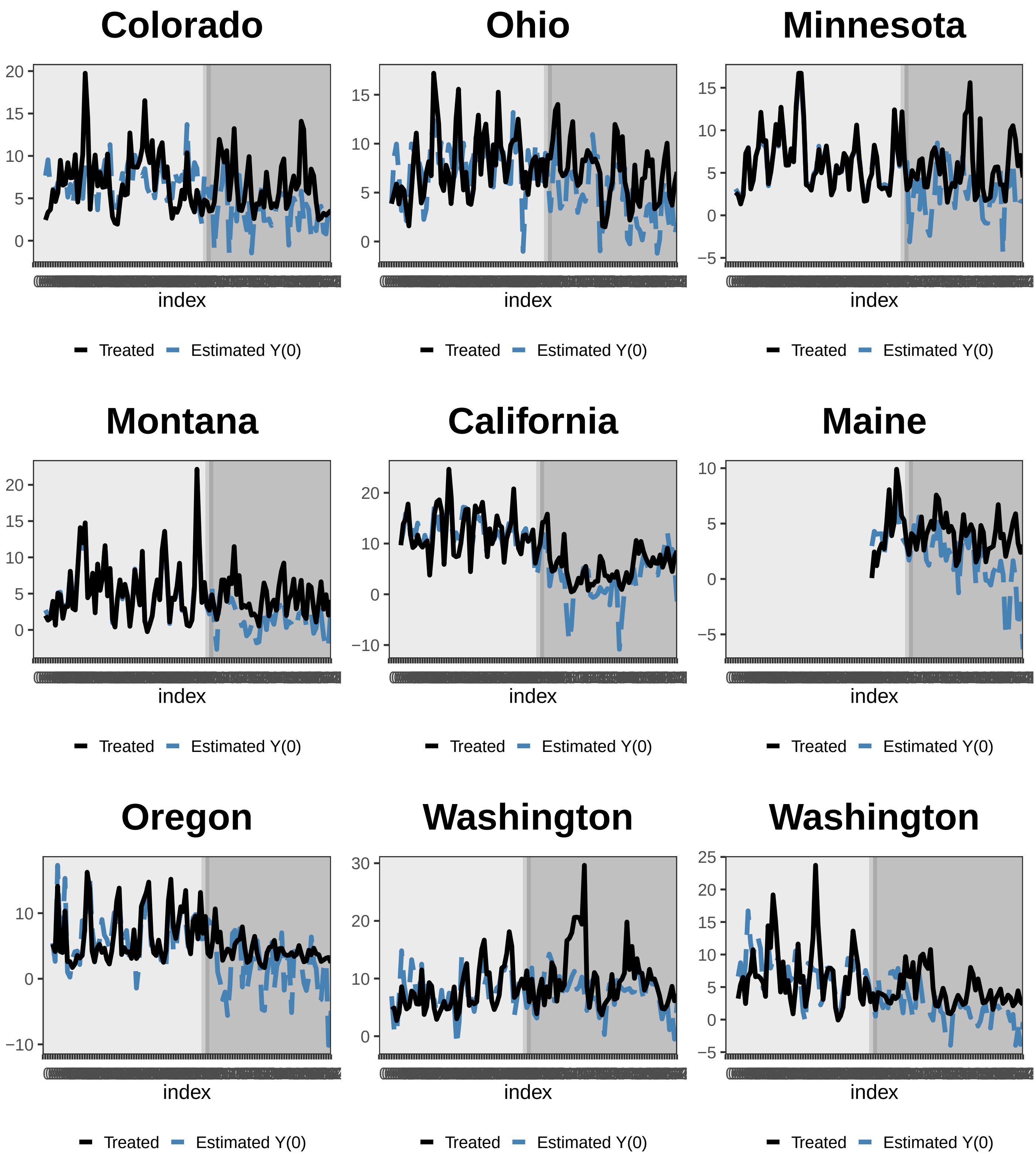
Colorado
20
15
10
5
0
index
Treated
Estimated Y(0)
Ohio
15
10
5
0
index
Treated
Estimated Y(0)
Minnesota
15
10
5
0
−5
index
Treated
Estimated Y(0)
Montana
20
15
10
5
0
index
Treated
Estimated Y(0)
California
20
10
0
−10
index
Treated
Estimated Y(0)
Maine
10
5
0
−5
index
Treated
Estimated Y(0)
Oregon
10
0
−10
index
Treated
Estimated Y(0)
Washington
30
20
10
0
index
Treated
Estimated Y(0)
Washington
25
20
15
10
5
0
−5
index
Treated
Estimated Y(0)

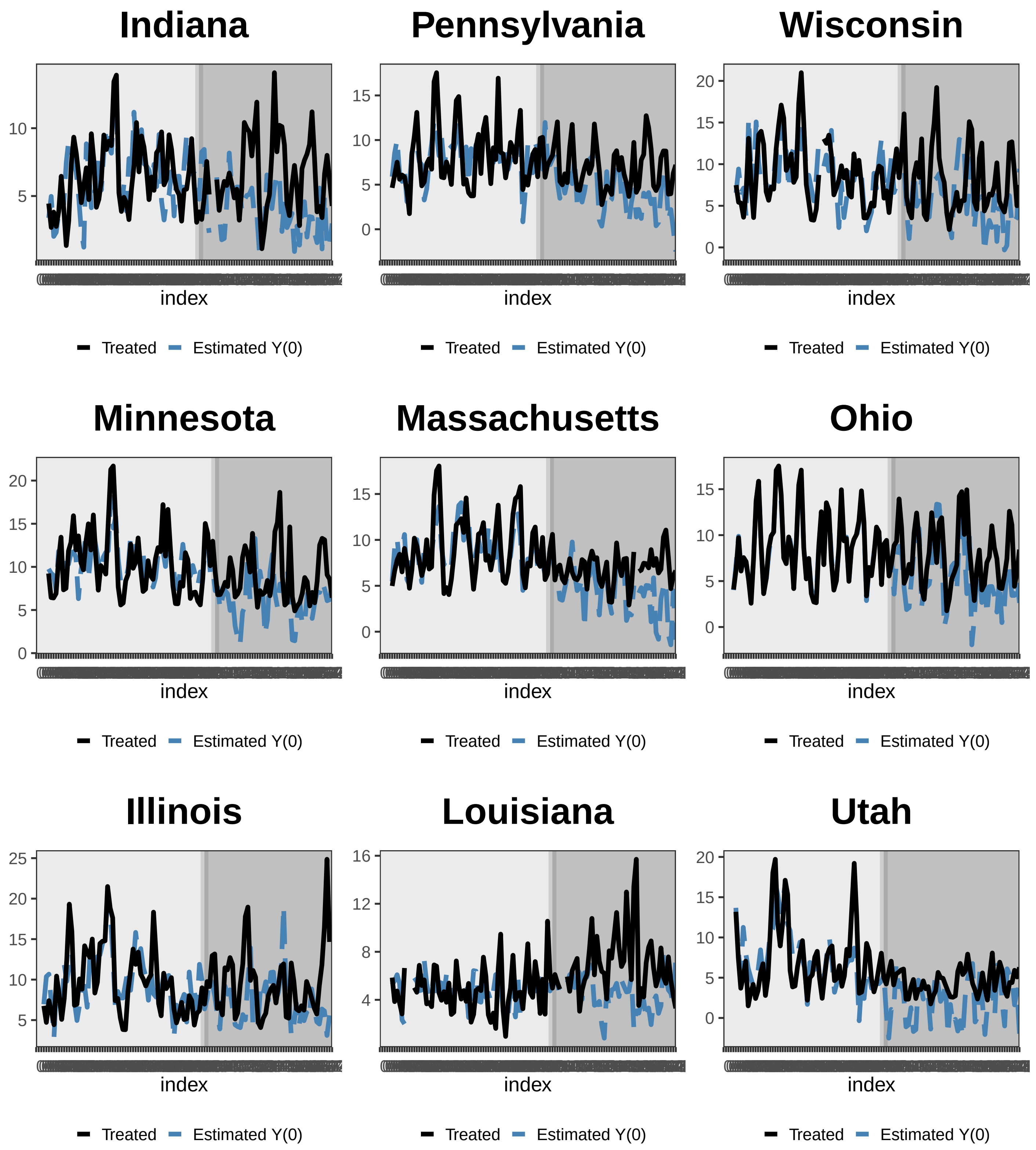
Indiana
Pennsylvania
Wisconsin
Minnesota
Massachusetts
Ohio
Illinois
Louisiana
Utah
index
Treated
Estimated Y(0)

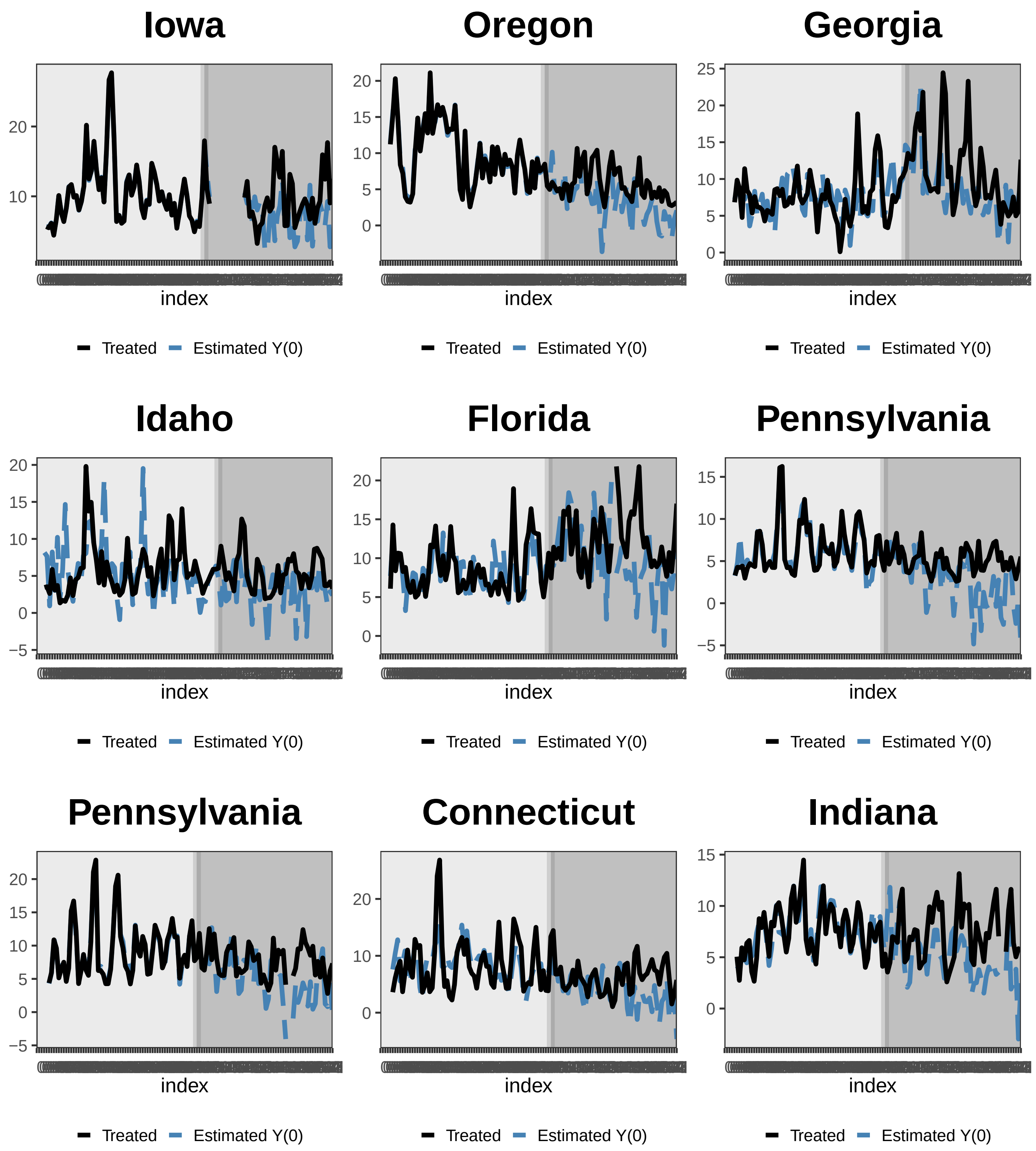
Iowa
Oregon
Georgia
Idaho
Florida
Pennsylvania
Pennsylvania
Connecticut
Indiana
index
Treated
Estimated Y(0)

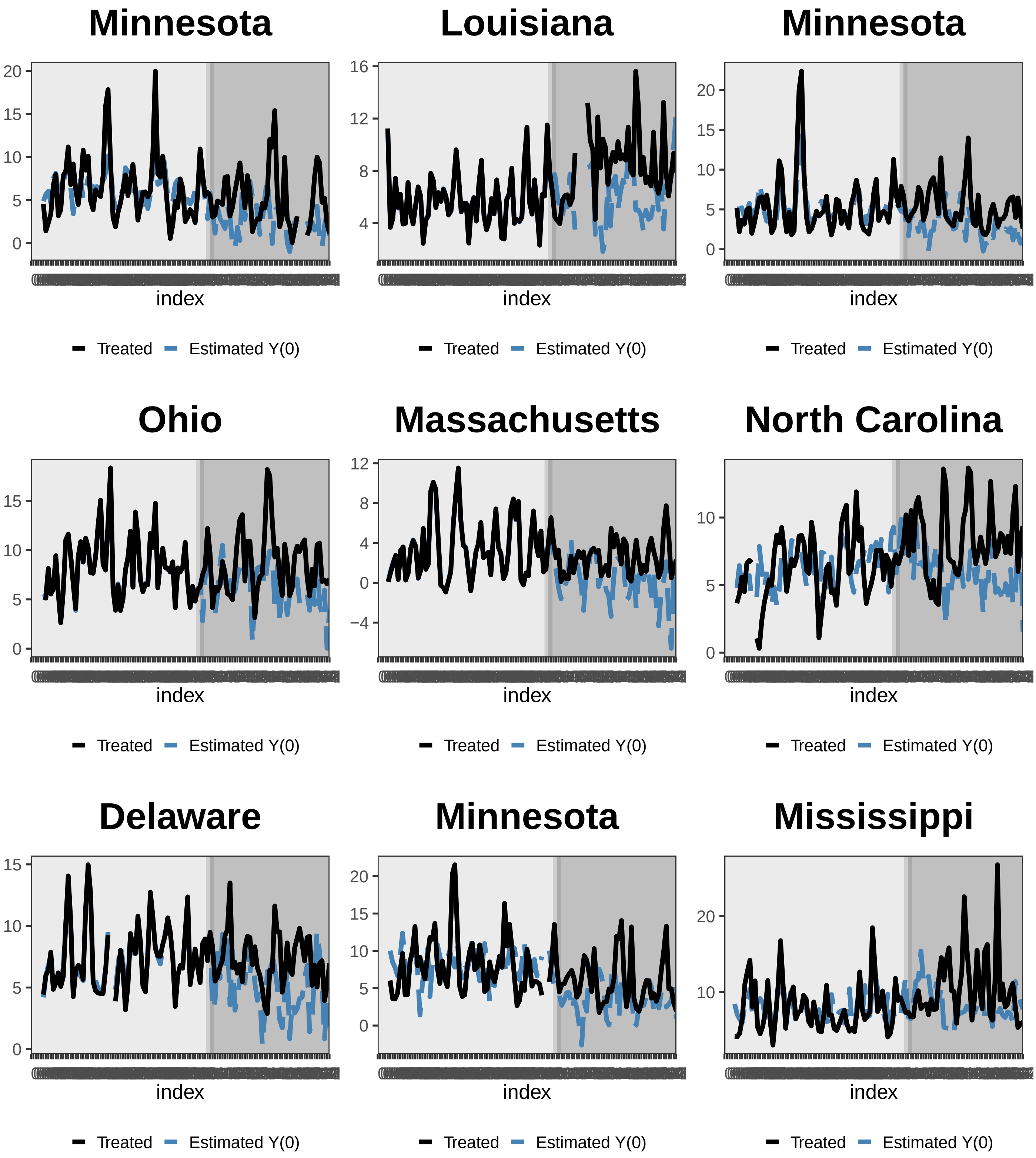
Minnesota
20
15
10
5
0
index
Treated
Estimated Y(0)
Louisiana
16
12
8
4
index
Treated
Estimated Y(0)
Minnesota
20
15
10
5
0
index
Treated
Estimated Y(0)
Ohio
15
10
5
0
index
Treated
Estimated Y(0)
Massachusetts
12
8
4
0
−4
index
Treated
Estimated Y(0)
North Carolina
10
5
0
index
Treated
Estimated Y(0)
Delaware
15
10
5
0
index
Treated
Estimated Y(0)
Minnesota
20
15
10
5
0
index
Treated
Estimated Y(0)
Mississippi
20
10
index
Treated
Estimated Y(0)

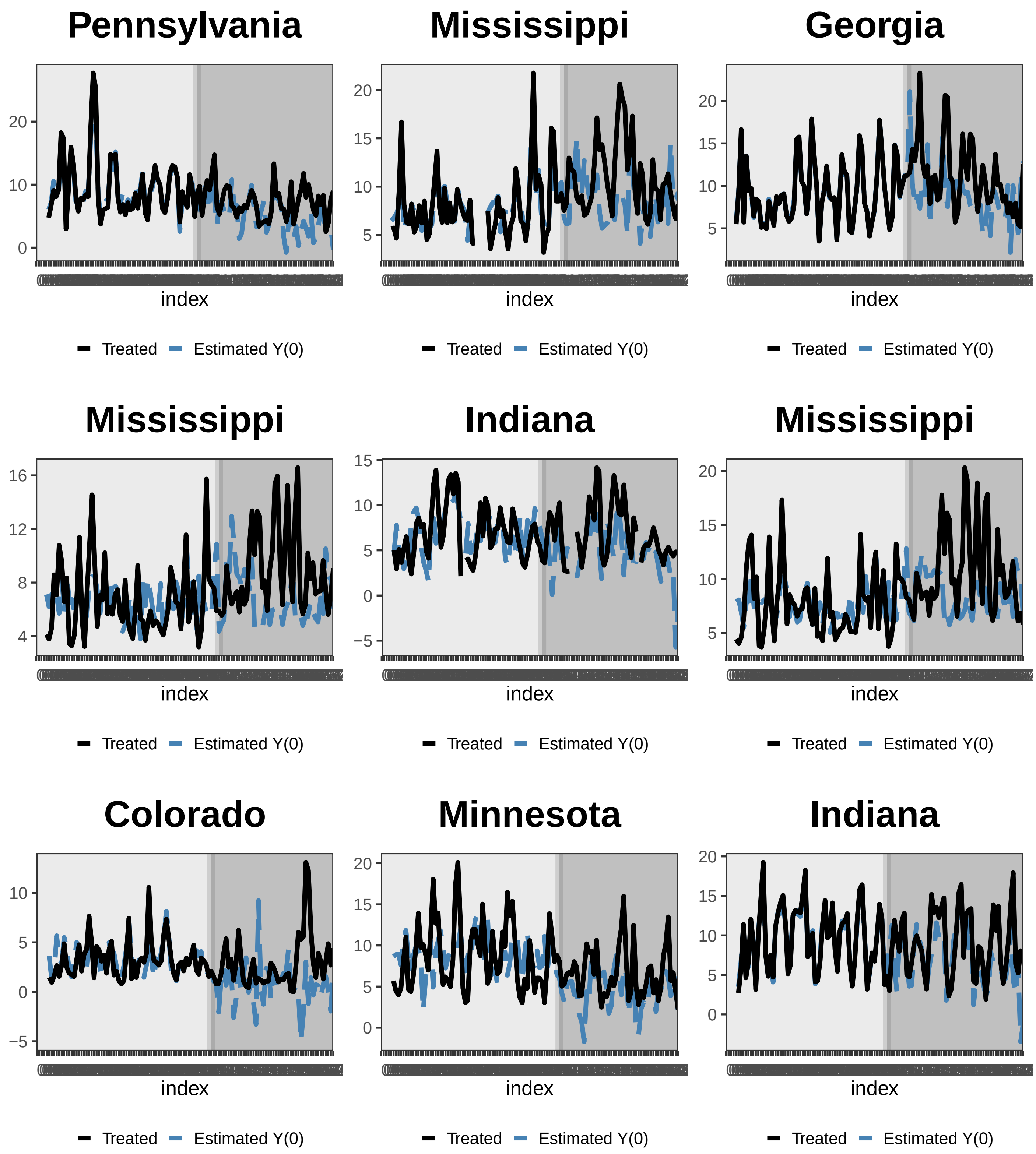
Pennsylvania
Mississippi
Georgia
Mississippi
Indiana
Mississippi
Colorado
Minnesota
Indiana
index
Treated
Estimated Y(0)

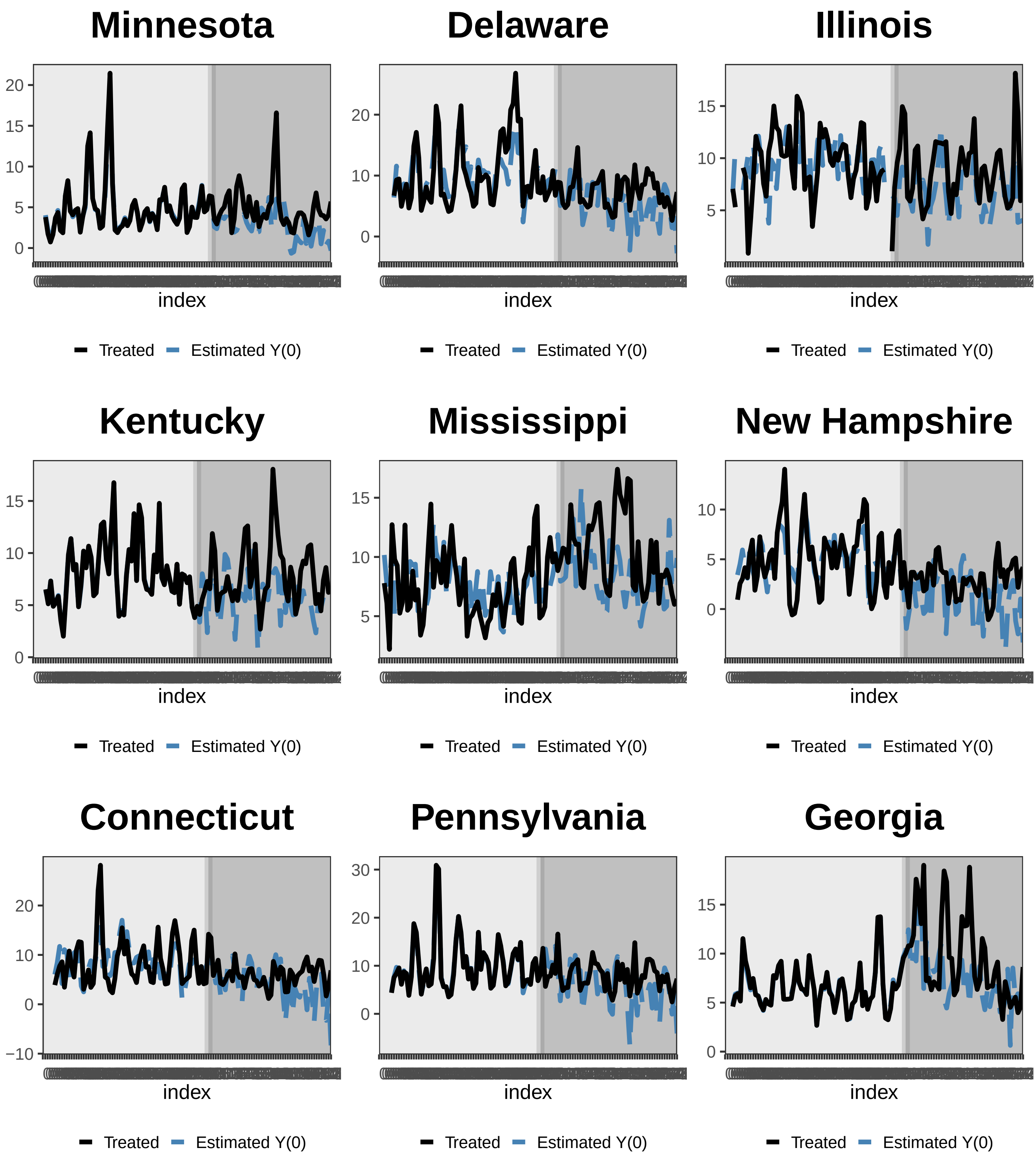
Minnesota
Delaware
Illinois
Kentucky
Mississippi
New Hampshire
Connecticut
Pennsylvania
Georgia
index
Treated
Estimated Y(0)

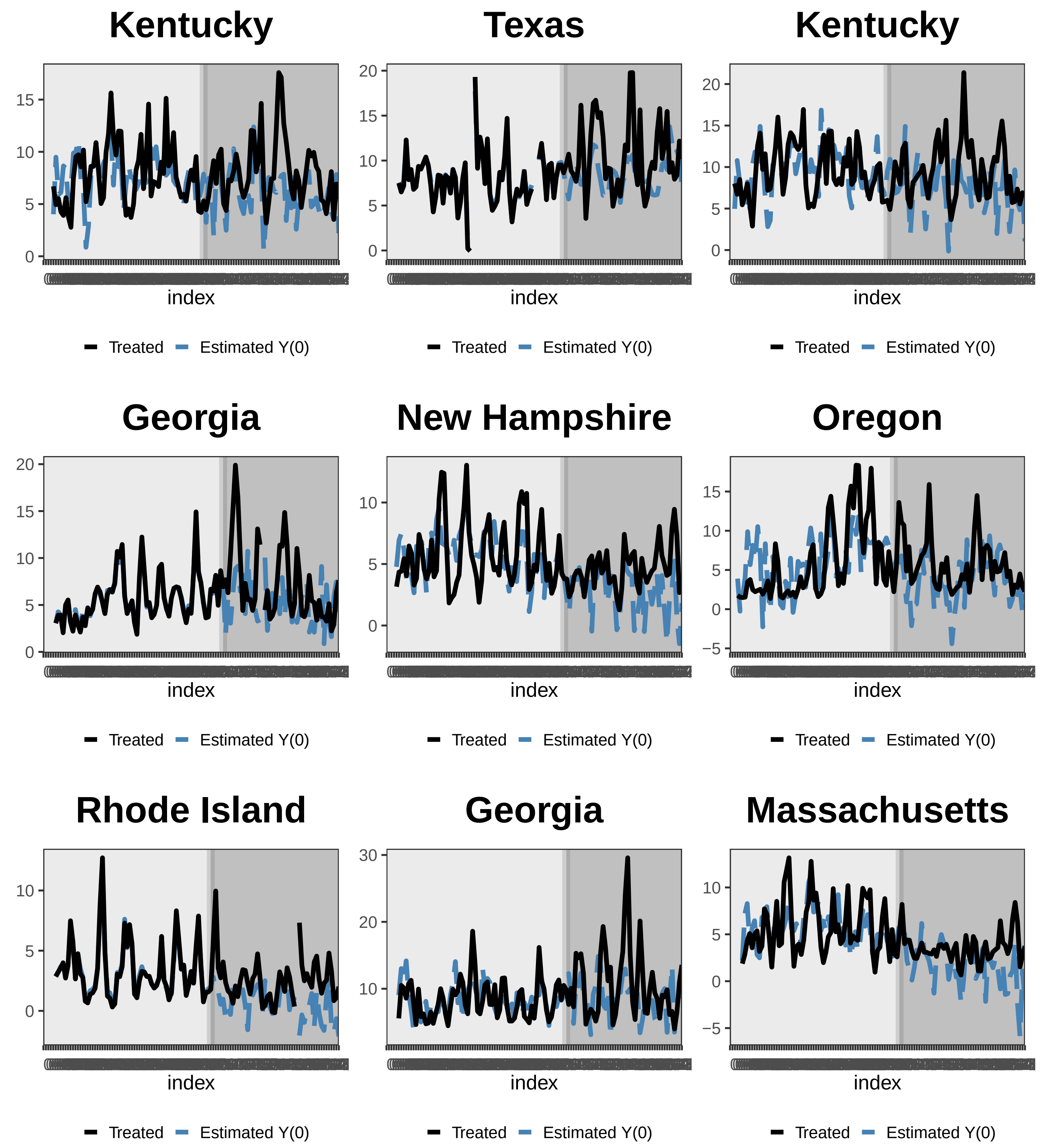
Kentucky
Texas
Kentucky
Georgia
New Hampshire
Oregon
Rhode Island
Georgia
Massachusetts
index
Treated
Estimated Y(0)

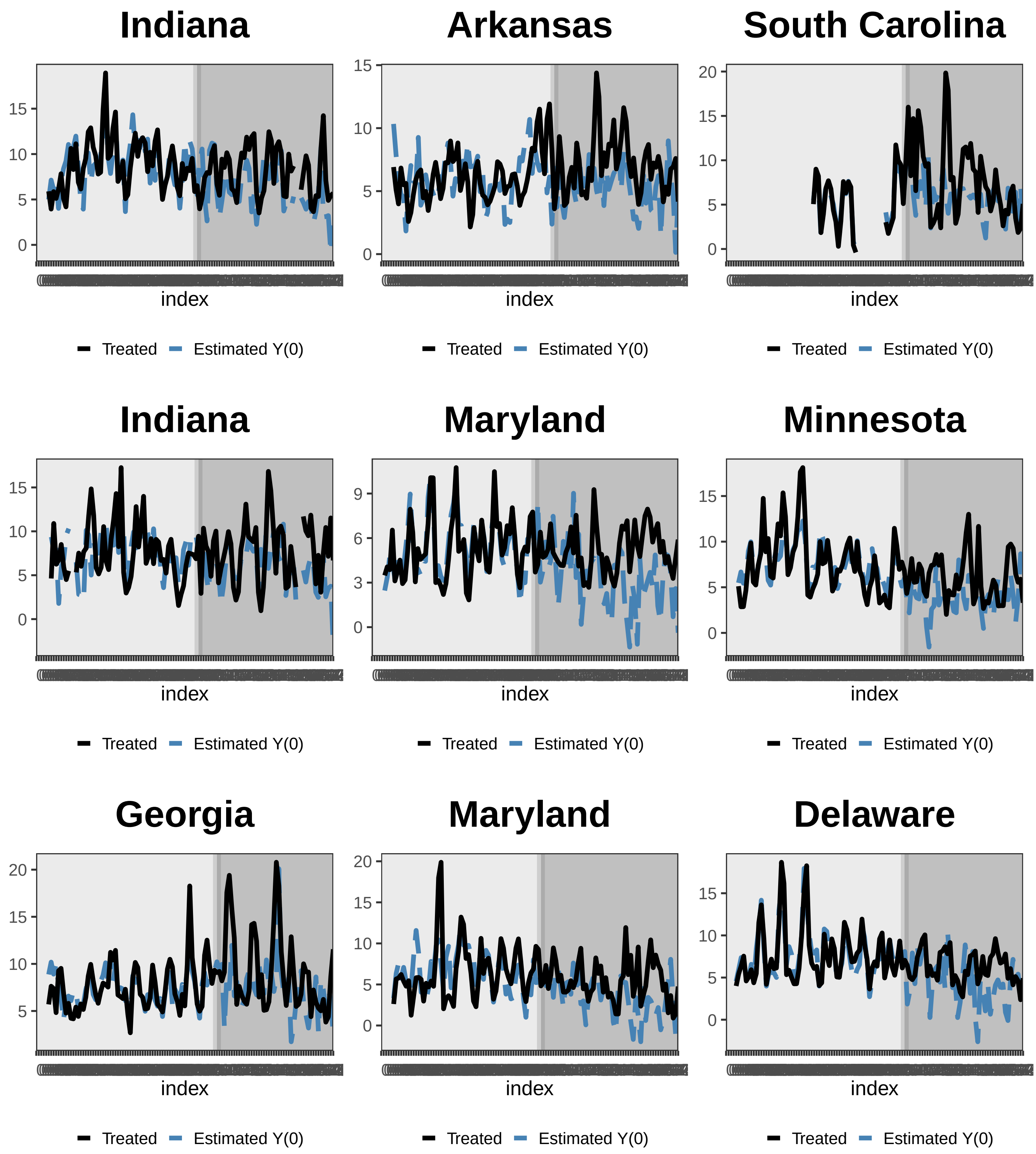
Indiana
Arkansas
South Carolina
Indiana
Maryland
Minnesota
Georgia
Maryland
Delaware
0
5
10
15
20
index
Treated
Estimated Y(0)

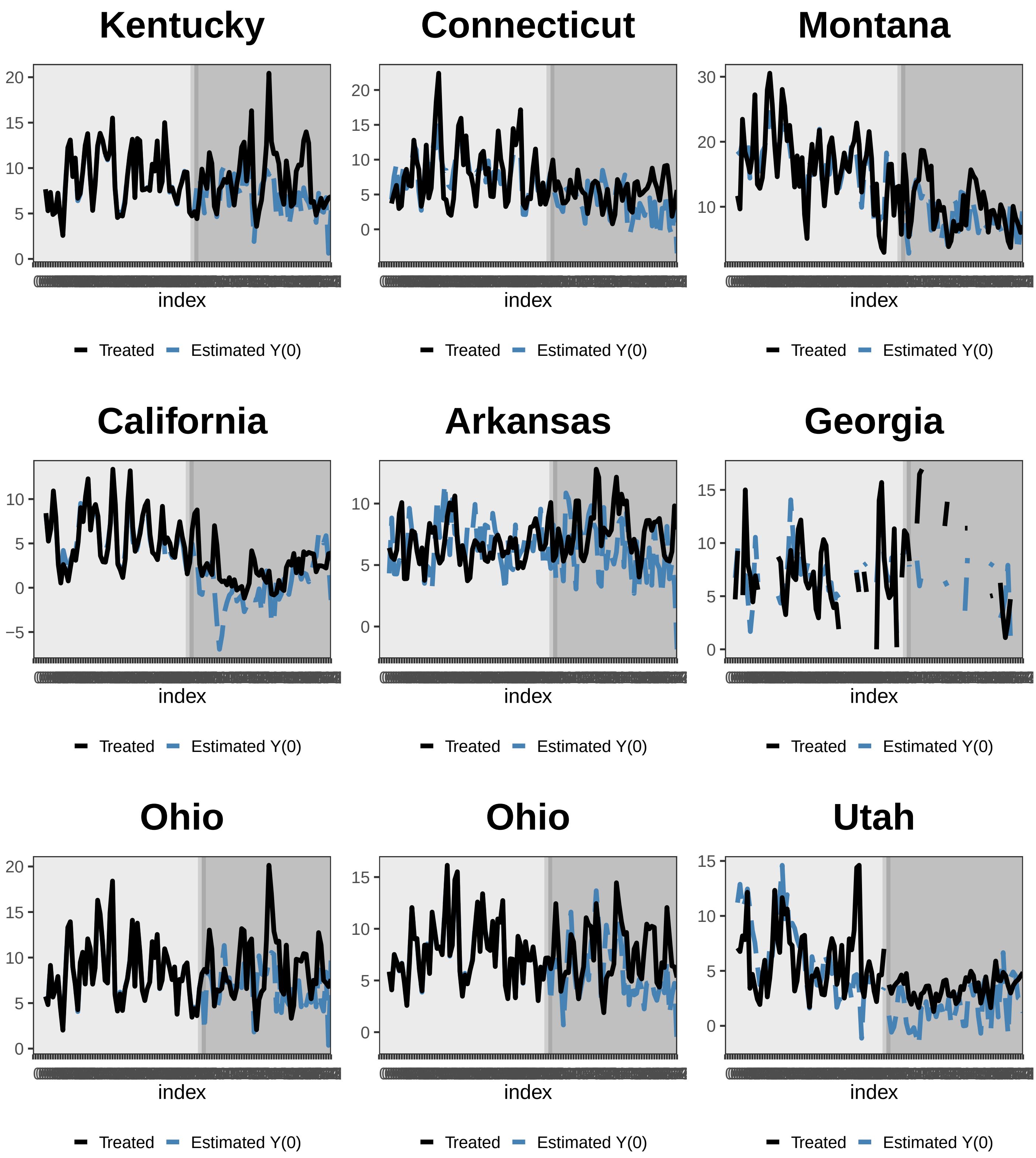
Kentucky
20
15
10
5
0
index
Treated
Estimated Y(0)
Connecticut
20
15
10
5
0
index
Treated
Estimated Y(0)
Montana
30
20
10
index
Treated
Estimated Y(0)
California
10
5
0
−5
index
Treated
Estimated Y(0)
Arkansas
10
5
0
index
Treated
Estimated Y(0)
Georgia
15
10
5
0
index
Treated
Estimated Y(0)
Ohio
20
15
10
5
0
index
Treated
Estimated Y(0)
Ohio
15
10
5
0
index
Treated
Estimated Y(0)
Utah
15
10
5
0
index
Treated
Estimated Y(0)

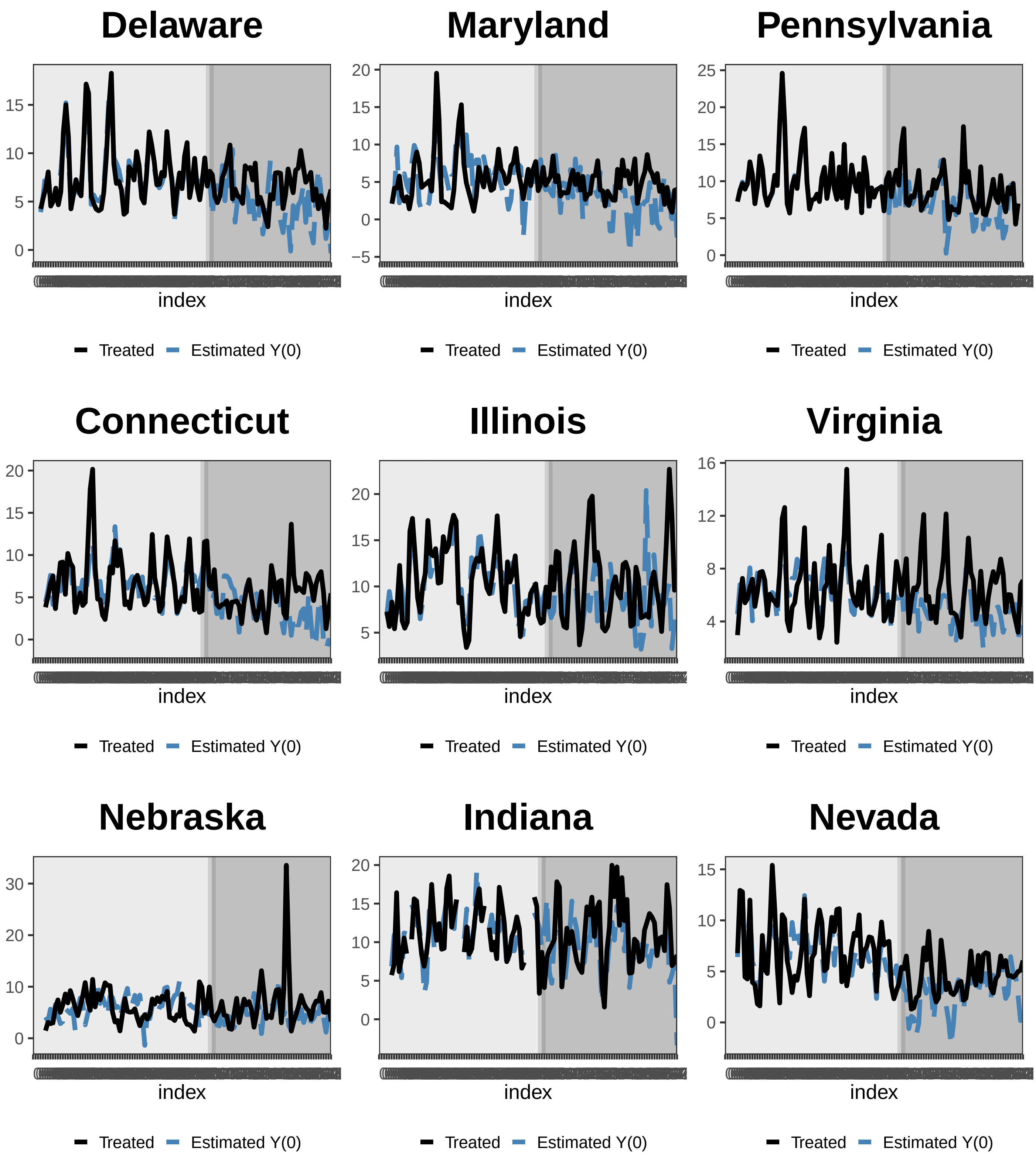
Delaware
Maryland
Pennsylvania
Connecticut
Illinois
Virginia
Nebraska
Indiana
Nevada
index
Treated
Estimated Y(0)

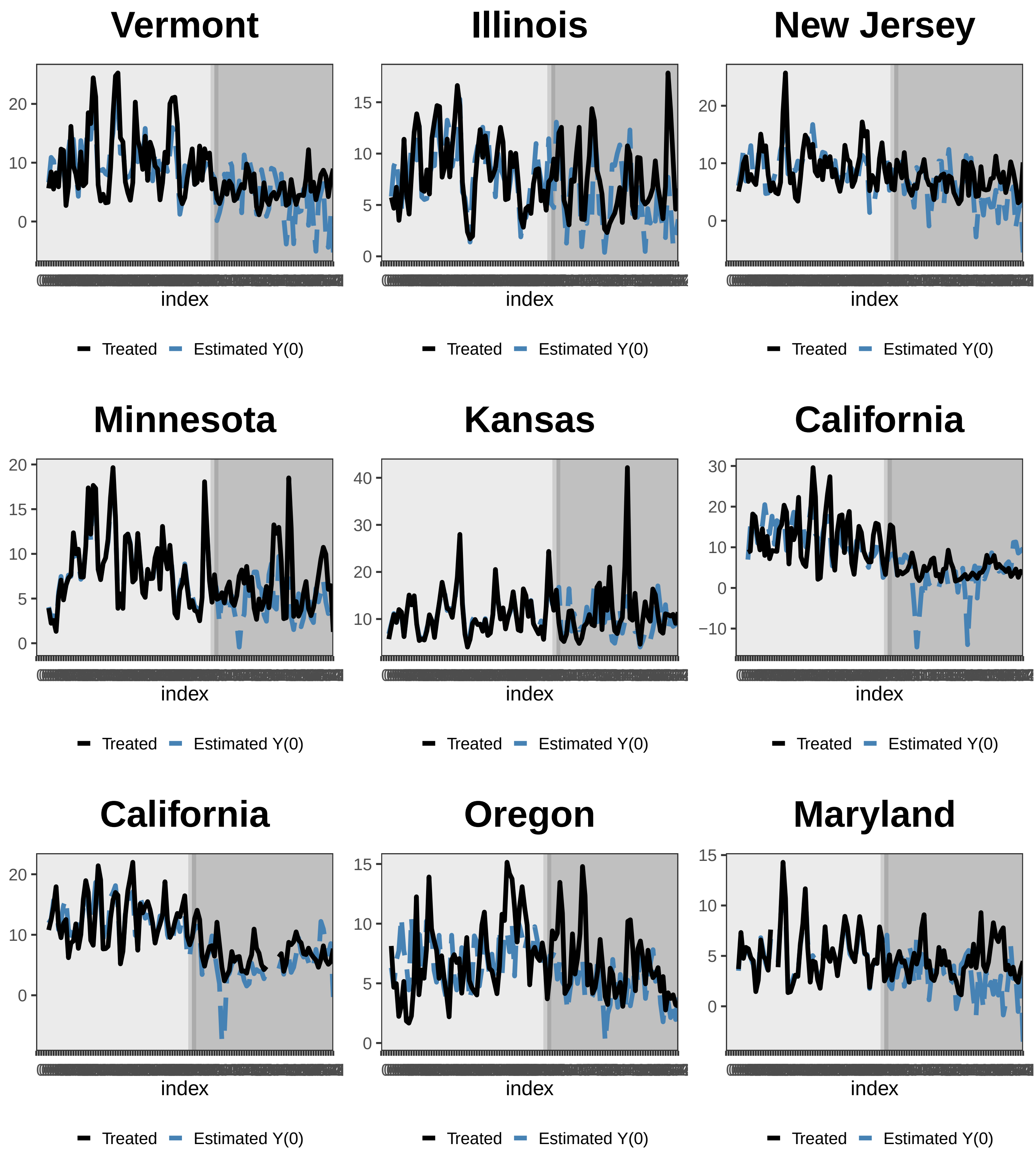
Vermont
index
Treated
Estimated Y(0)
Illinois
index
Treated
Estimated Y(0)
New Jersey
index
Treated
Estimated Y(0)
Minnesota
index
Treated
Estimated Y(0)
Kansas
index
Treated
Estimated Y(0)
California
index
Treated
Estimated Y(0)
California
index
Treated
Estimated Y(0)
Oregon
index
Treated
Estimated Y(0)
Maryland
index
Treated
Estimated Y(0)

### Minnesota

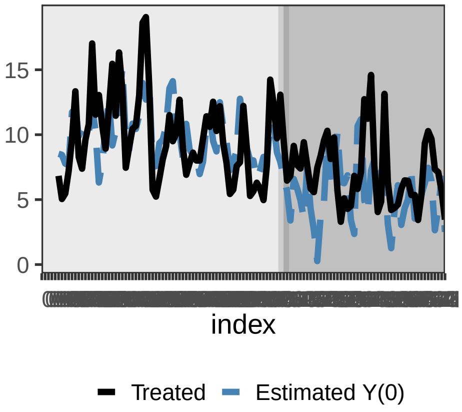


### Missouri

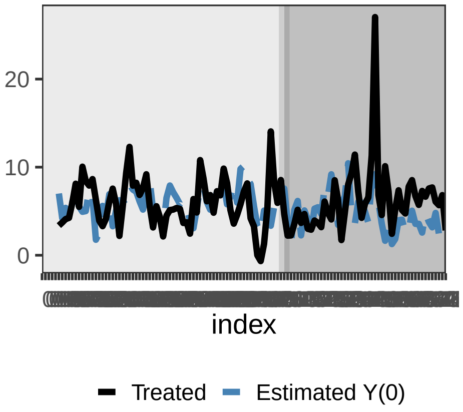


### Oregon

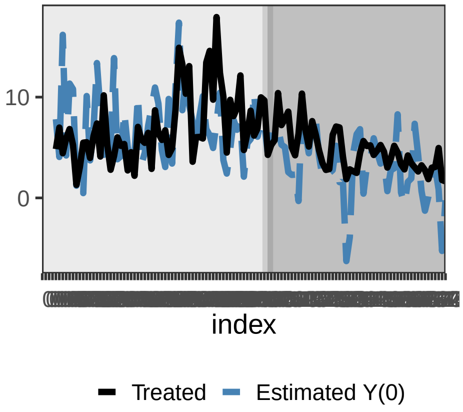


### Pennsylvania

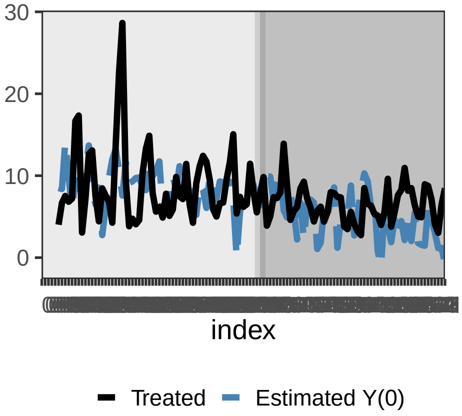


### Indiana

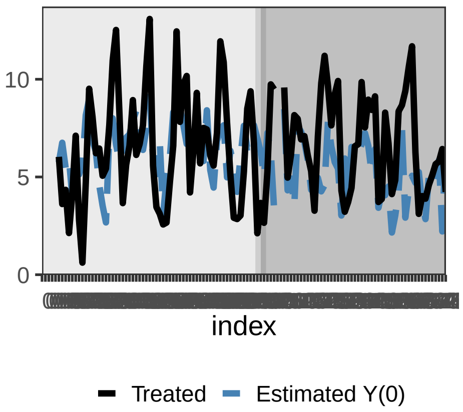


### Louisiana

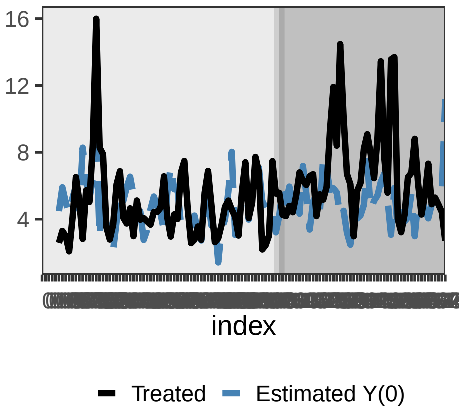


### Idaho

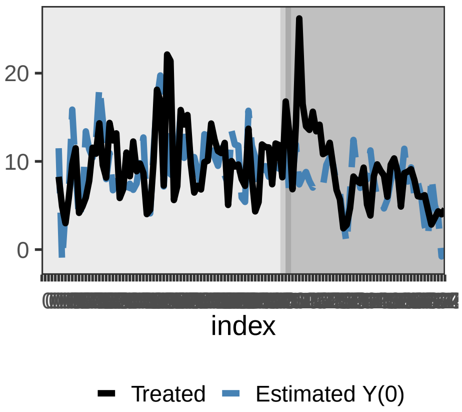


### Kansas

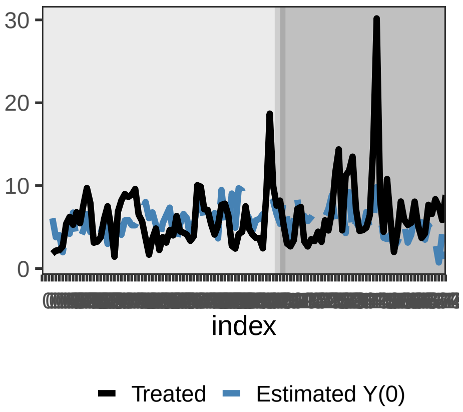


### South Dakota

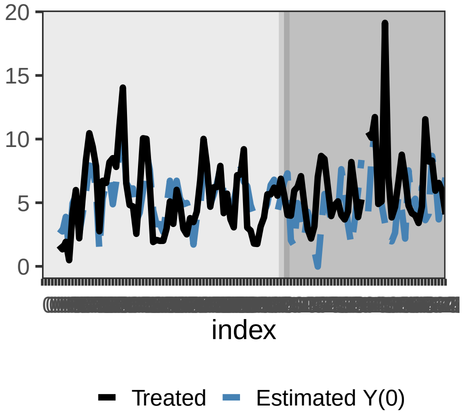

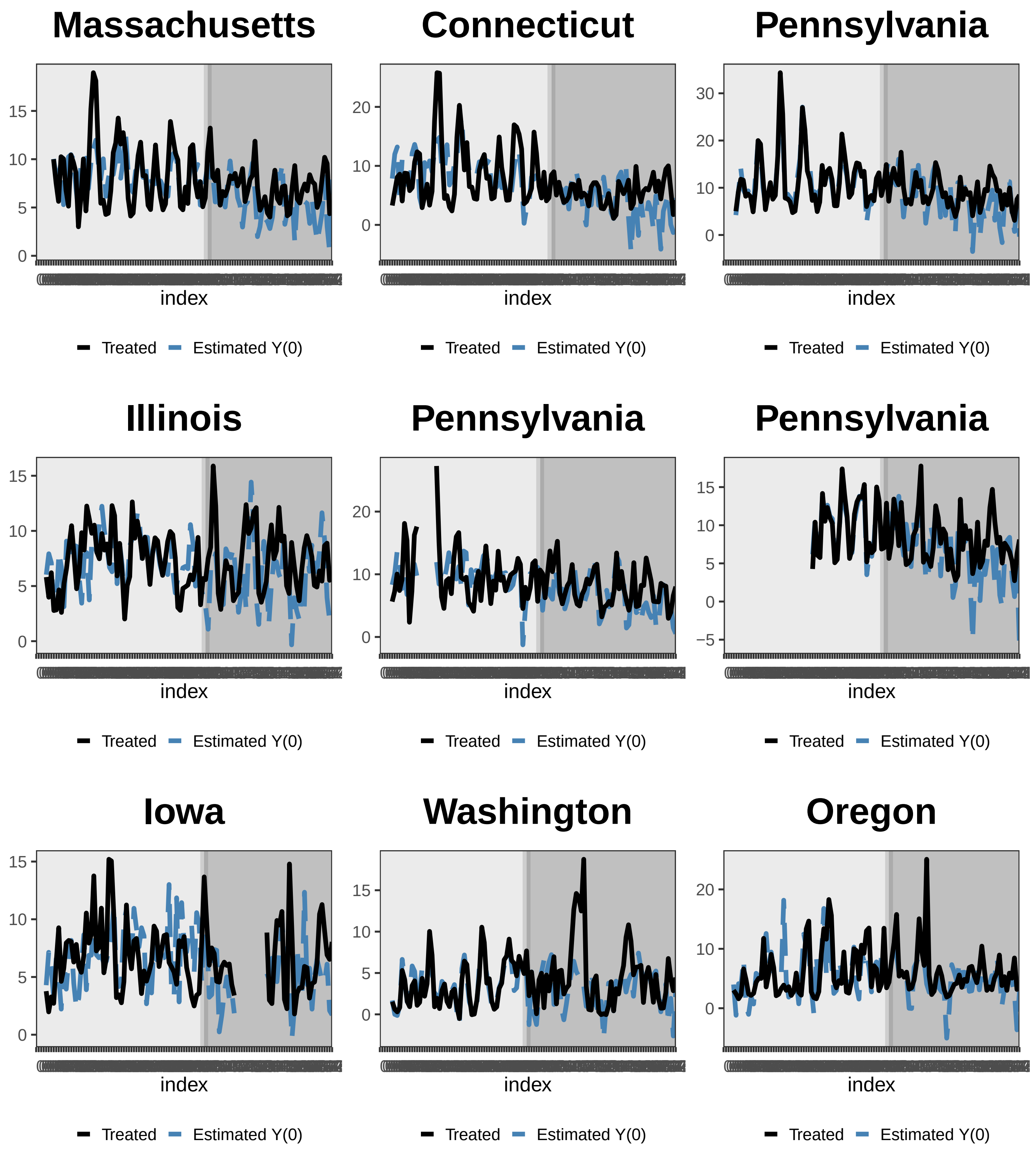
Massachusetts
Connecticut
Pennsylvania
Illinois
Pennsylvania
Pennsylvania
Iowa
Washington
Oregon
index
Treated
Estimated Y(0)

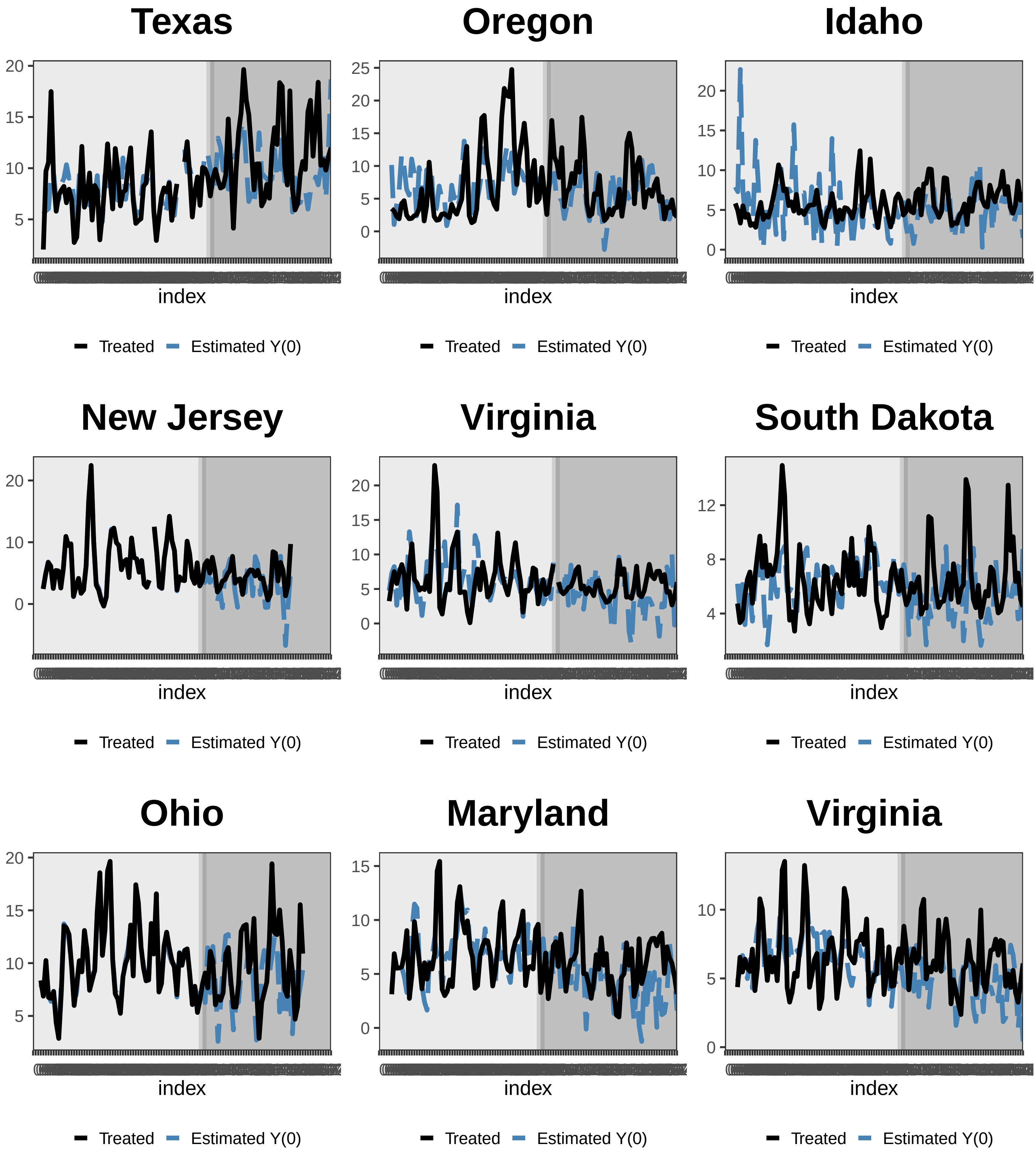
Texas
20
15
10
5
index
Treated
Estimated Y(0)
Oregon
25
20
15
10
5
0
index
Treated
Estimated Y(0)
Idaho
20
15
10
5
0
index
Treated
Estimated Y(0)
New Jersey
20
10
0
index
Treated
Estimated Y(0)
Virginia
20
15
10
5
0
index
Treated
Estimated Y(0)
South Dakota
12
8
4
index
Treated
Estimated Y(0)
Ohio
20
15
10
5
index
Treated
Estimated Y(0)
Maryland
15
10
5
0
index
Treated
Estimated Y(0)
Virginia
10
5
0
index
Treated
Estimated Y(0)

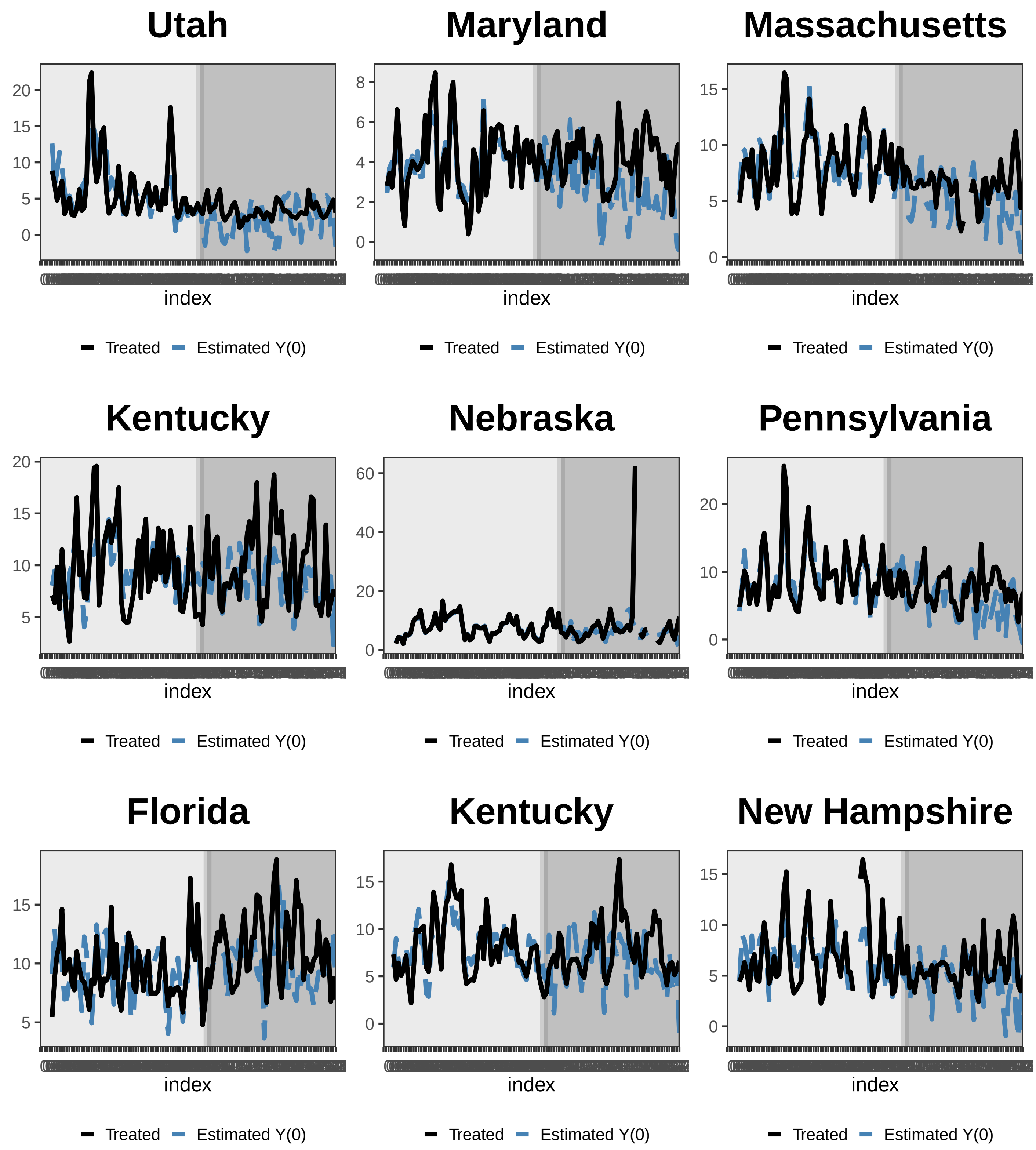
Utah
index
Treated Estimated Y(0)
Maryland
index
Treated Estimated Y(0)
Massachusetts
index
Treated Estimated Y(0)
Kentucky
index
Treated Estimated Y(0)
Nebraska
index
Treated Estimated Y(0)
Pennsylvania
index
Treated Estimated Y(0)
Florida
index
Treated Estimated Y(0)
Kentucky
index
Treated Estimated Y(0)
New Hampshire
index
Treated Estimated Y(0)

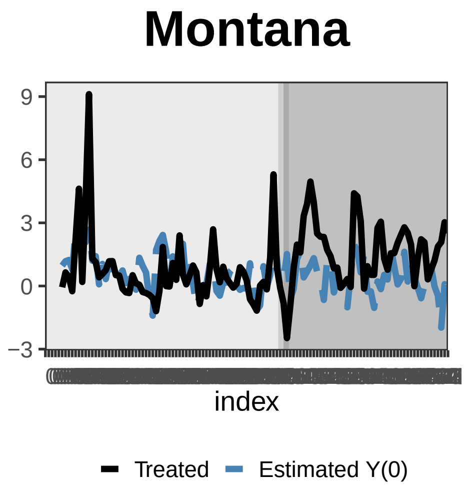
Montana
9
6
3
0
−3
index
Treated
Estimated Y(0)

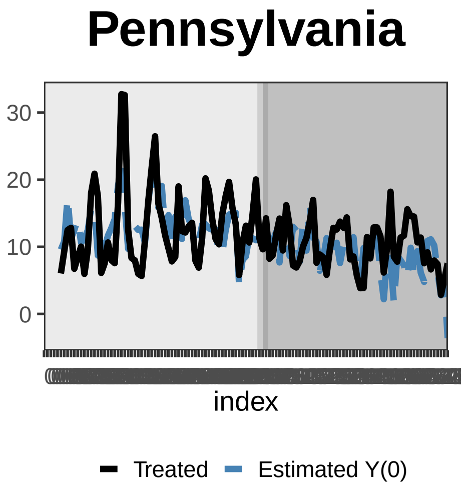
Pennsylvania
30
20
10
0
index
Treated
Estimated Y(0)

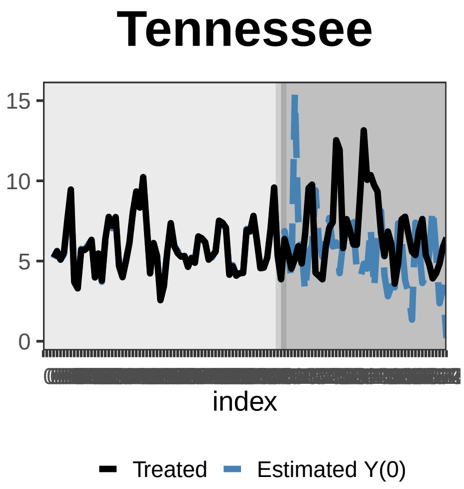
Tennessee
15
10
5
0
index
Treated
Estimated Y(0)

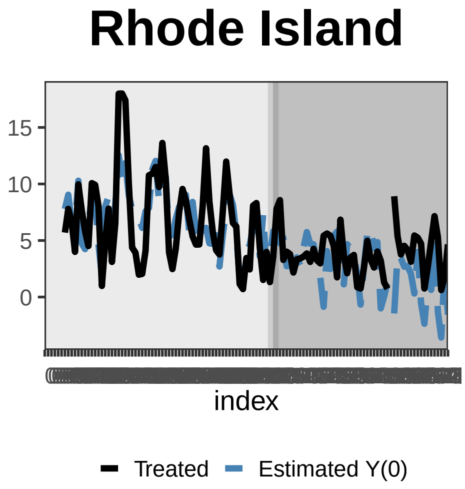
Rhode Island
15
10
5
0
index
Treated
Estimated Y(0)

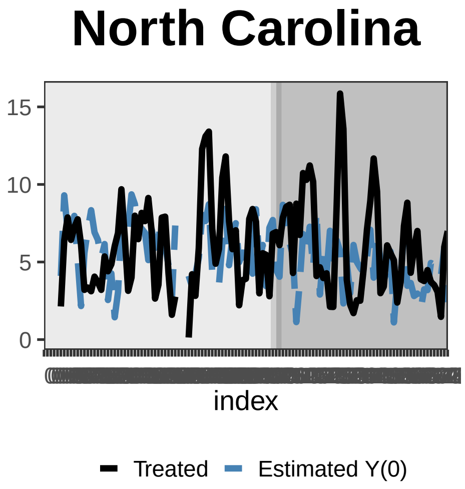
North Carolina
15
10
5
0
index
Treated
Estimated Y(0)

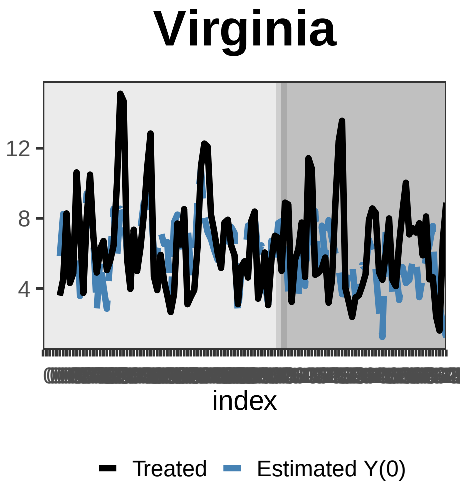
Virginia
12
8
4
index
Treated
Estimated Y(0)

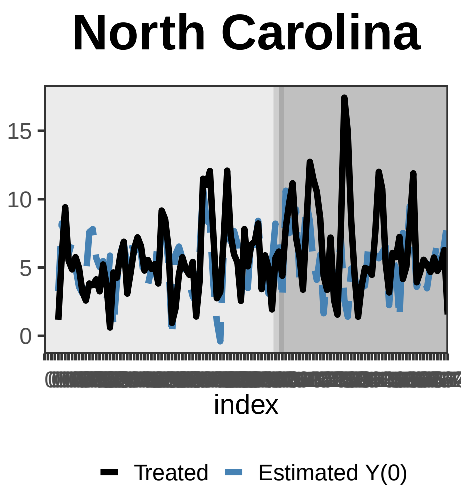
North Carolina
15
10
5
0
index
Treated
Estimated Y(0)

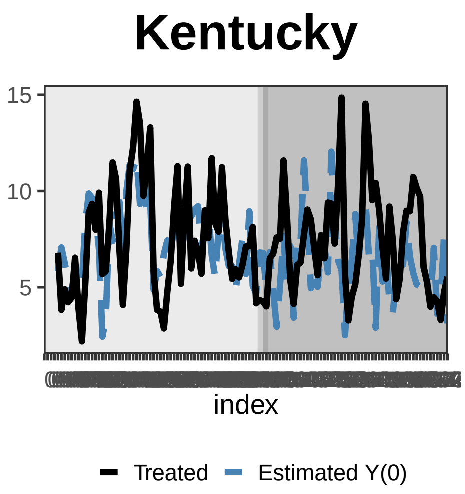
Kentucky
15
10
5
index
Treated
Estimated Y(0)

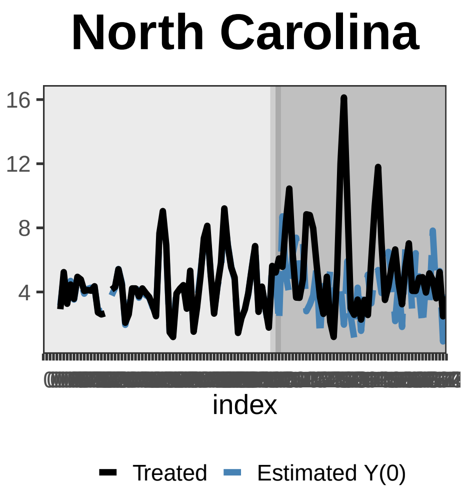
North Carolina
16
12
8
4
index
Treated
Estimated Y(0)

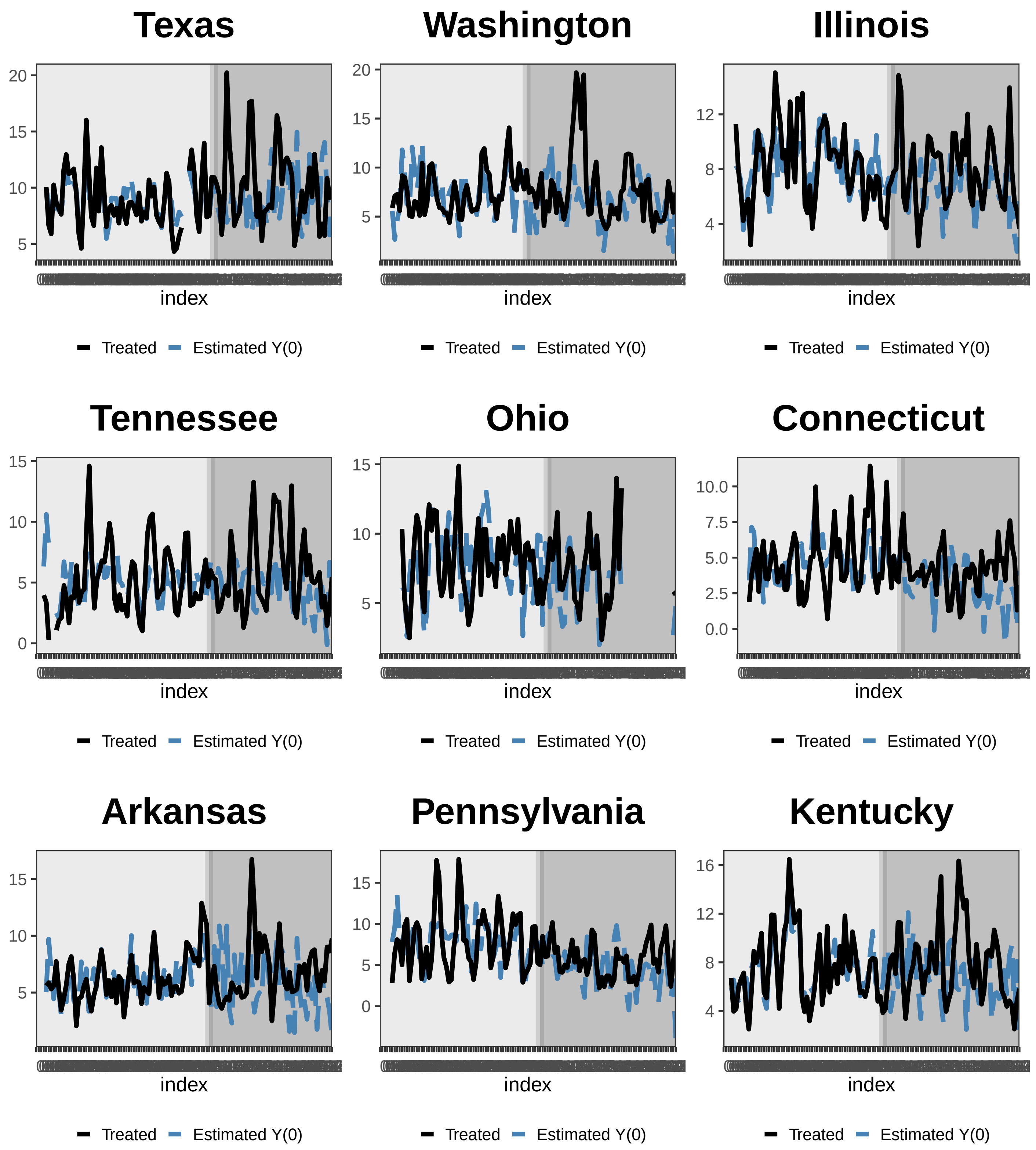
Texas
Washington
Illinois
Tennessee
Ohio
Connecticut
Arkansas
Pennsylvania
Kentucky
index
Treated
Estimated Y(0)

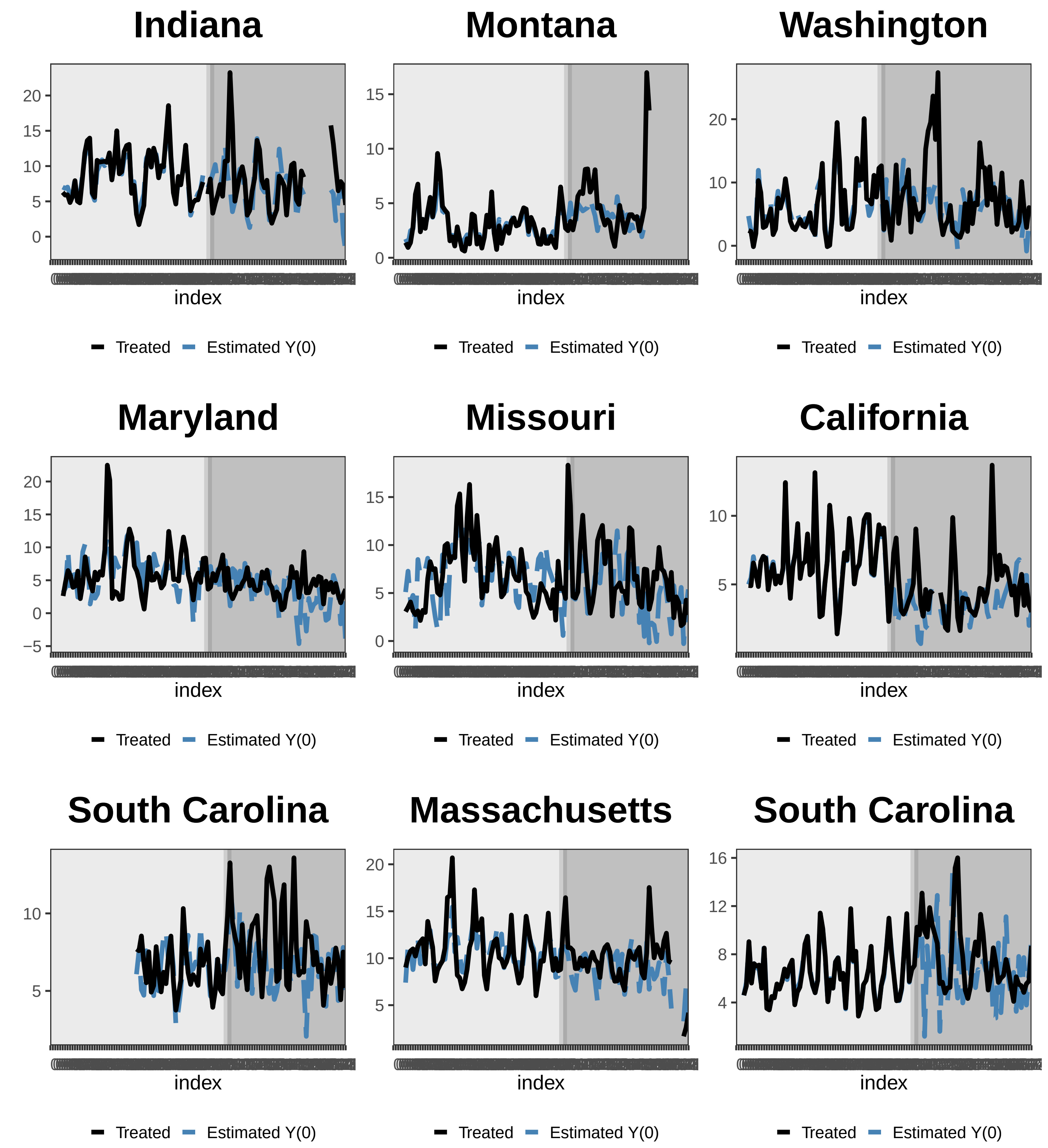
Indiana
Montana
Washington
Maryland
Missouri
California
South Carolina
Massachusetts
South Carolina
index
Treated
Estimated Y(0)

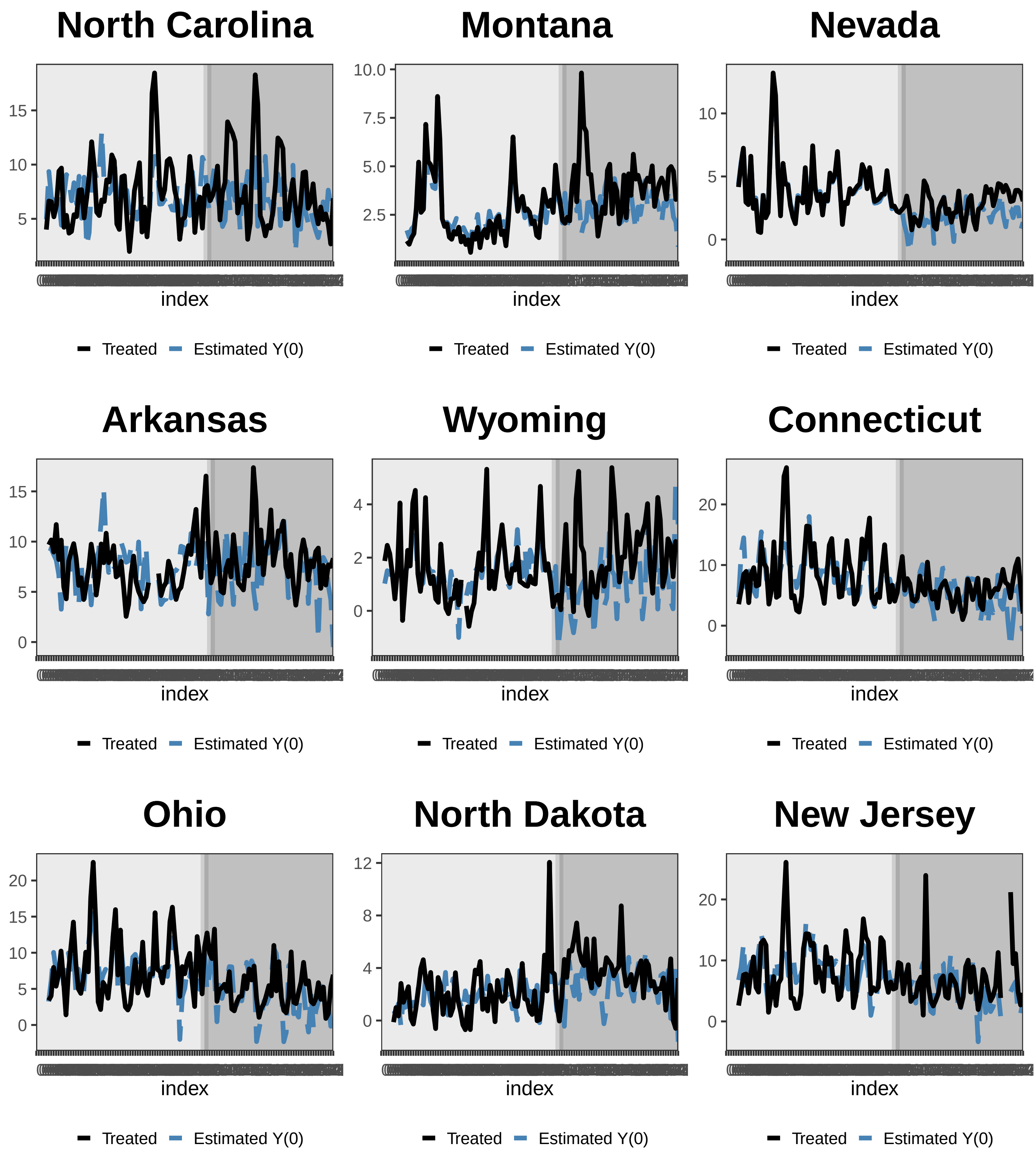
North Carolina
15
10
5
index
Treated
Estimated Y(0)
Montana
10.0
7.5
5.0
2.5
index
Treated
Estimated Y(0)
Nevada
10
5
0
index
Treated
Estimated Y(0)
Arkansas
15
10
5
0
index
Treated
Estimated Y(0)
Wyoming
4
2
0
index
Treated
Estimated Y(0)
Connecticut
20
10
0
index
Treated
Estimated Y(0)
Ohio
20
15
10
5
0
index
Treated
Estimated Y(0)
North Dakota
12
8
4
0
index
Treated
Estimated Y(0)
New Jersey
20
10
0
index
Treated
Estimated Y(0)

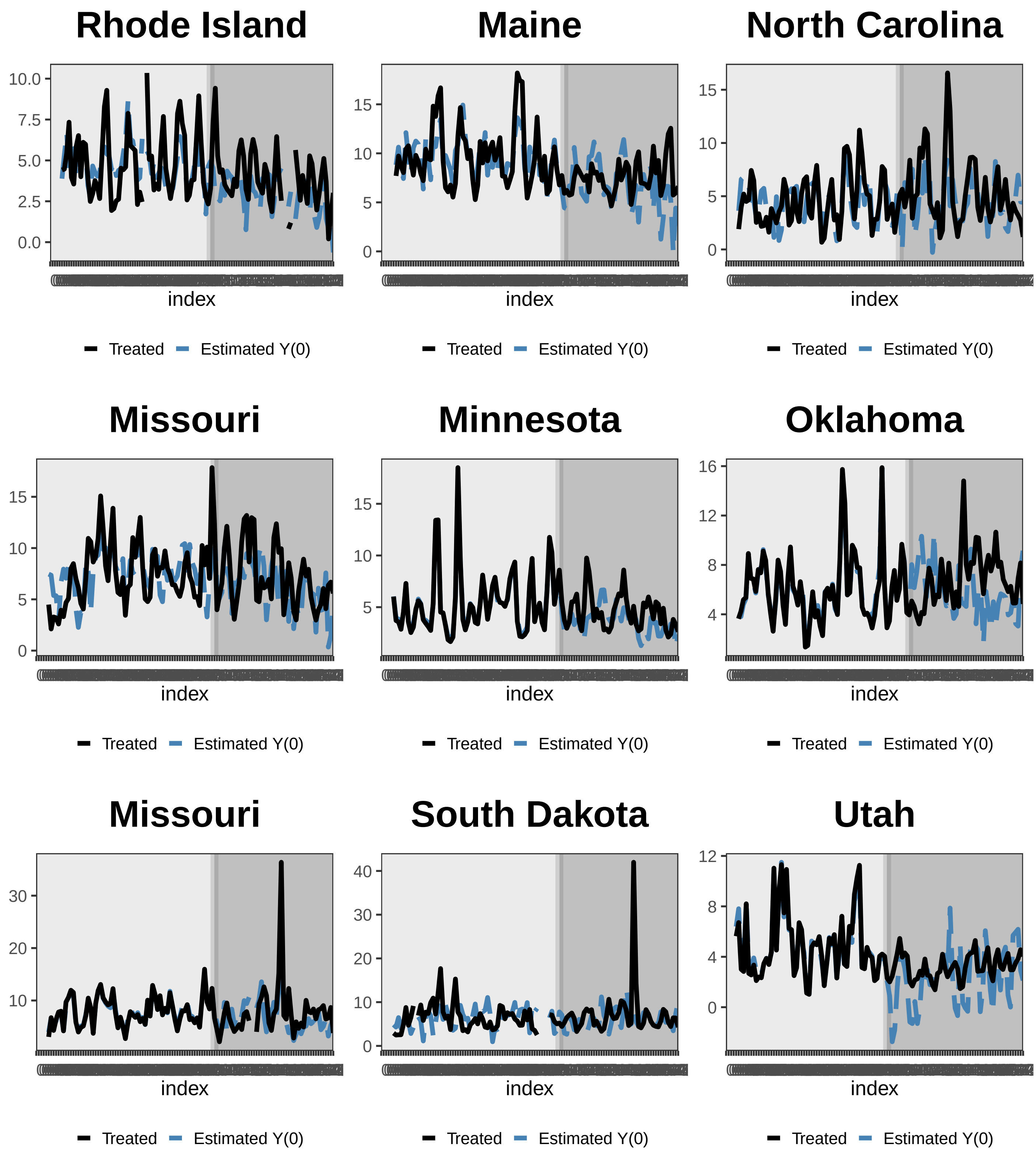
Rhode Island
10.0
7.5
5.0
2.5
0.0
index
Treated
Estimated Y(0)
Maine
15
10
5
0
index
Treated
Estimated Y(0)
North Carolina
15
10
5
0
index
Treated
Estimated Y(0)
Missouri
15
10
5
0
index
Treated
Estimated Y(0)
Minnesota
15
10
5
index
Treated
Estimated Y(0)
Oklahoma
16
12
8
4
index
Treated
Estimated Y(0)
Missouri
30
20
10
index
Treated
Estimated Y(0)
South Dakota
40
30
20
10
0
index
Treated
Estimated Y(0)
Utah
12
8
4
0
index
Treated
Estimated Y(0)

### New Jersey

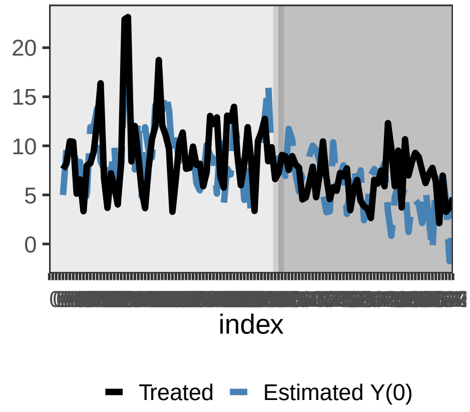


### Maryland

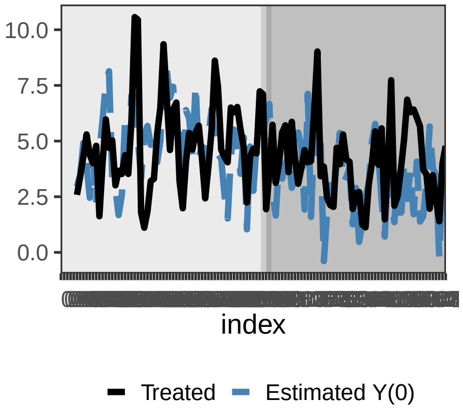


### Missouri

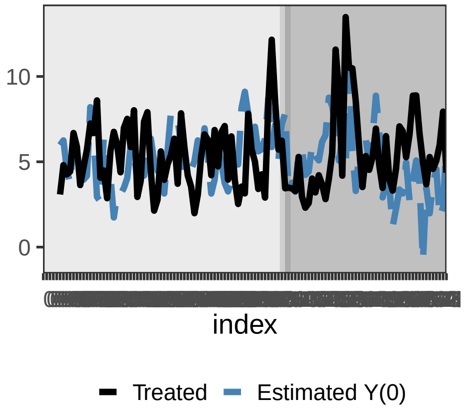


### Washington

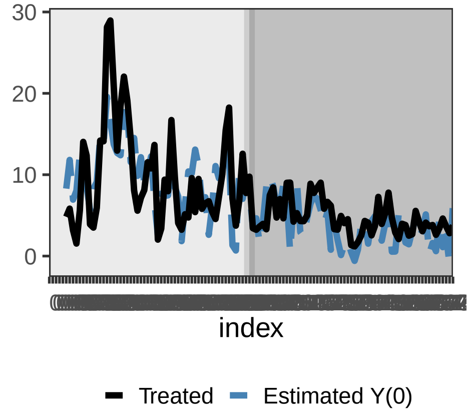


### Maryland

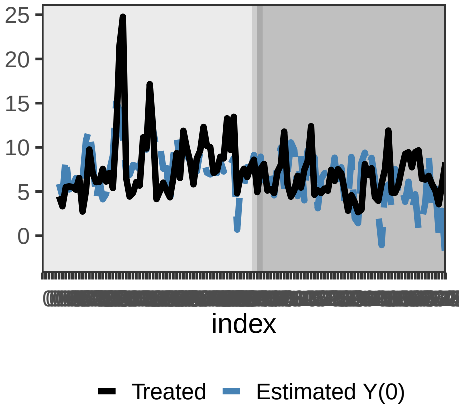


### Florida

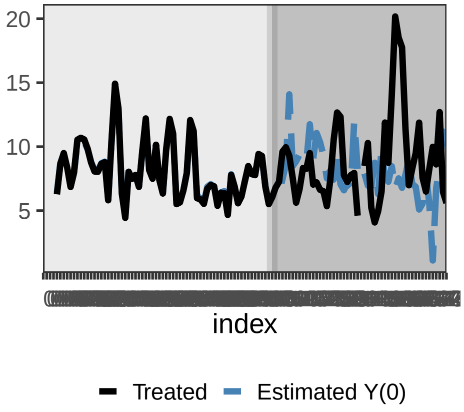


### Missouri

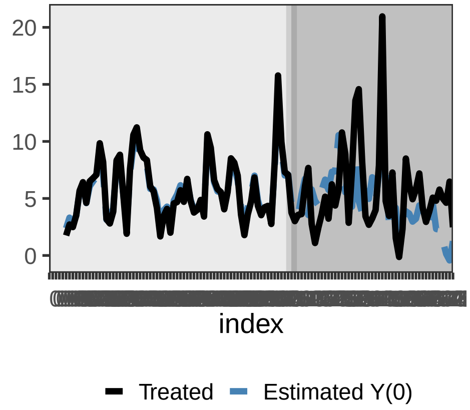


### Alabama

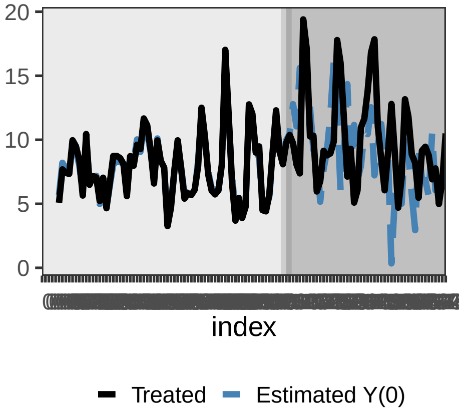


### Florida

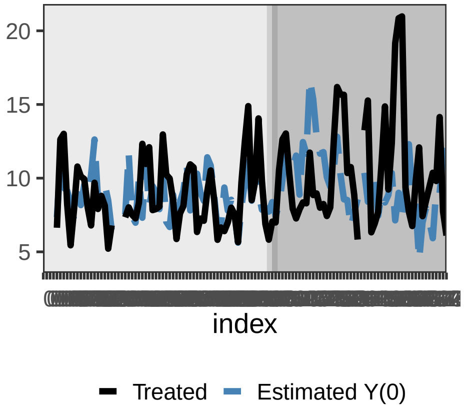

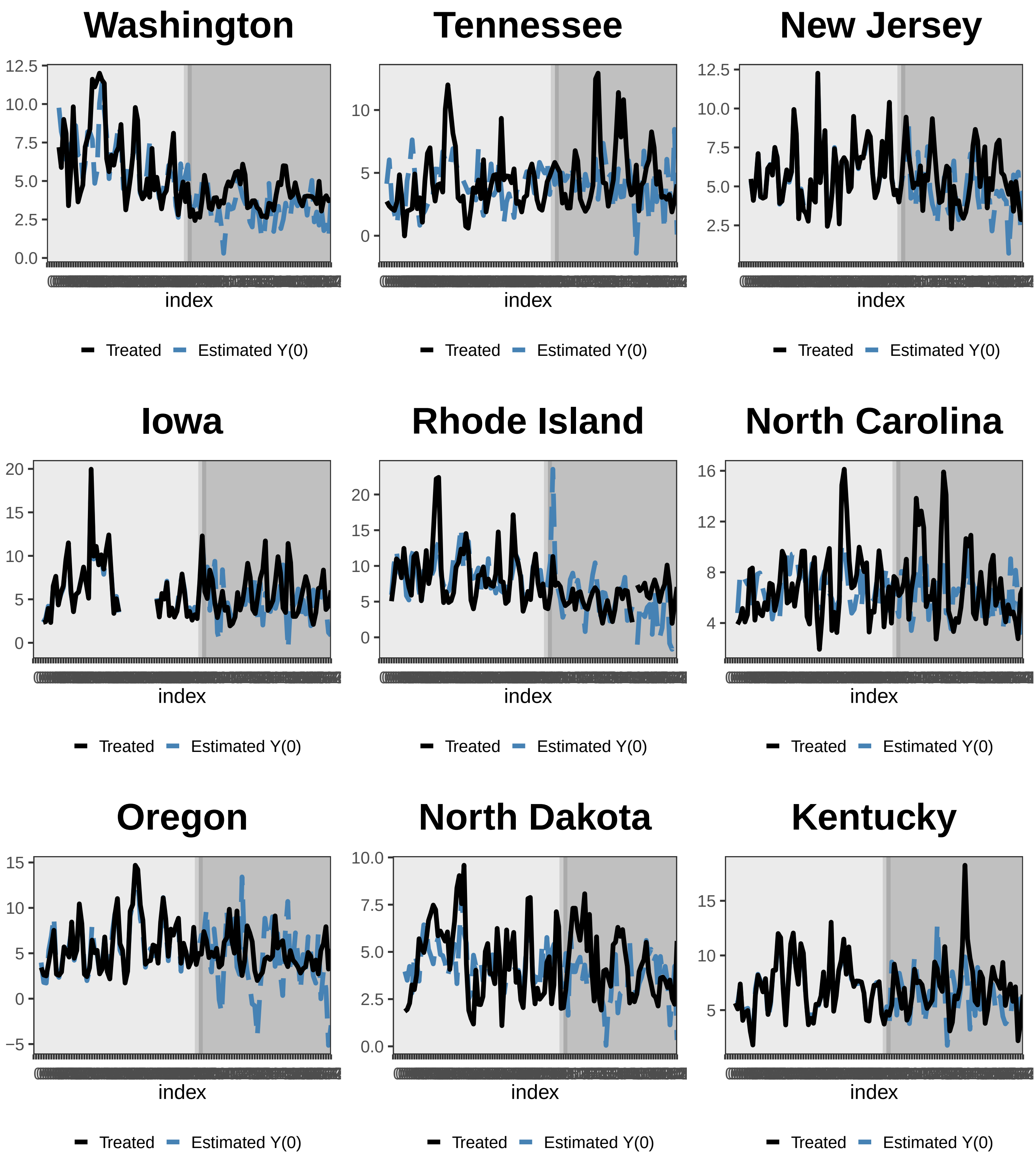
Washington
12.5
10.0
7.5
5.0
2.5
0.0
index
Treated
Estimated Y(0)
Tennessee
10
5
0
index
Treated
Estimated Y(0)
New Jersey
12.5
10.0
7.5
5.0
2.5
index
Treated
Estimated Y(0)
Iowa
20
15
10
5
0
index
Treated
Estimated Y(0)
Rhode Island
20
15
10
5
0
index
Treated
Estimated Y(0)
North Carolina
16
12
8
4
index
Treated
Estimated Y(0)
Oregon
15
10
5
0
−5
index
Treated
Estimated Y(0)
North Dakota
10.0
7.5
5.0
2.5
0.0
index
Treated
Estimated Y(0)
Kentucky
15
10
5
index
Treated
Estimated Y(0)

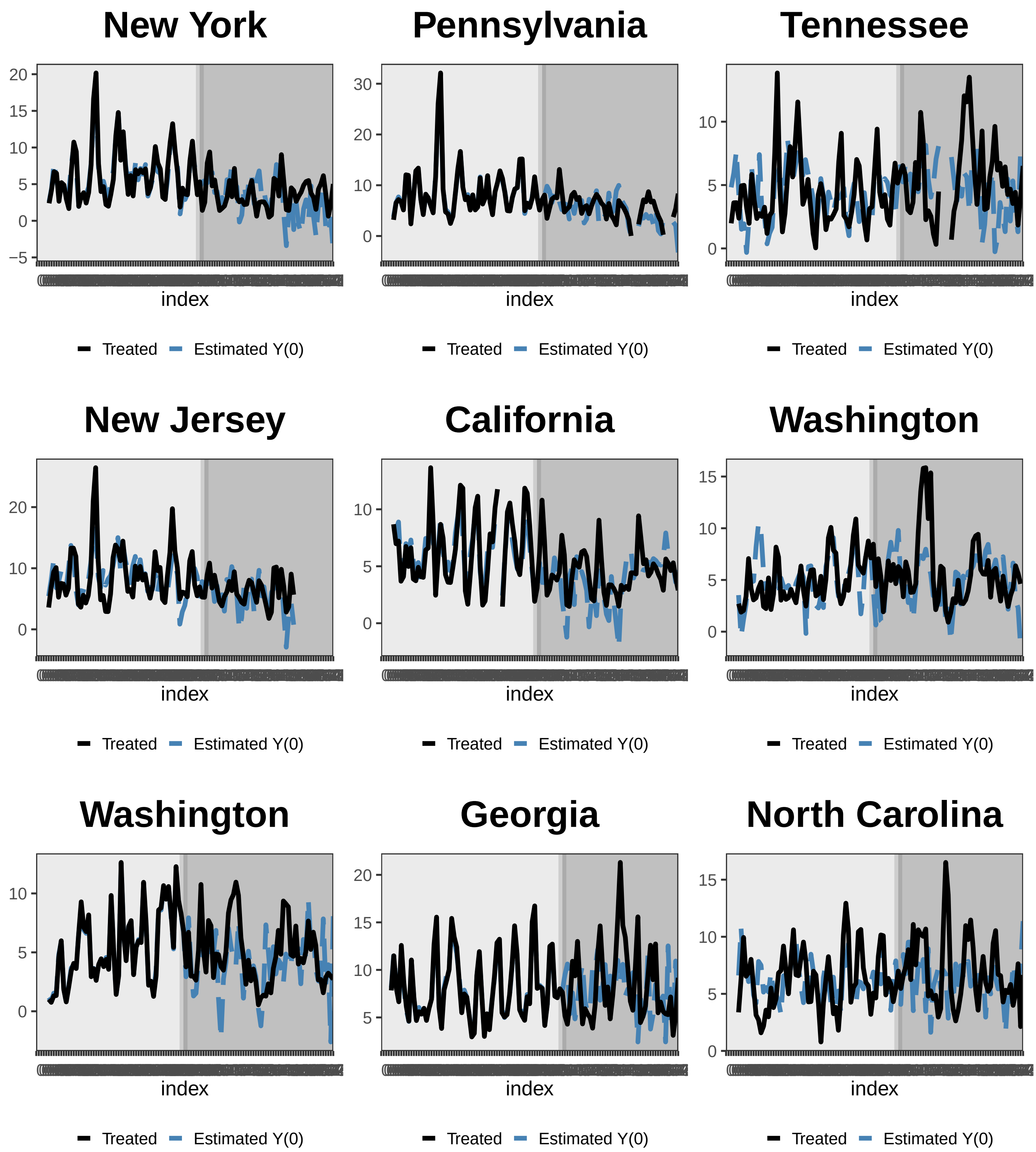
New York
Pennsylvania
Tennessee
New Jersey
California
Washington
Washington
Georgia
North Carolina
index
Treated
Estimated Y(0)

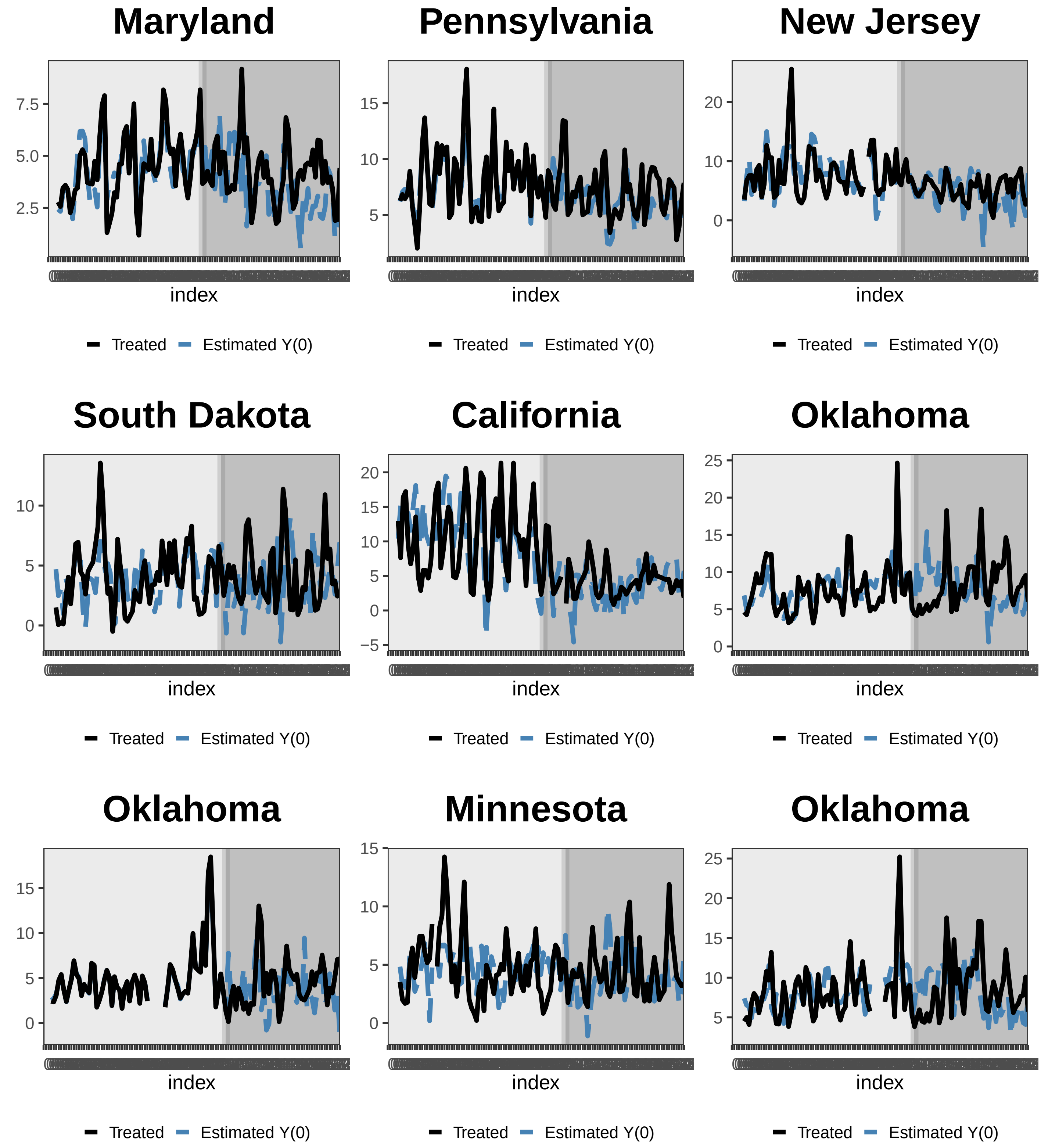
Maryland
7.5
5.0
2.5
index
Treated
Estimated Y(0)
Pennsylvania
15
10
5
index
Treated
Estimated Y(0)
New Jersey
20
10
0
index
Treated
Estimated Y(0)
South Dakota
10
5
0
index
Treated
Estimated Y(0)
California
20
15
10
5
0
−5
index
Treated
Estimated Y(0)
Oklahoma
25
20
15
10
5
0
index
Treated
Estimated Y(0)
Oklahoma
15
10
5
0
index
Treated
Estimated Y(0)
Minnesota
15
10
5
0
index
Treated
Estimated Y(0)
Oklahoma
25
20
15
10
5
index
Treated
Estimated Y(0)

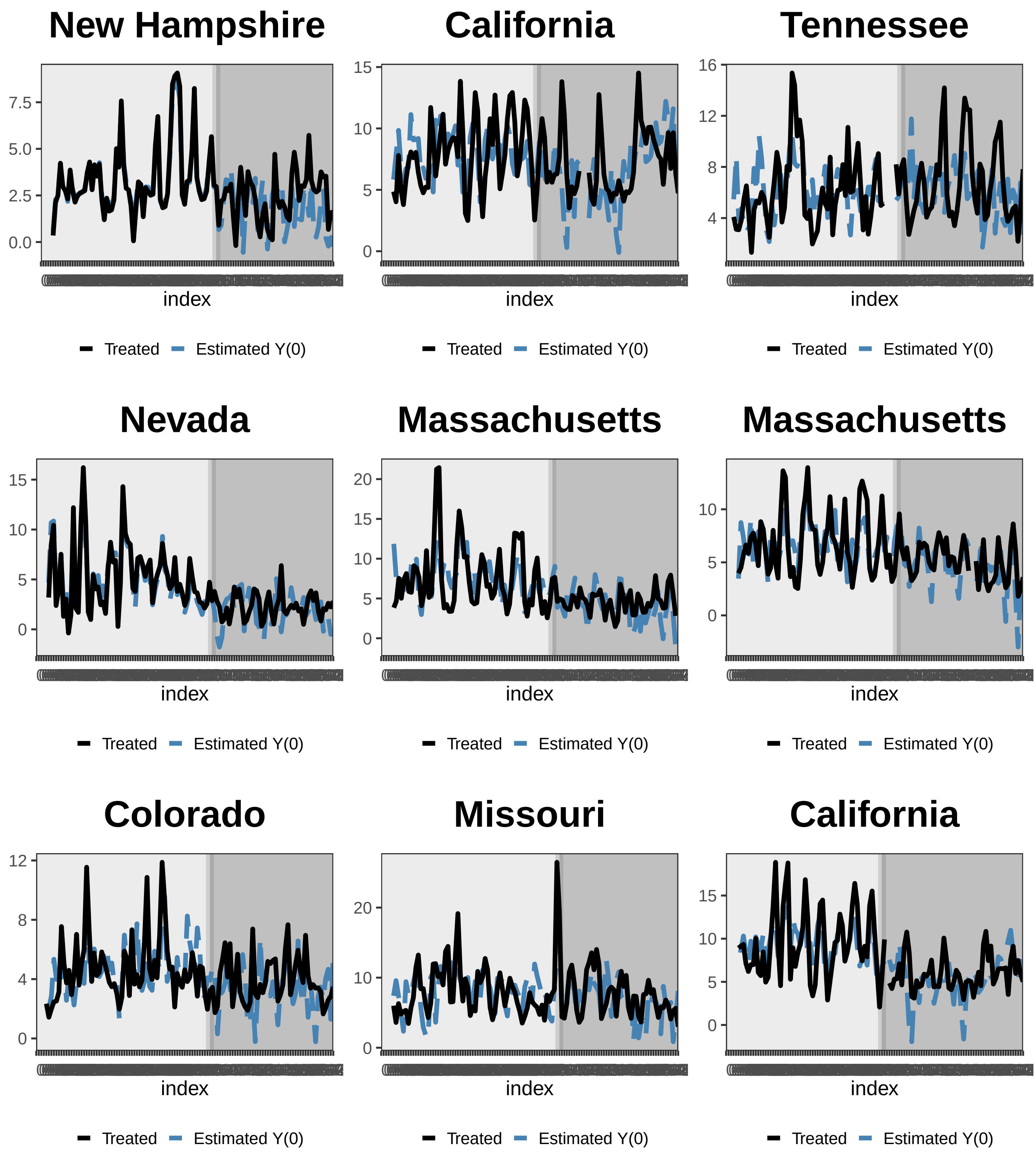
New Hampshire
7.5
5.0
2.5
0.0
index
Treated
Estimated Y(0)
California
15
10
5
0
index
Treated
Estimated Y(0)
Tennessee
16
12
8
4
index
Treated
Estimated Y(0)
Nevada
15
10
5
0
index
Treated
Estimated Y(0)
Massachusetts
20
15
10
5
0
index
Treated
Estimated Y(0)
Massachusetts
10
5
0
index
Treated
Estimated Y(0)
Colorado
12
8
4
0
index
Treated
Estimated Y(0)
Missouri
20
10
0
index
Treated
Estimated Y(0)
California
15
10
5
0
index
Treated
Estimated Y(0)

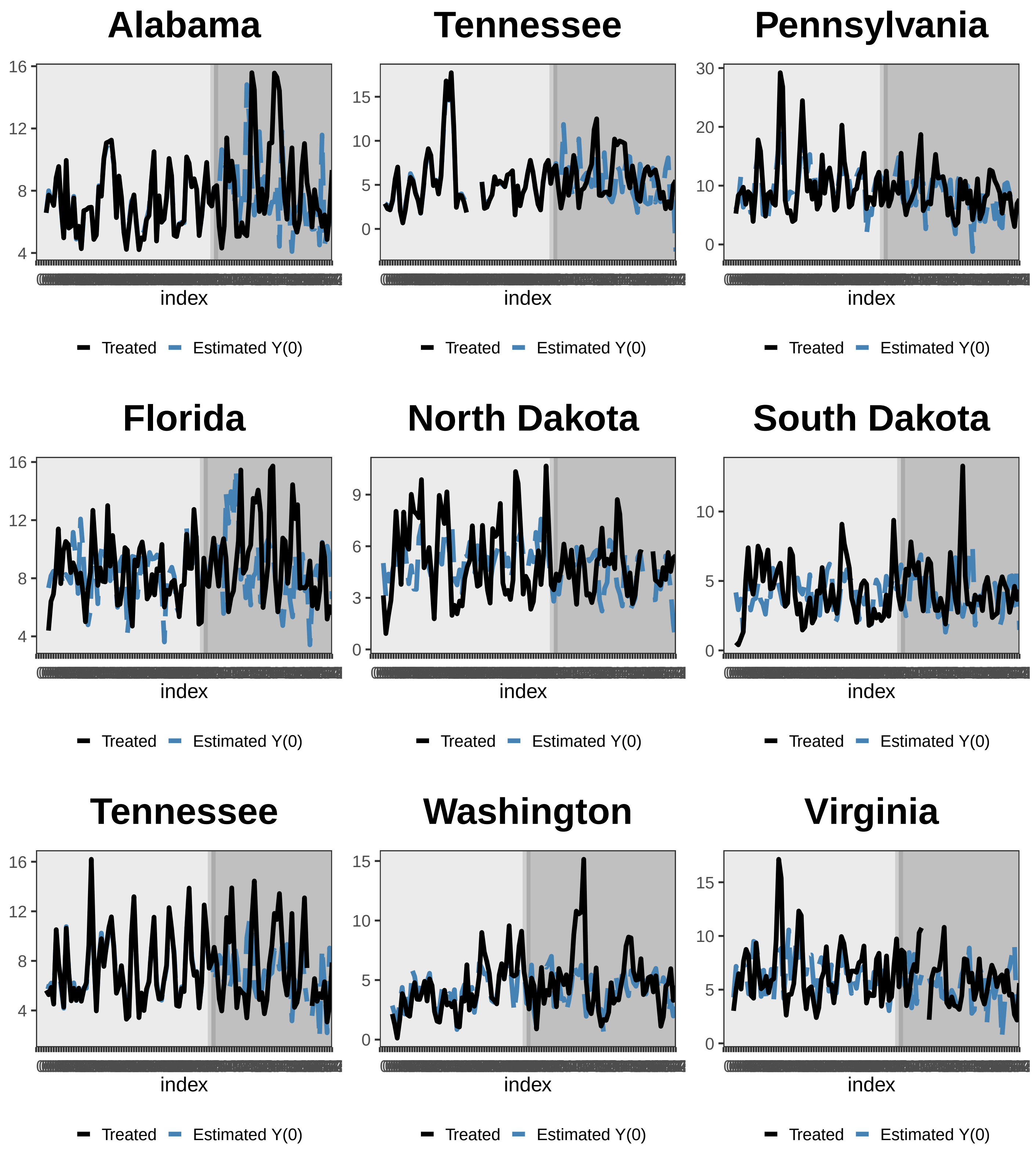
Alabama
16
12
8
4
index
Treated
Estimated Y(0)
Tennessee
15
10
5
0
index
Treated
Estimated Y(0)
Pennsylvania
30
20
10
0
index
Treated
Estimated Y(0)
Florida
16
12
8
4
index
Treated
Estimated Y(0)
North Dakota
9
6
3
0
index
Treated
Estimated Y(0)
South Dakota
10
5
0
index
Treated
Estimated Y(0)
Tennessee
16
12
8
4
index
Treated
Estimated Y(0)
Washington
15
10
5
0
index
Treated
Estimated Y(0)
Virginia
15
10
5
0
index
Treated
Estimated Y(0)

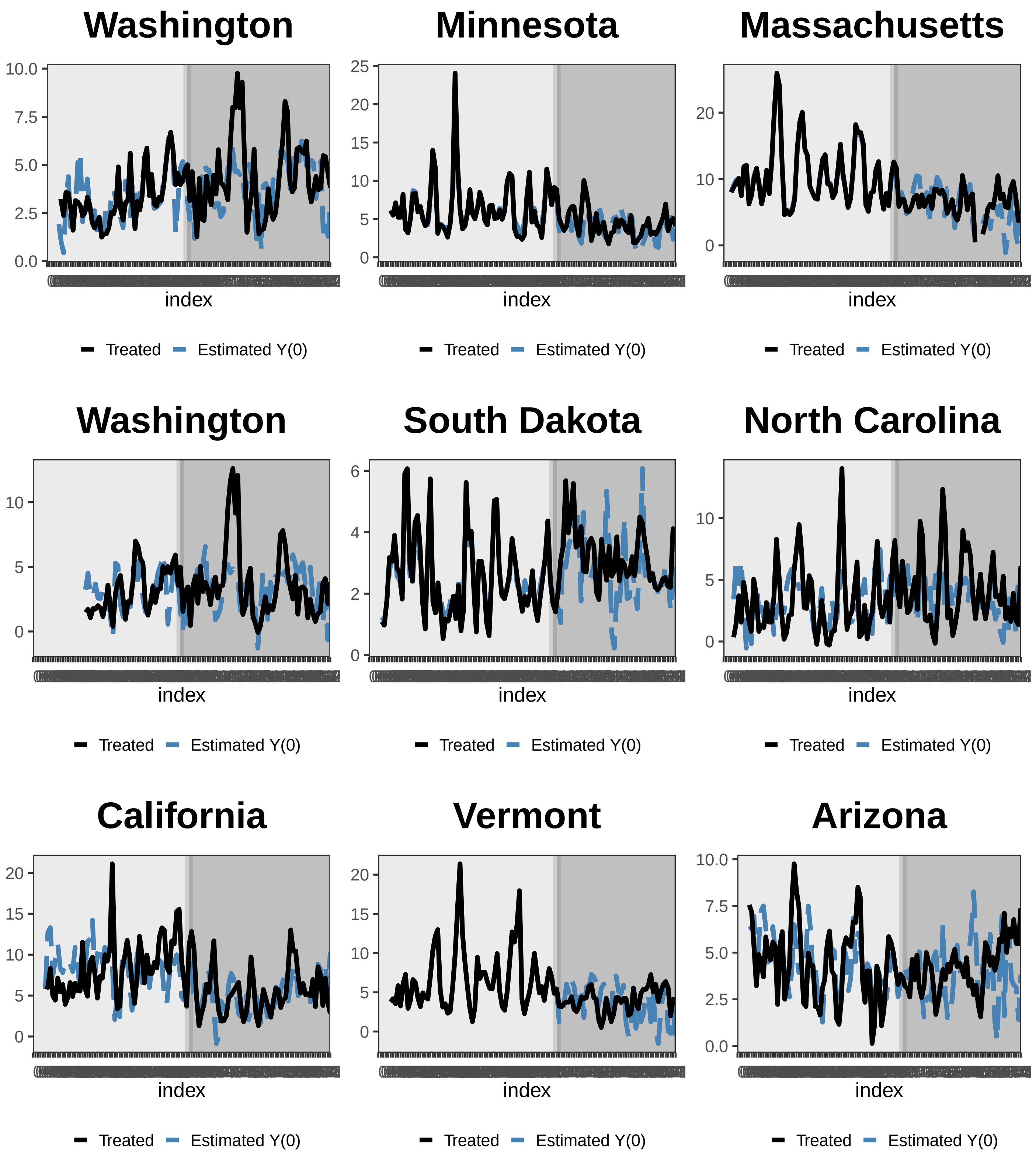
Washington
10.0
7.5
5.0
2.5
0.0
index
Treated
Estimated Y(0)
Minnesota
25
20
15
10
5
0
index
Treated
Estimated Y(0)
Massachusetts
20
10
0
index
Treated
Estimated Y(0)
Washington
10
5
0
index
Treated
Estimated Y(0)
South Dakota
6
4
2
0
index
Treated
Estimated Y(0)
North Carolina
10
5
0
index
Treated
Estimated Y(0)
California
20
15
10
5
0
index
Treated
Estimated Y(0)
Vermont
20
15
10
5
0
index
Treated
Estimated Y(0)
Arizona
10.0
7.5
5.0
2.5
0.0
index
Treated
Estimated Y(0)

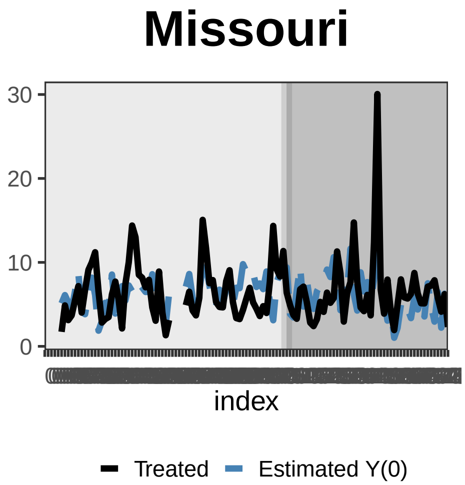
Missouri
30
20
10
0
index
Treated
Estimated Y(0)

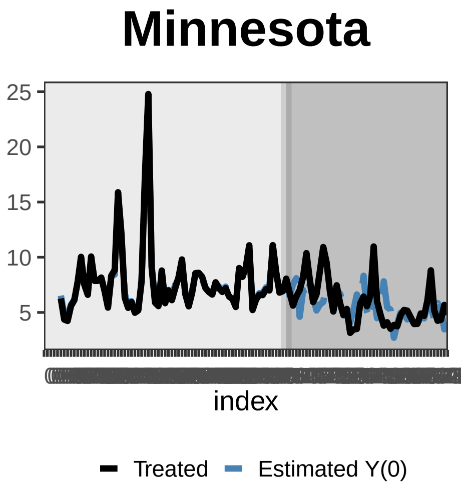
Minnesota
25
20
15
10
5
index
Treated
Estimated Y(0)

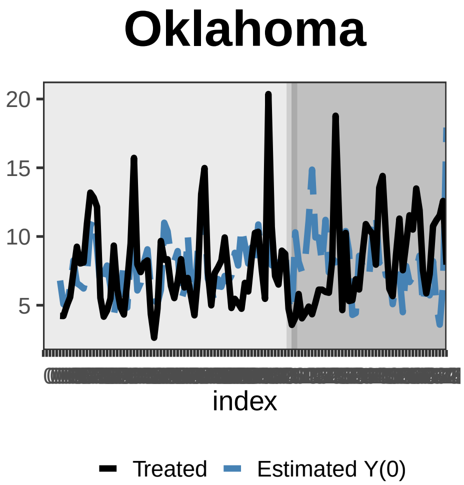
Oklahoma
20
15
10
5
index
Treated
Estimated Y(0)

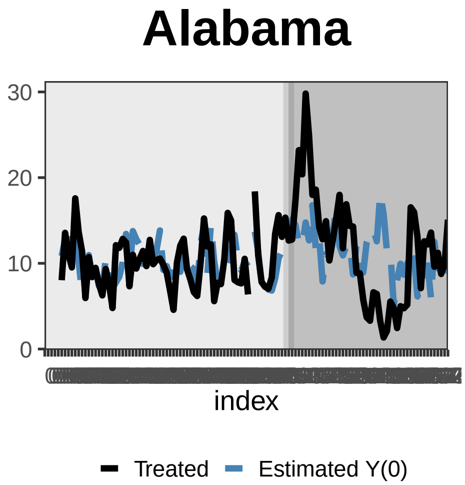
Alabama
30
20
10
0
index
Treated
Estimated Y(0)

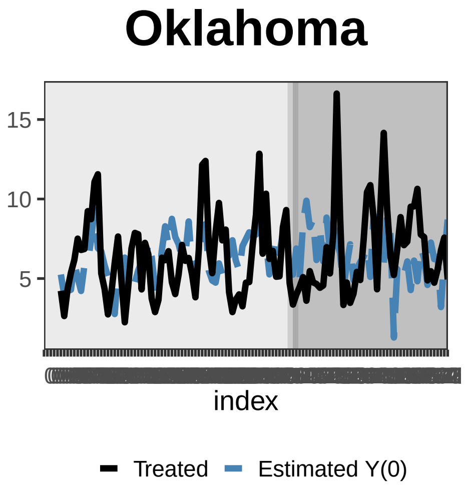
Oklahoma
15
10
5
index
Treated
Estimated Y(0)

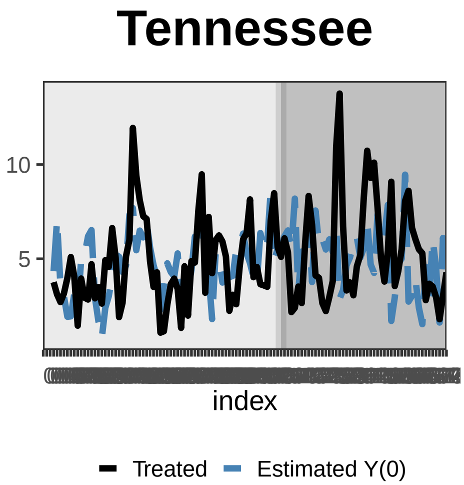
Tennessee
10
5
index
Treated
Estimated Y(0)

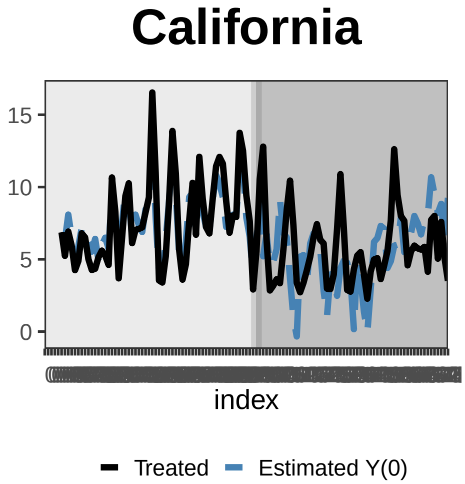
California
15
10
5
0
index
Treated
Estimated Y(0)

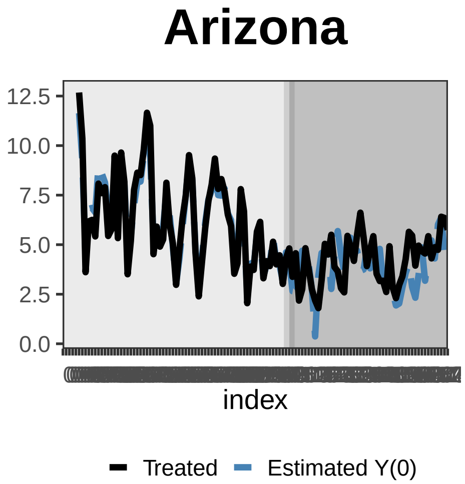
Arizona
12.5
10.0
7.5
5.0
2.5
0.0
index
Treated
Estimated Y(0)

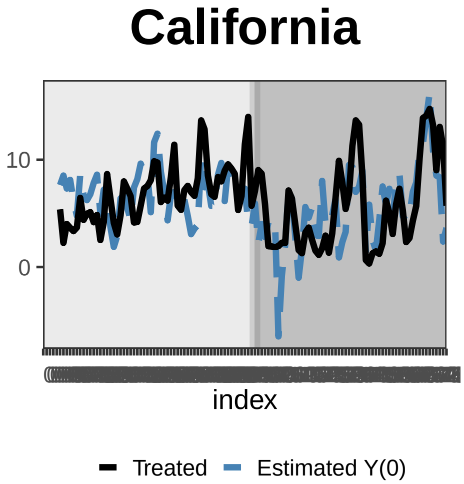
California
10
0
index
Treated
Estimated Y(0)

### South Carolina

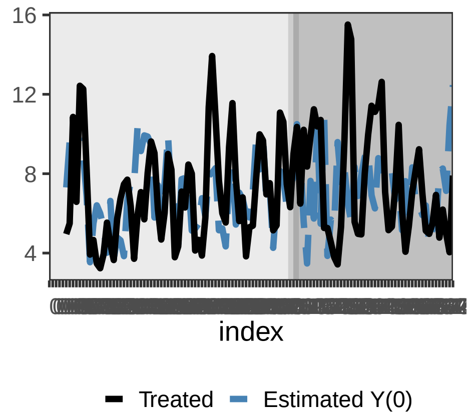


### California

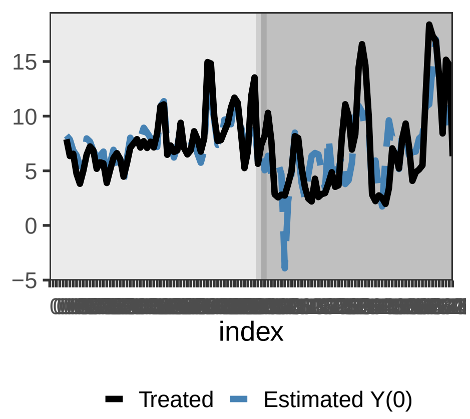


### Montana

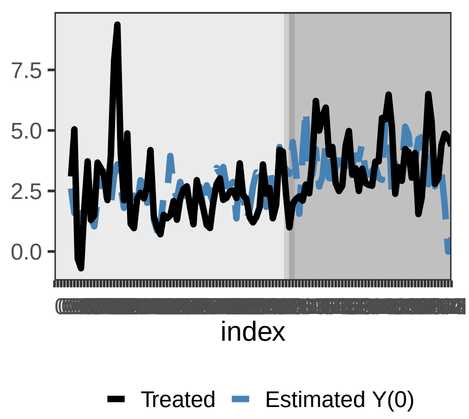


### Kentucky

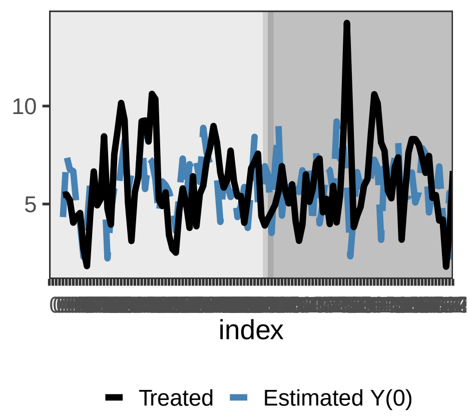


### Illinois

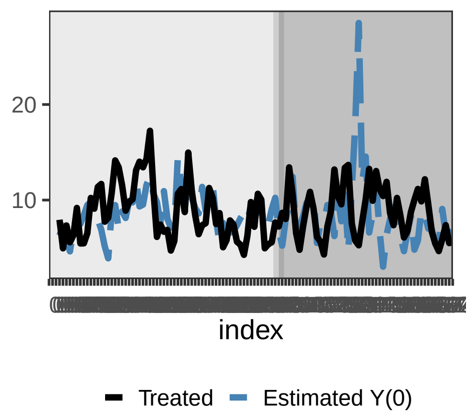


### Minnesota

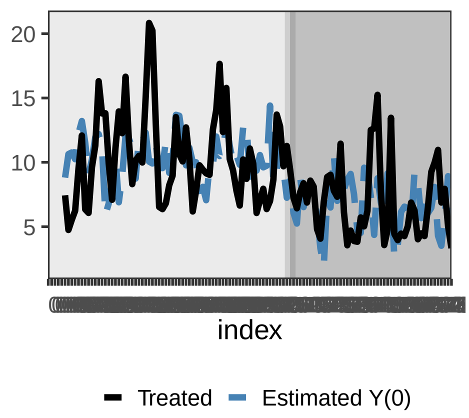


### Pennsylvania

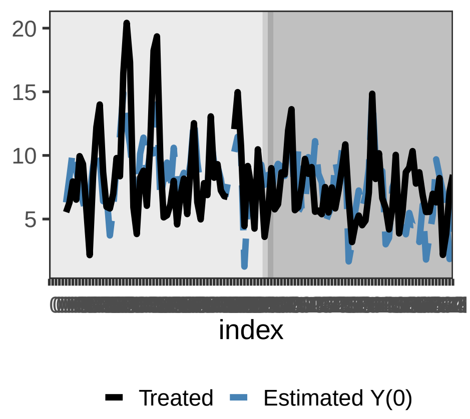


### North Dakota

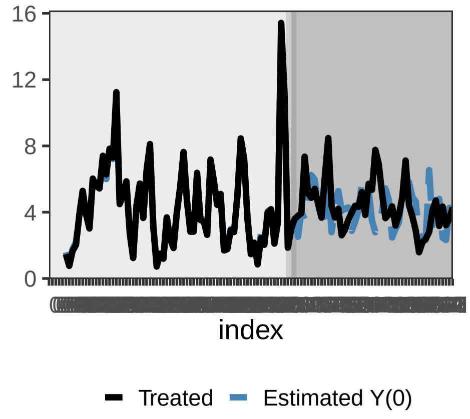


### New York

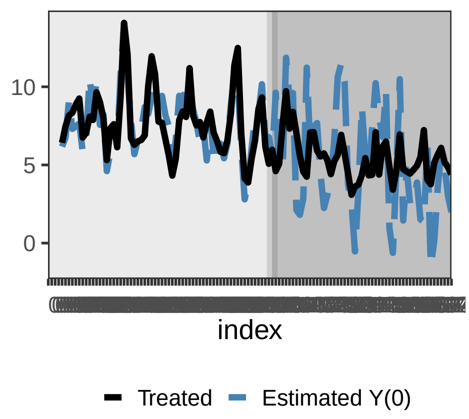

### Oklahoma

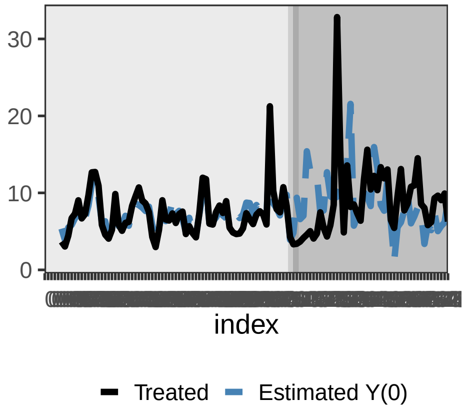


### South Dakota

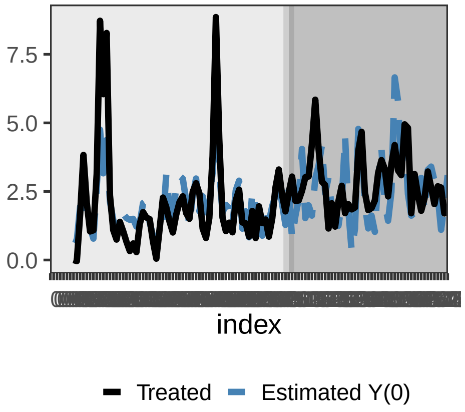


### Pennsylvania

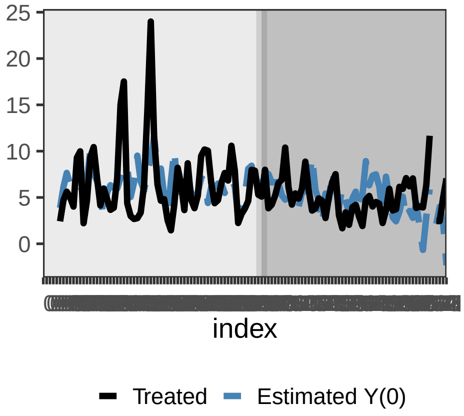


### Oklahoma

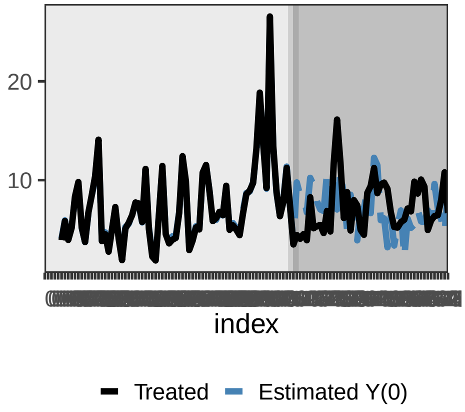


### California

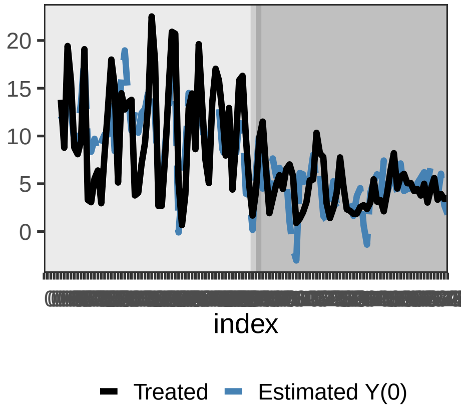


### California

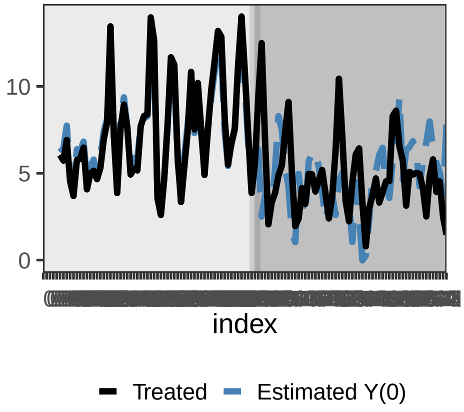


### Colorado

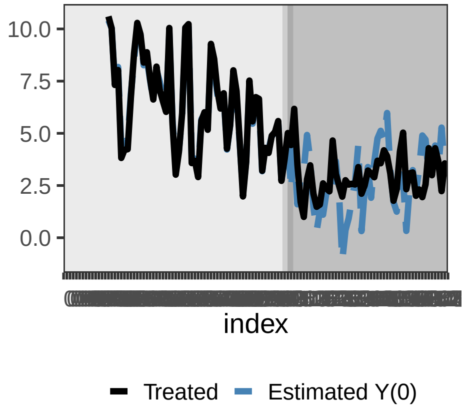


### Wyoming

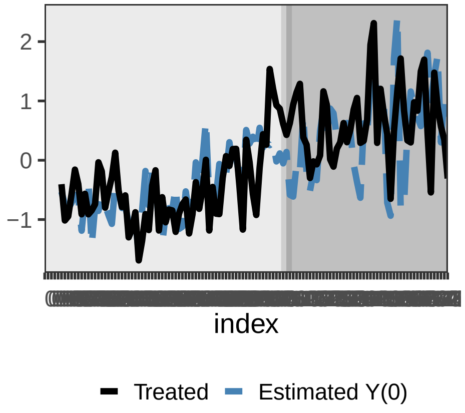


### Missouri

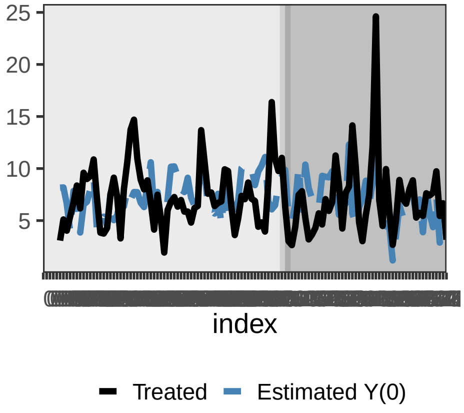

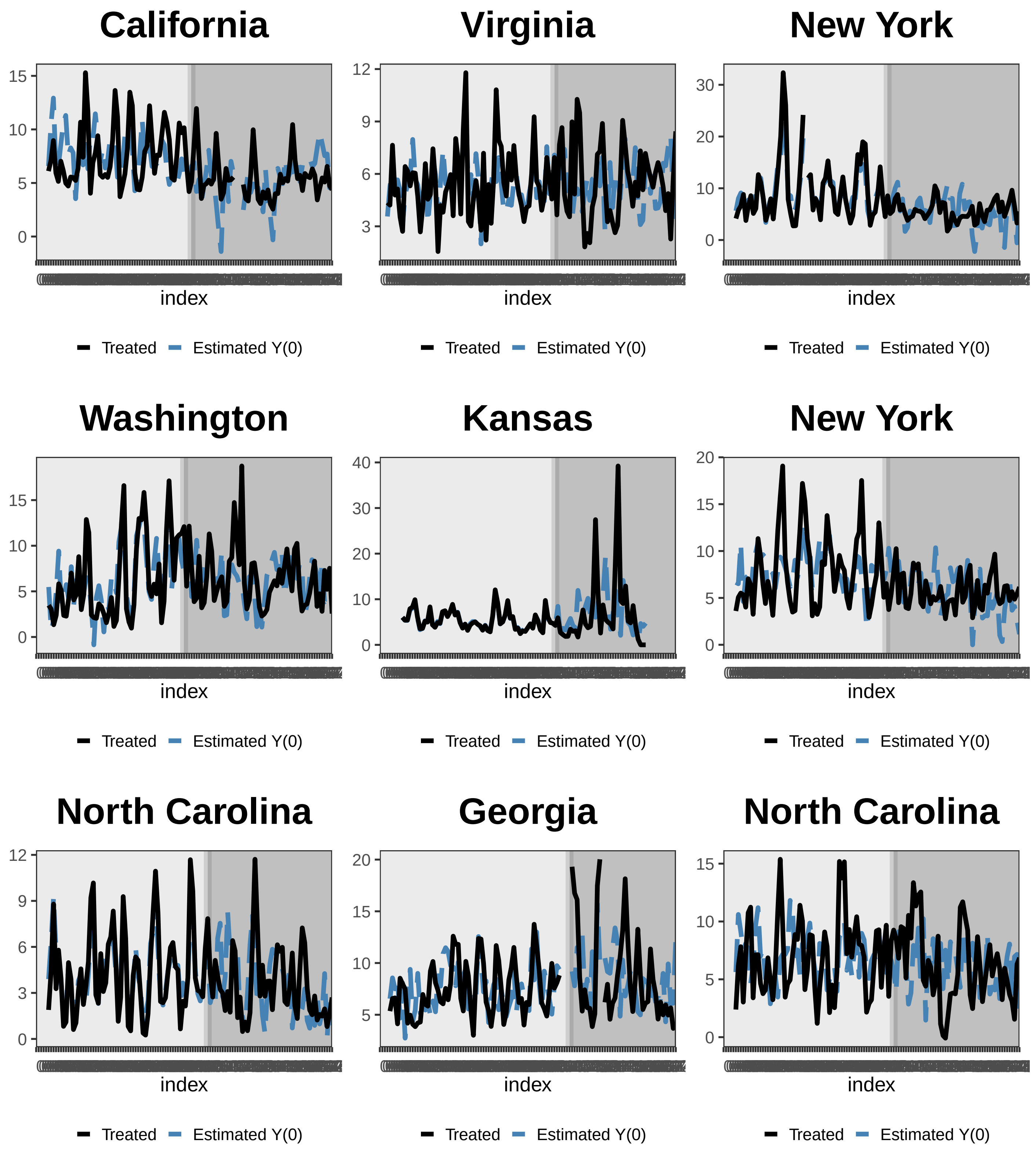
California
15
10
5
0
index
Treated
Estimated Y(0)
Virginia
12
9
6
3
index
Treated
Estimated Y(0)
New York
30
20
10
0
index
Treated
Estimated Y(0)
Washington
15
10
5
0
index
Treated
Estimated Y(0)
Kansas
40
30
20
10
0
index
Treated
Estimated Y(0)
New York
20
15
10
5
0
index
Treated
Estimated Y(0)
North Carolina
12
9
6
3
0
index
Treated
Estimated Y(0)
Georgia
20
15
10
5
index
Treated
Estimated Y(0)
North Carolina
15
10
5
0
index
Treated
Estimated Y(0)

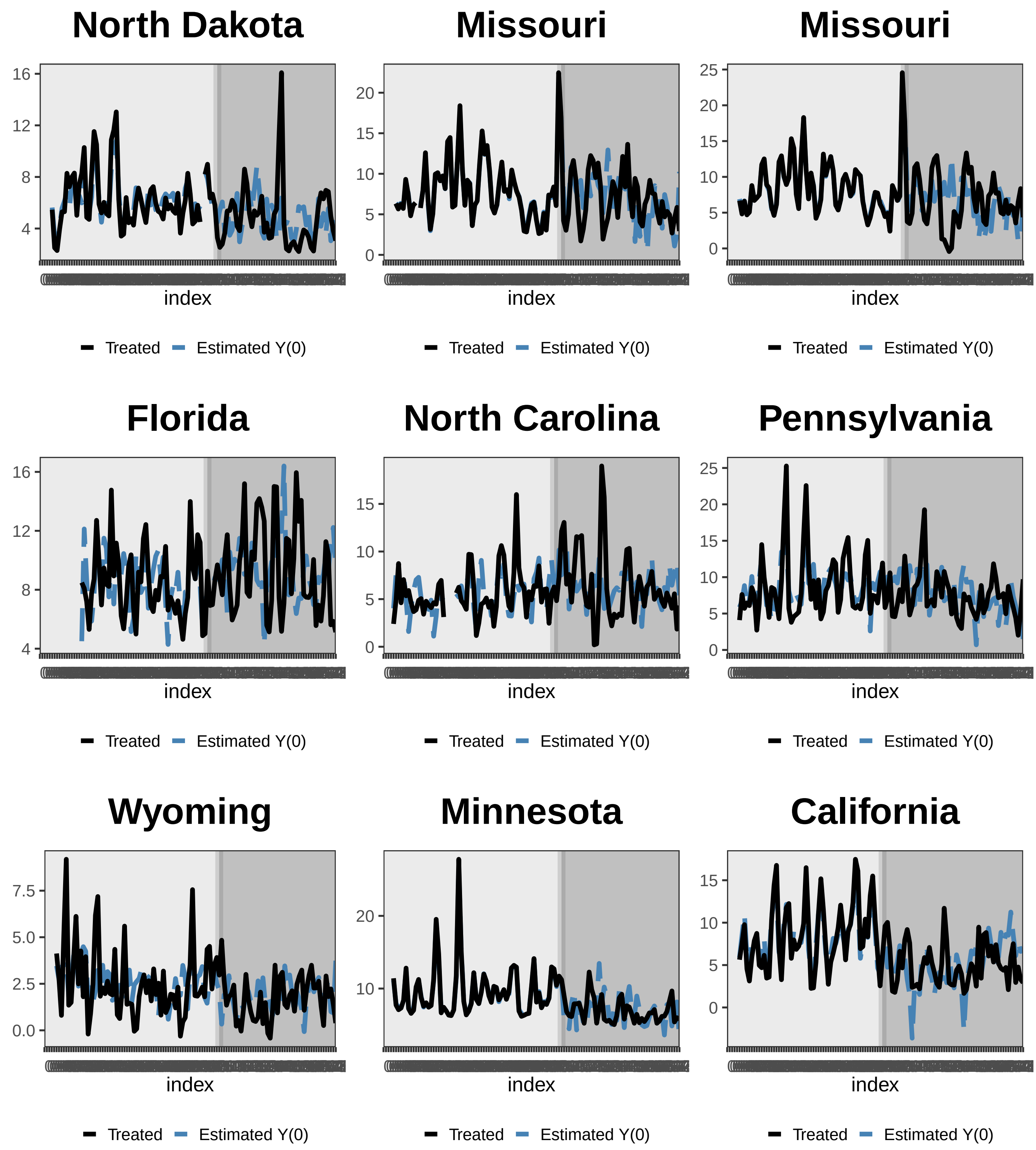
North Dakota
Missouri
Missouri
Florida
North Carolina
Pennsylvania
Wyoming
Minnesota
California
index
Treated
Estimated Y(0)

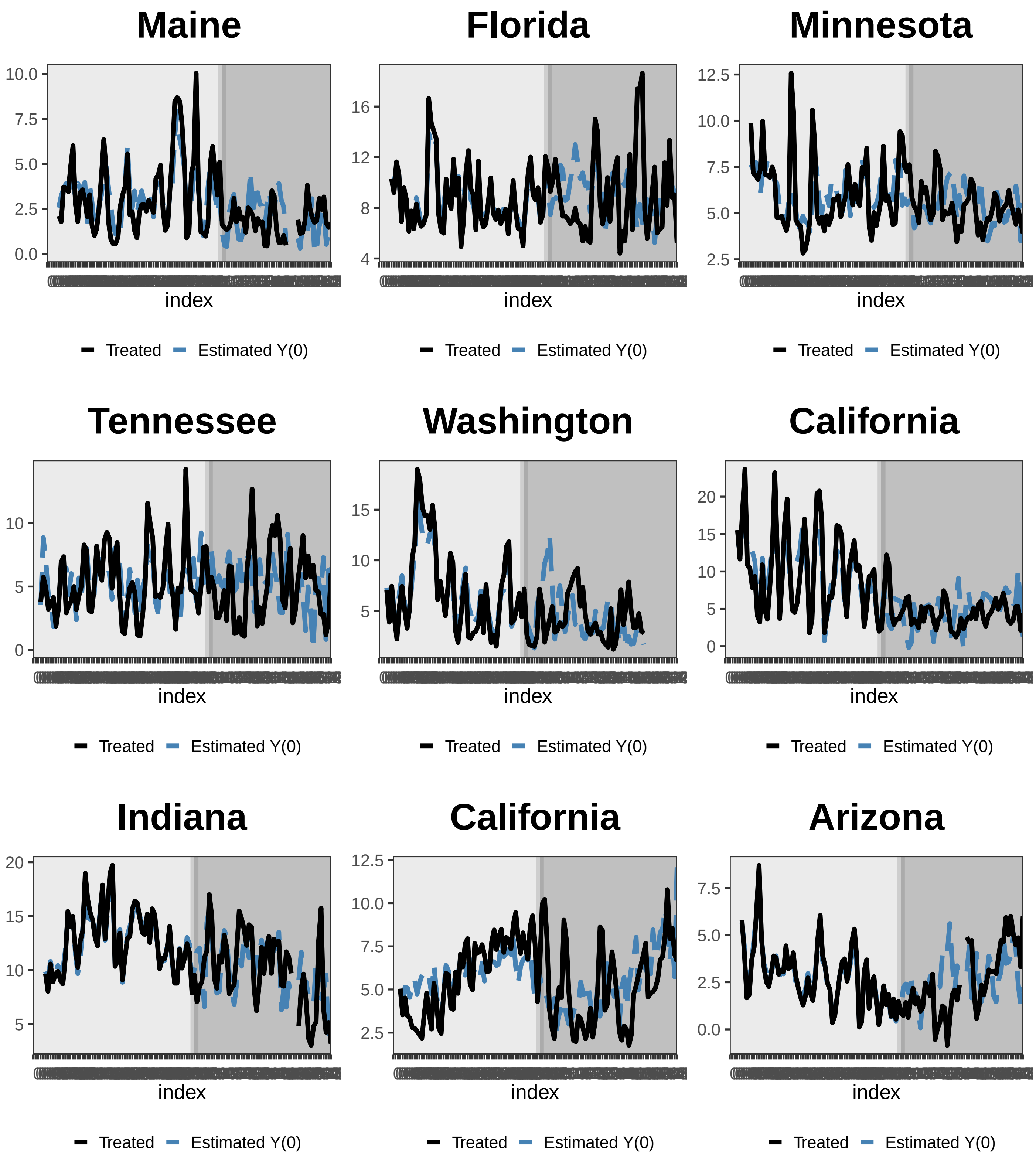
Maine
10.0
7.5
5.0
2.5
0.0
index
Treated
Estimated Y(0)
Florida
16
12
8
4
index
Treated
Estimated Y(0)
Minnesota
12.5
10.0
7.5
5.0
2.5
index
Treated
Estimated Y(0)
Tennessee
10
5
0
index
Treated
Estimated Y(0)
Washington
15
10
5
index
Treated
Estimated Y(0)
California
20
15
10
5
0
index
Treated
Estimated Y(0)
Indiana
20
15
10
5
index
Treated
Estimated Y(0)
California
12.5
10.0
7.5
5.0
2.5
index
Treated
Estimated Y(0)
Arizona
7.5
5.0
2.5
0.0
index
Treated
Estimated Y(0)

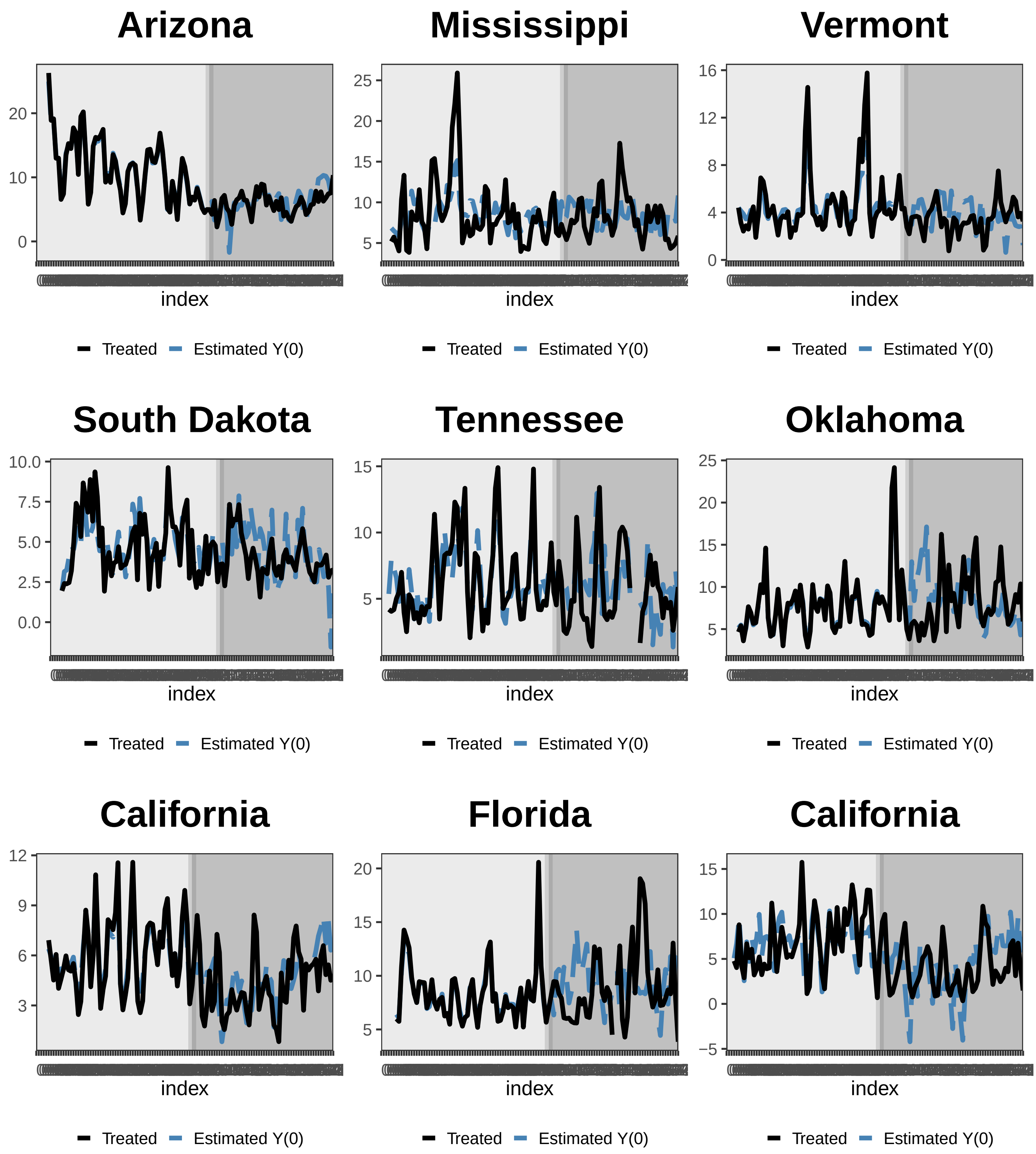
Arizona
index
Treated
Estimated Y(0)
Mississippi
index
Treated
Estimated Y(0)
Vermont
index
Treated
Estimated Y(0)
South Dakota
index
Treated
Estimated Y(0)
Tennessee
index
Treated
Estimated Y(0)
Oklahoma
index
Treated
Estimated Y(0)
California
index
Treated
Estimated Y(0)
Florida
index
Treated
Estimated Y(0)
California
index
Treated
Estimated Y(0)

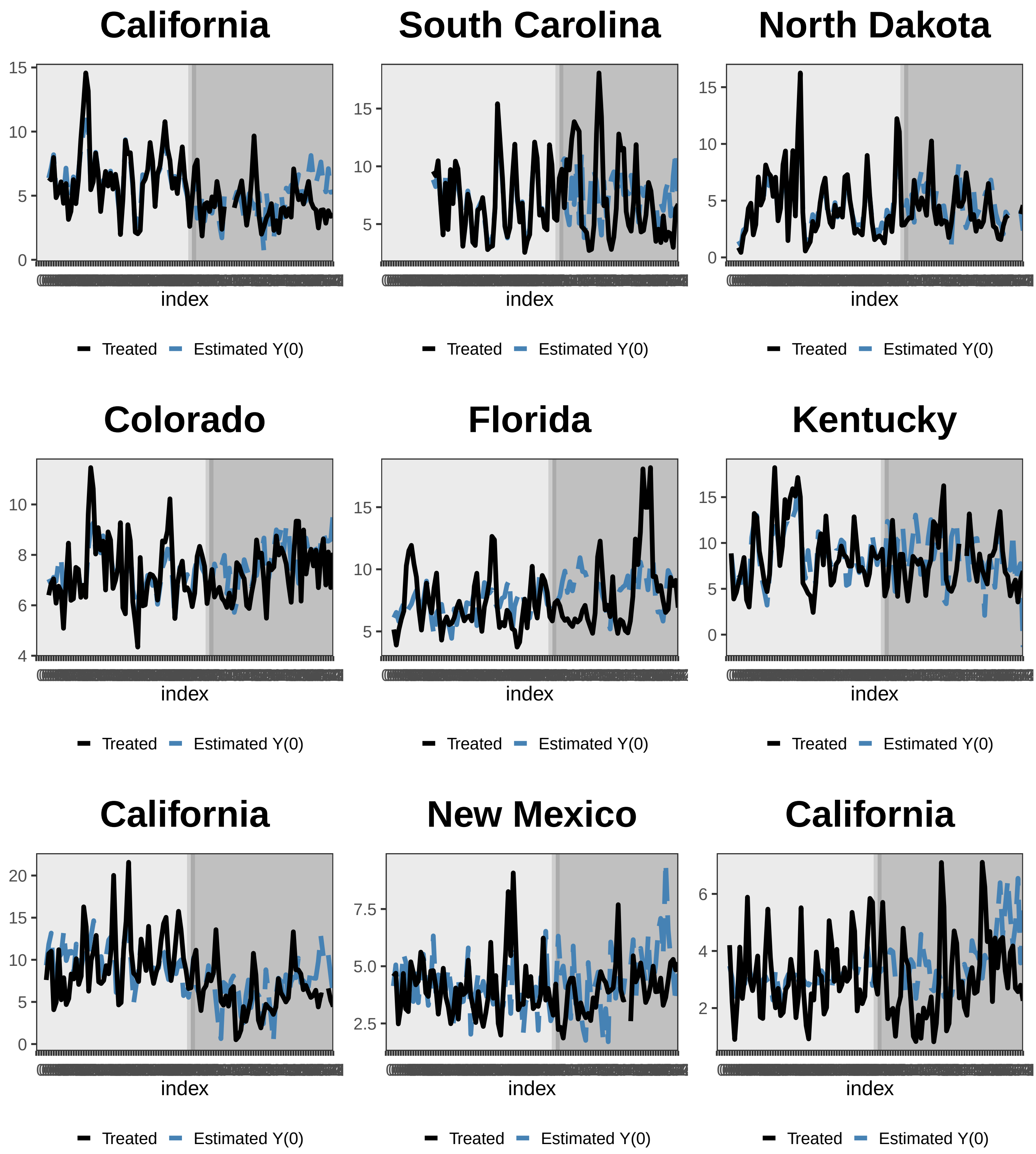
California
15
10
5
0
index
Treated
Estimated Y(0)
South Carolina
15
10
5
index
Treated
Estimated Y(0)
North Dakota
15
10
5
0
index
Treated
Estimated Y(0)
Colorado
10
8
6
4
index
Treated
Estimated Y(0)
Florida
15
10
5
index
Treated
Estimated Y(0)
Kentucky
15
10
5
0
index
Treated
Estimated Y(0)
California
20
15
10
5
0
index
Treated
Estimated Y(0)
New Mexico
7.5
5.0
2.5
index
Treated
Estimated Y(0)
California
6
4
2
index
Treated
Estimated Y(0)

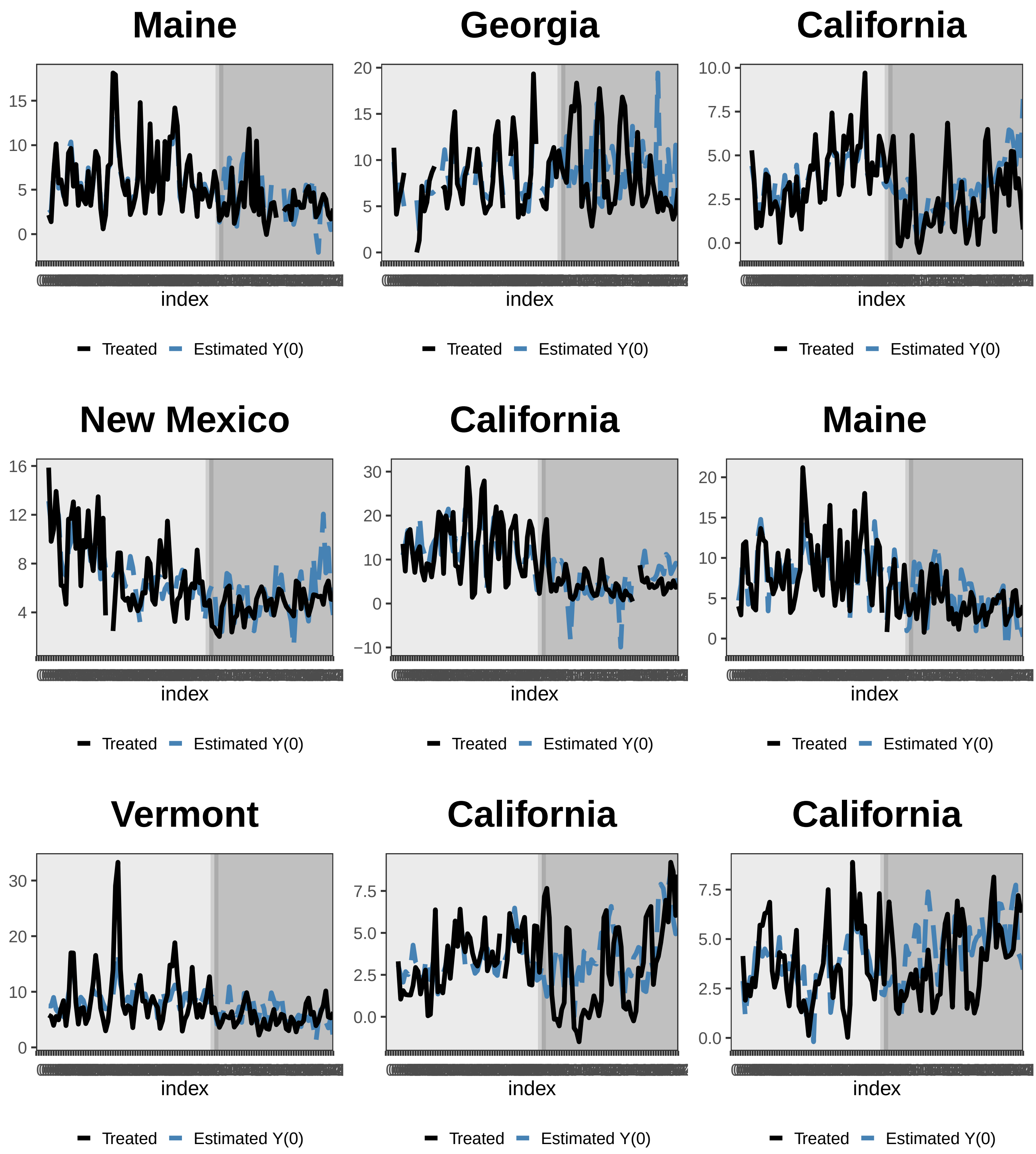
Maine
15
10
5
0
index
Treated
Estimated Y(0)
Georgia
20
15
10
5
0
index
Treated
Estimated Y(0)
California
10.0
7.5
5.0
2.5
0.0
index
Treated
Estimated Y(0)
New Mexico
16
12
8
4
index
Treated
Estimated Y(0)
California
30
20
10
0
−10
index
Treated
Estimated Y(0)
Maine
20
15
10
5
0
index
Treated
Estimated Y(0)
Vermont
30
20
10
0
index
Treated
Estimated Y(0)
California
7.5
5.0
2.5
0.0
index
Treated
Estimated Y(0)
California
7.5
5.0
2.5
0.0
index
Treated
Estimated Y(0)

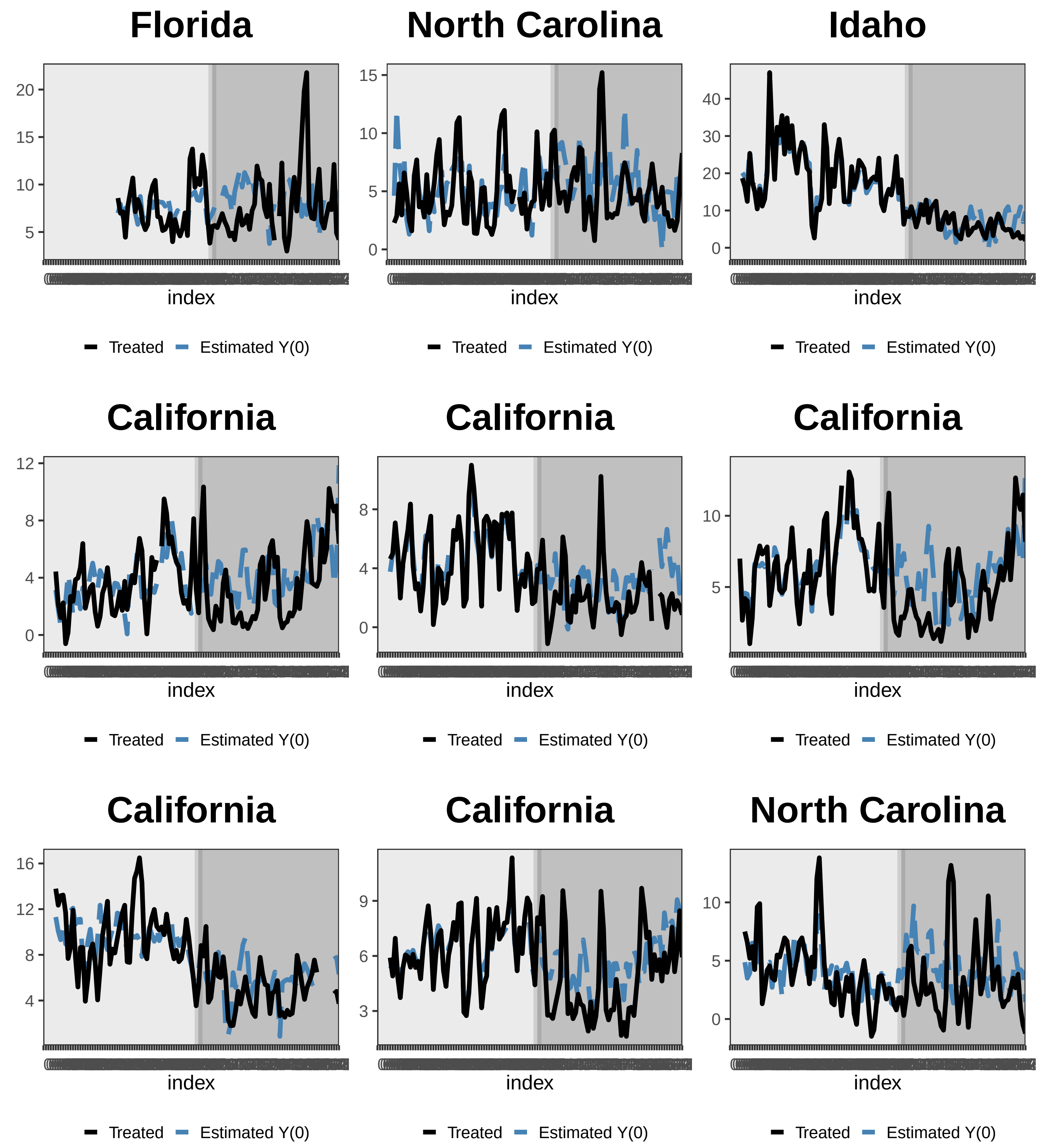
Florida
North Carolina
Idaho
California
California
California
California
California
North Carolina
index
Treated
Estimated Y(0)

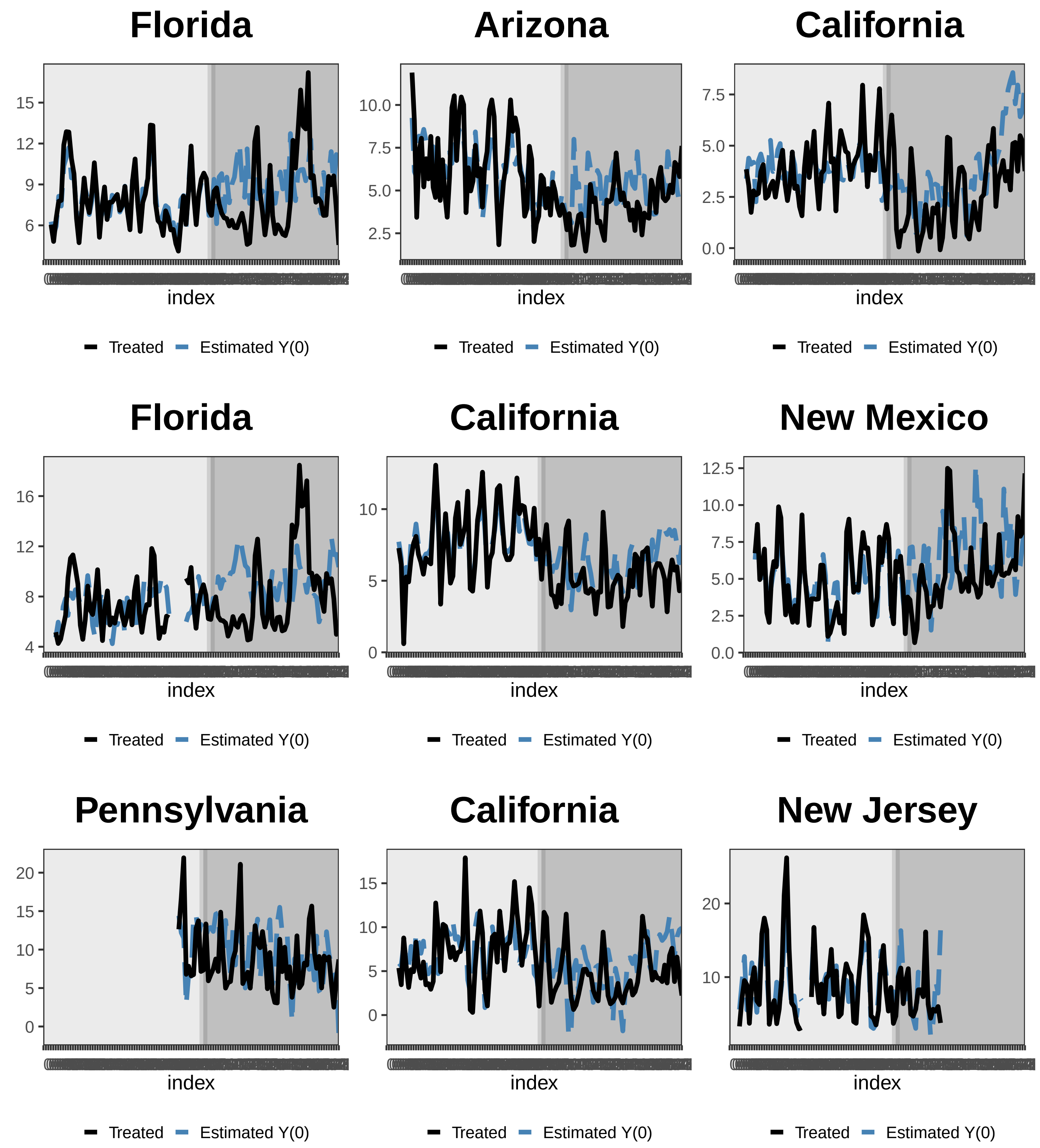
Florida
15
12
9
6
index
Treated
Estimated Y(0)
Arizona
10.0
7.5
5.0
2.5
index
Treated
Estimated Y(0)
California
7.5
5.0
2.5
0.0
index
Treated
Estimated Y(0)
Florida
16
12
8
4
index
Treated
Estimated Y(0)
California
10
5
0
index
Treated
Estimated Y(0)
New Mexico
12.5
10.0
7.5
5.0
2.5
0.0
index
Treated
Estimated Y(0)
Pennsylvania
20
15
10
5
0
index
Treated
Estimated Y(0)
California
15
10
5
0
index
Treated
Estimated Y(0)
New Jersey
20
10
index
Treated
Estimated Y(0)

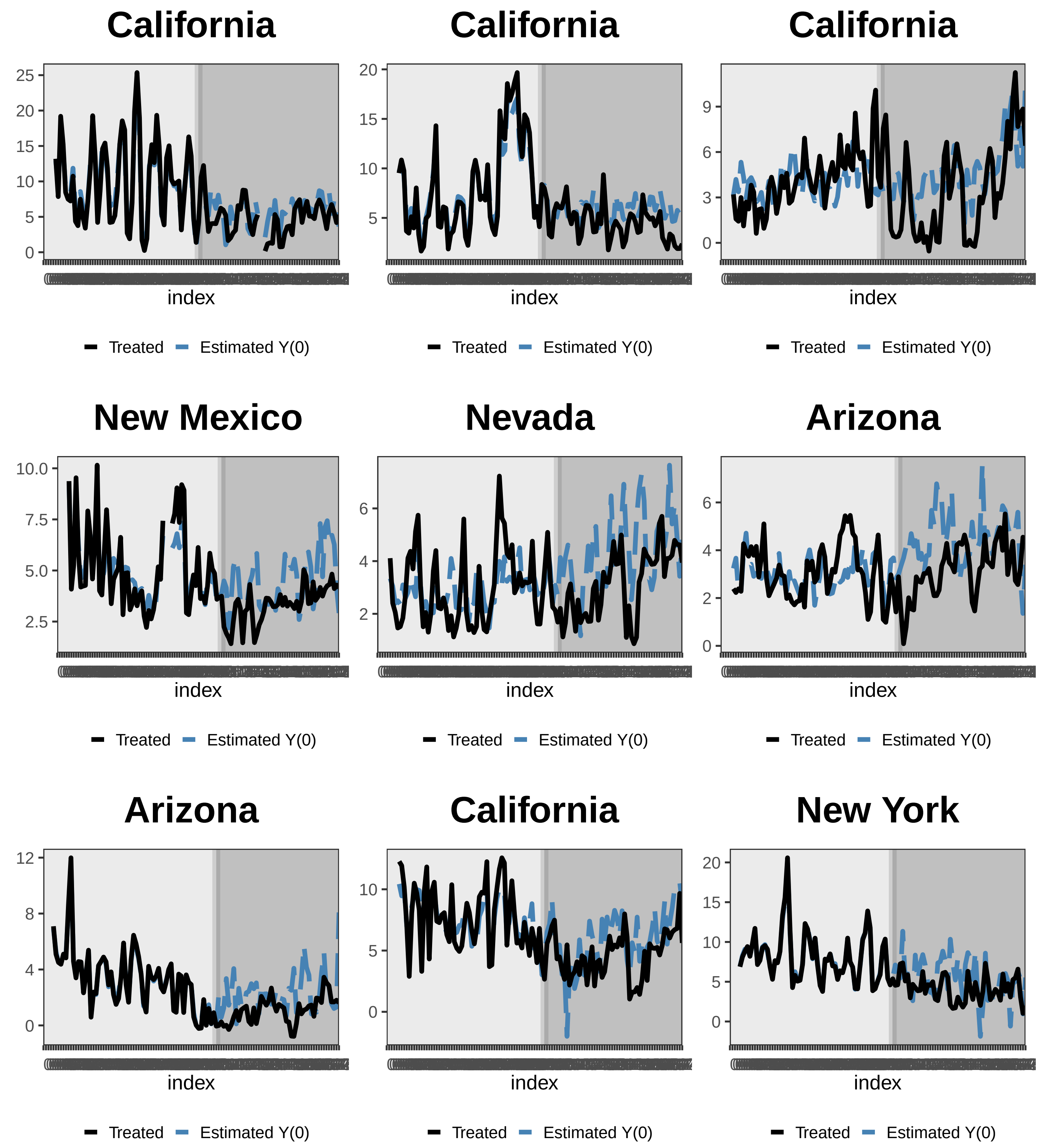
California
25
20
15
10
5
0
index
Treated
Estimated Y(0)
California
20
15
10
5
index
Treated
Estimated Y(0)
California
9
6
3
0
index
Treated
Estimated Y(0)
New Mexico
10.0
7.5
5.0
2.5
index
Treated
Estimated Y(0)
Nevada
6
4
2
index
Treated
Estimated Y(0)
Arizona
6
4
2
0
index
Treated
Estimated Y(0)
Arizona
12
8
4
0
index
Treated
Estimated Y(0)
California
10
5
0
index
Treated
Estimated Y(0)
New York
20
15
10
5
0
index
Treated
Estimated Y(0)

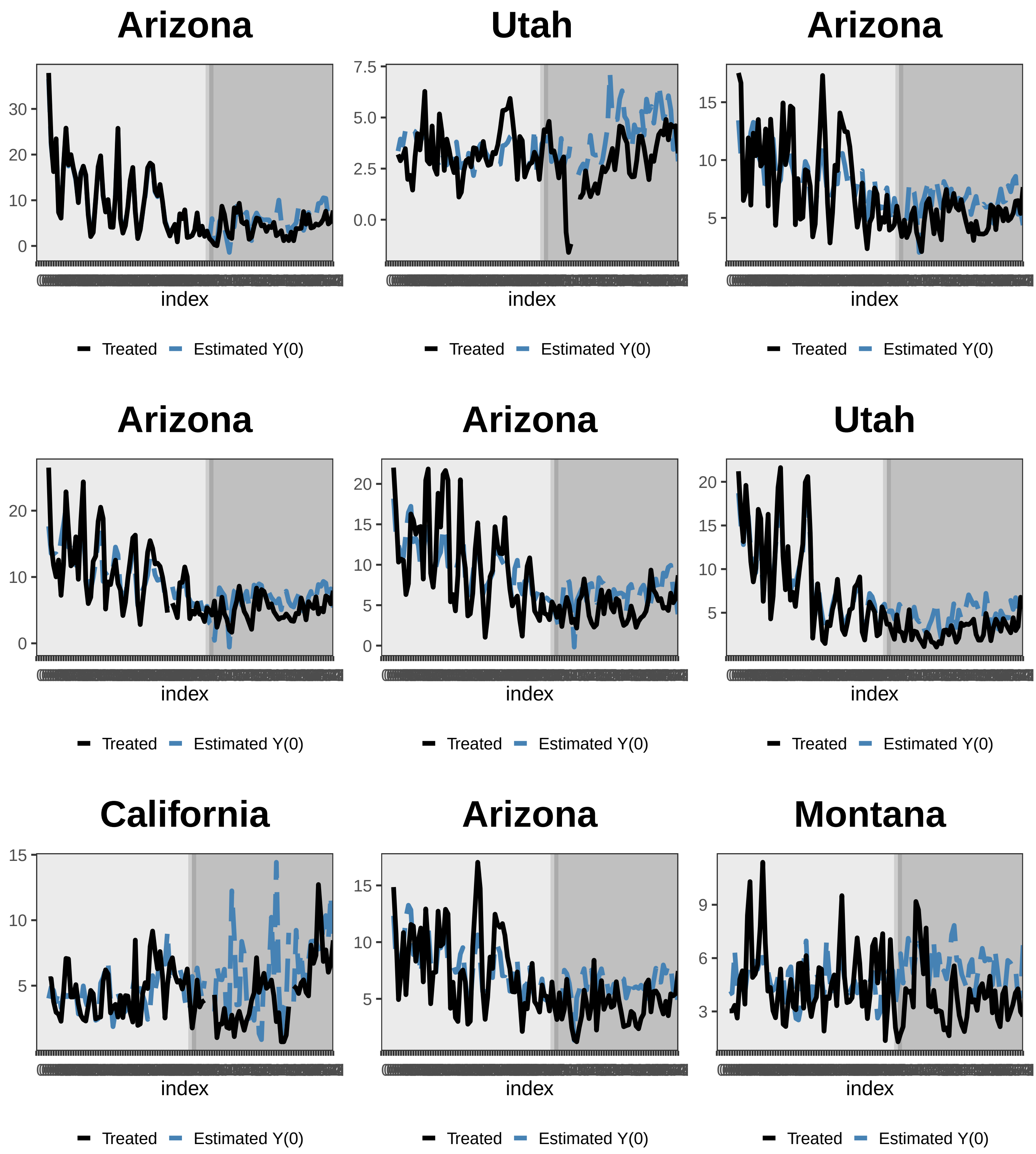
Arizona
30
20
10
0
index
Treated
Estimated Y(0)
Utah
7.5
5.0
2.5
0.0
index
Treated
Estimated Y(0)
Arizona
15
10
5
index
Treated
Estimated Y(0)
Arizona
20
10
0
index
Treated
Estimated Y(0)
Arizona
20
15
10
5
0
index
Treated
Estimated Y(0)
Utah
20
15
10
5
index
Treated
Estimated Y(0)
California
15
10
5
index
Treated
Estimated Y(0)
Arizona
15
10
5
index
Treated
Estimated Y(0)
Montana
9
6
3
index
Treated
Estimated Y(0)

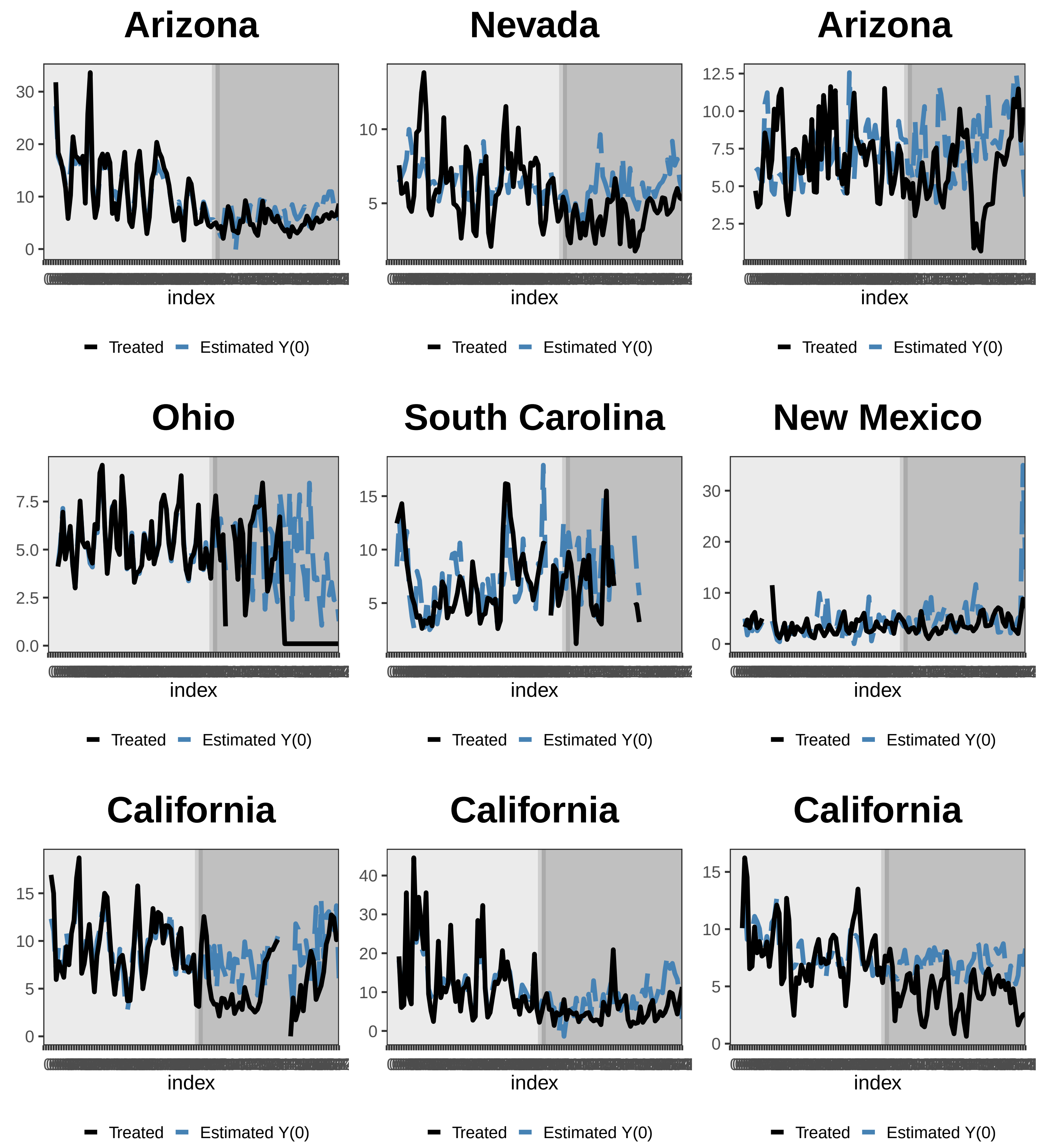
Arizona
Nevada
Arizona
Ohio
South Carolina
New Mexico
California
California
California
index
Treated
Estimated Y(0)

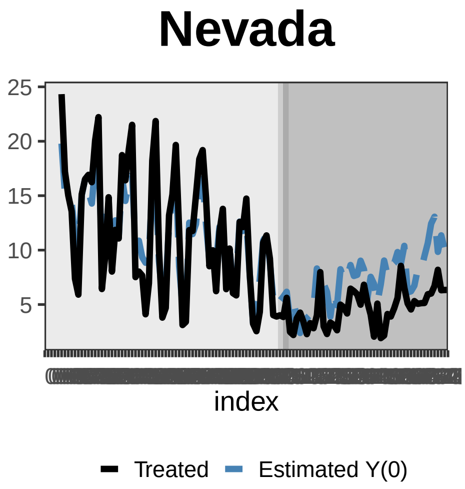
Nevada
25
20
15
10
5
index
Treated
Estimated Y(0)

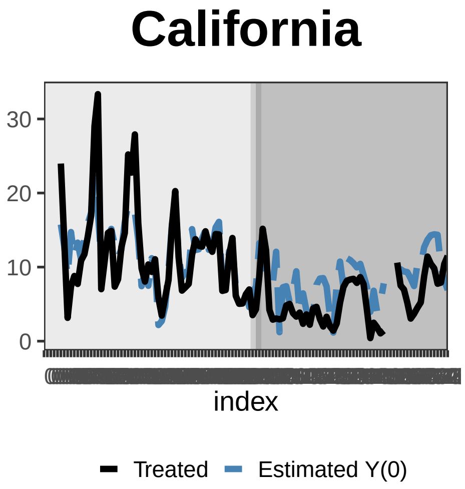
California
30
20
10
0
index
Treated
Estimated Y(0)

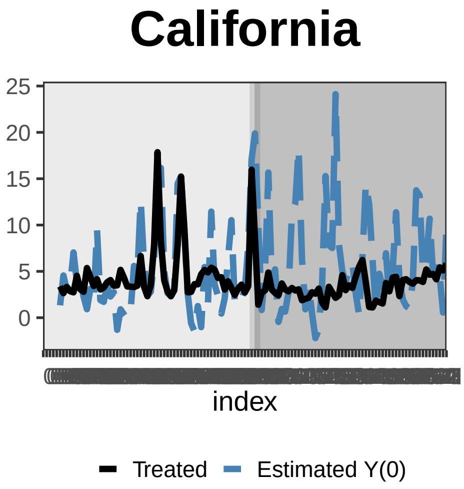
California
25
20
15
10
5
0
index
Treated
Estimated Y(0)

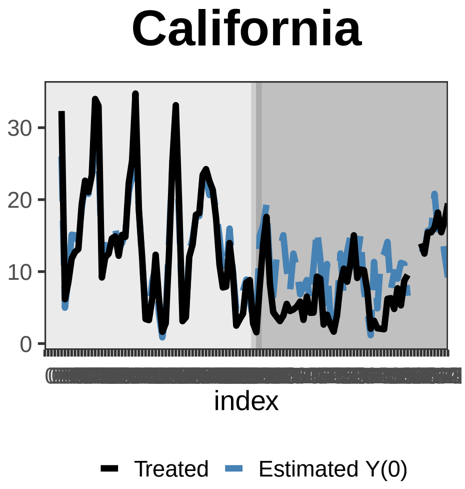
California
30
20
10
0
index
Treated
Estimated Y(0)

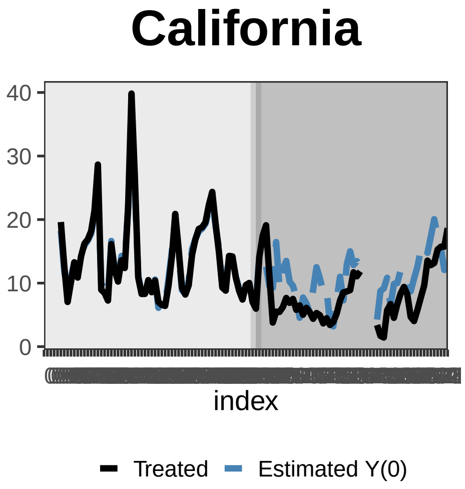
California
40
30
20
10
0
index
Treated
Estimated Y(0)

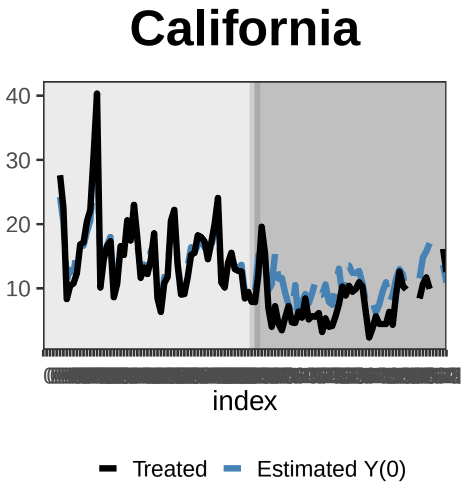
California
40
30
20
10
index
Treated
Estimated Y(0)

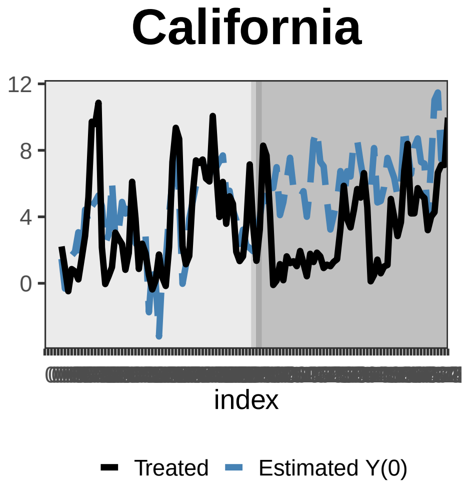
California
12
8
4
0
index
Treated
Estimated Y(0)

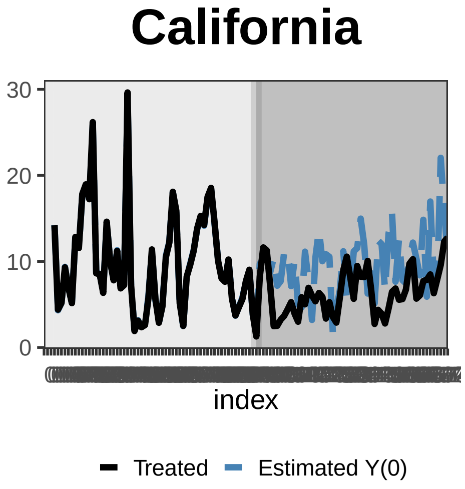
California
30
20
10
0
index
Treated
Estimated Y(0)

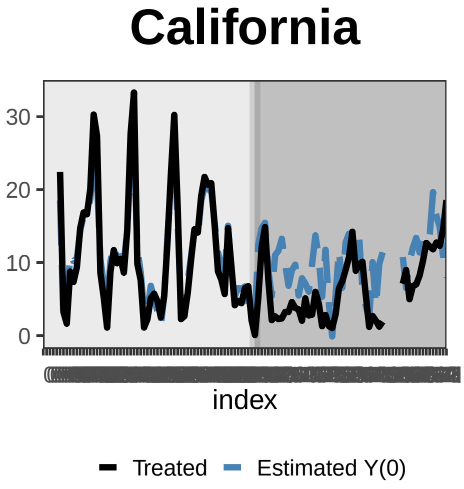
California
30
20
10
0
index
Treated
Estimated Y(0)

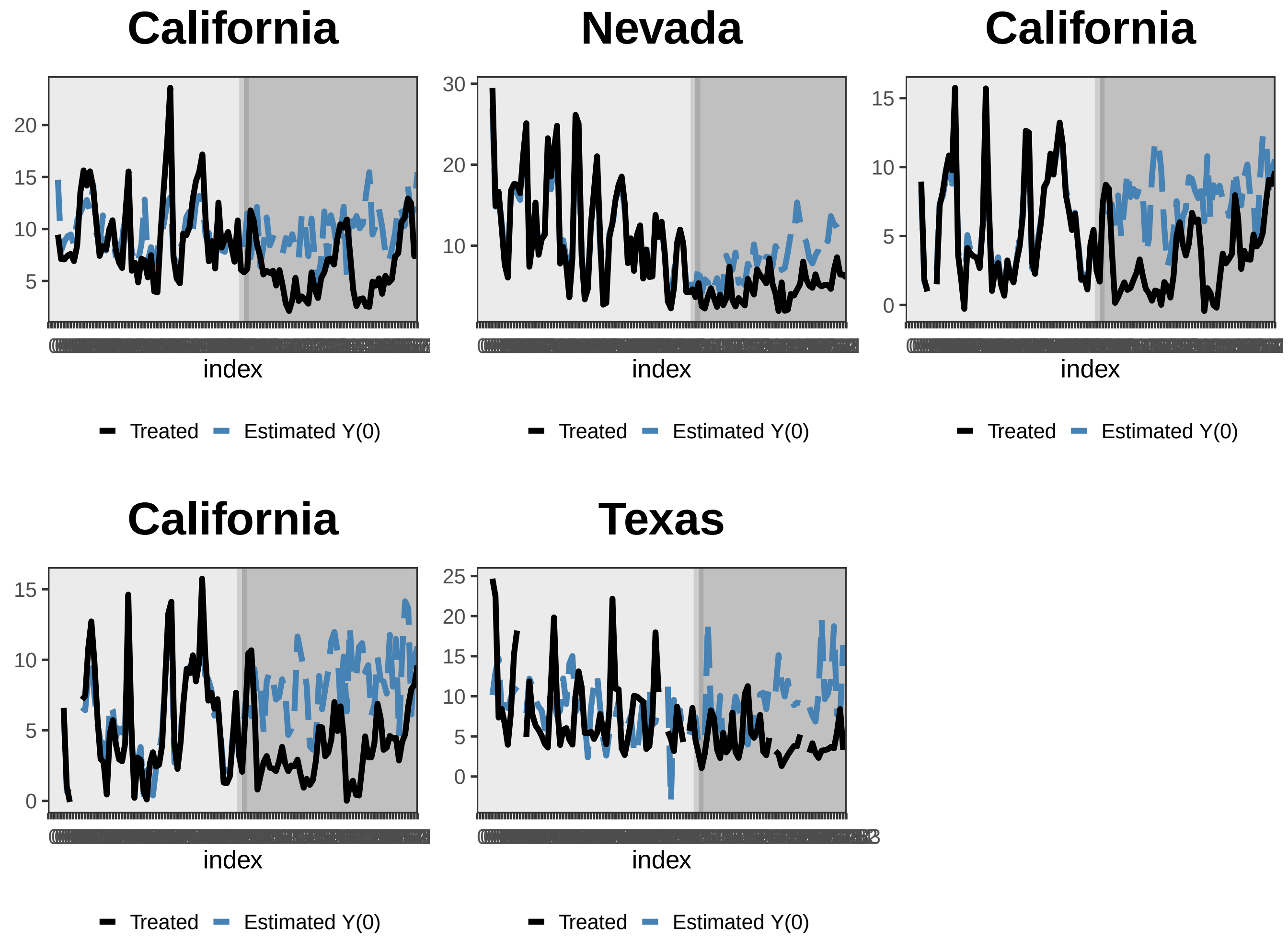


**Figure 1.** Plots of observed and counterfactual $PM_{2.5}$ concentrations at each of the 455 monitor sites in 2020. The black lines shown are the observed levels, the blue dotted lines are the counterfactuals, and dark gray areas indicate the lockdown period while light gray areas indicate the pre-lockdown period in 2020.
